# Antibiotic Resistance Spread and Resistance Control Options. Estonian Experience

**DOI:** 10.1101/2024.04.02.24304316

**Authors:** Tanel Tenson, Kaidi Telling, Piret Mitt, Epp Sepp, Paul Naaber, Jana Lass, Irja Lutsar, Piret Kalmus, Epp Moks, Liidia Häkkinen, Veljo Kisand, Koit Herodes, Age Brauer, Maido Remm, Ülar Allas

**Affiliations:** University of Tartu; Estonian University of Life Sciences; Veterinary and Food Laboratory; Tartu University Hospital; Synlab OÜ

## Abstract

Antibiotic resistance refers to the ability of microbes to grow in the presence of an antibiotic that would have originally killed or inhibited the growth of these microorganisms. Microorganisms resistant to antibiotics exist in humans, animals and in the environment. Resistant microbes can spread from animals to humans and vice versa either through direct contact or through the environment. Resistant bacteria survive in the body during a course of antibiotics and continue to multiply. Treatment of antibiotic-resistant infections takes more time, costs more, and sometimes may prove impossible.

The aim of the AMR-RITA project was to develop recommendations based on scientific evidence including the “One Health” principle for the formulation of policy on antibiotic resistance. In order to achieve the goal, the role of human behaviour, human and animal medicine, and the environment was implicated in the development of antibiotic resistance. The evaluation of the resistance spread routes, risks and levels, and the possible measures to control the spread of antibiotic resistance were identified.

Topics related to antibiotic resistance were analysed in medicine, veterinary medicine and environment subsections. Existing data were combined with new data to assess the transmission routes and mechanisms of antibiotic resistance. For this purpose, samples were collected from people, animals, food, and the environment. The analysis of the samples focused on the main resistent organisms, resistance genes and antibiotic residues.

As a result of the study, we conclude that the use of antibiotics in Estonia is generally low compared to other European countries. However, there are bottlenecks that concern both human and veterinary medicine. In both cases, we admit that for some diagnoses there were no treatment guidelines and antibiotics were used for the wrong indications. The lack of specialists of clinical microbiology is a problem in Estonain hospitals. For example, many hospitals lack an infection control specialist. The major worrying trends are the unwarranted use of broad-spectrum antibiotics in humans and the high use of antibiotics critical for human medicine (cephalosporins, quinolones) in the teratment of animals.

If more antibiotics are being used, resistance will also spread. We found that those cattle farms that use more cephalosporins also have higher levels of resistance (ESBL-mediated resistance). It also turned out that genetically close clusters of bacteria are often shared by humans and animals. This is evidence of a transfer of resistance between species. However, such transfer occurs slowly, and we did not detect any transfer events in the recent years.

Antibiotic residues, just like other drug residues, can reach the environment. The use of slurry and composted sewage sludge as fertilizer are the main pathways. We detected fluroquinolones and tetracyclines in comparable concentrations in slurry and uncomposted sewage sludge. Composting reduces the content of drug residues, and the efficiency of the process depends on the technology used. In addition to antibiotic residues, we also determined some other drug residues accumulating in the environment. High levels of diclofenac and carbamazepine in surface water are a special concern. These are medicines for human use only, so they reach the environment through sewage treatment plants.

Based on the results obtained during the research, we propose a series of evidence-based recommendations to the state for the formulation of antimicrobial resistance policy. We propose that Estonia needs sustainable AMR surveillance institution, which (1) continuously collects and analyses data on the use of antimicrobials and antimicrobial resistance and provides regular feedback to relevant institutions (state, health and research institutions), (2) assesses the reliability of the data and ensures carrying out additional and confirming studies, (3) coordinates the activities of national and international research and monitoring networks and projects. We recommend creation of a competence centre that would deal with the topic of AMR across all fields. This should also include funding for research.

## 1. Introduction

Antibiotic resistance (AMR) refers to the ability of microorganisms to grow in the presence of antibiotics, which would have originally killed or inhibited these microorganisms. Resistant microorganisms exist in humans, animals, and the environment. Resistant microorganisms can spread from animals to humans and vice versa, either through direct contact or environmental routes. Resistant bacteria survive in the body during antibiotic treatment and continue to reproduce. Treating antibiotic-resistant infections takes longer, costs more, and can sometimes even be impossible. A recent study showed that in the United States alone, infections caused by multidrug-resistant bacteria cost $1.9 billion in healthcare expenses and more than 400,000 hospital days each year (Nelson et al., 2022). In the European Union and European Economic Area countries, AMR causes nearly 33,000 deaths per year and costs healthcare systems approximately €1.1 billion annually (OECD, 2019).

The most commonly used antibacterial agents in medicine are amoxicillin + clavulanic acid, doxycycline, amoxicillin, clarithromycin, and cefuroxime (Estonian Medicines Agency, 2022). Unmetabolized antibiotic residues enter the sewage from healthcare facilities and households through urine and feces, which are then directed to wastewater treatment plants. The technology used in wastewater treatment plants generally does not yet allow for the complete removal of micro-pollutants. Therefore, antibiotic residues can re-enter the water circulation from wastewater treatment plants. An additional problem arises when sewage is used for the disposal of unused medications.

Antibiotics are used in veterinary medicine for three purposes: 1) the treatment of sick animals and metaphylaxis, 2) as a prophylactic measure in disease prevention, and 3) as a growth promoter. Antibiotic residues enter the environment with manure and sludge. In the European Union, the use of antibiotics as growth promoters is prohibited, as is prophylactic use, but in many countries around the world, this practice is still allowed. Beta-lactam antibiotics, macrolides, and tetracyclines are the most common antibiotics administered to animals. These antibiotics are also used in human medicine.

Environmental contamination caused by drug residues is a growing problem, posing potential risks to ecosystems and human health. In addition to promoting the development and spread of drug resistance, antibiotic residues in the environment can be absorbed by plants, disrupt their growth, and cause other ecotoxicological effects. Antibiotic residues in water bodies can have harmful effects on aquatic life and damage the entire ecosystem. Consumption of water or food contaminated with antibiotic residues can affect the normal functioning of the human microbiome and cause health problems, with allergies being the most common (Ben et al., 2019).

A key factor in reducing drug resistance is the optimization and control of antibiotic use. The Council of the European Union has recommended relying on the principles of the “One Health” approach in addressing the problem of resistance (Council of the European Union, 2016). This approach is based on the principle that human and animal health are interconnected because diseases are transmitted from humans to animals and vice versa. The “One Health” approach requires integrated actions to reduce antibiotic resistance in medicine, veterinary medicine, and the environment.

## 2. Objective of the Study

The aim of the AMR-RITA project was to develop scientifically based recommendations for shaping antibiotic resistance policies and reducing resistance transmission based on the “One Health” principle. The results of the project provide an overview of the situation in Estonia, but they should also be viewed in a global context. To achieve the goal, various areas, such as human and animal treatment, and the environment, were examined for their role in the development of antibiotic resistance, along with assessments of transmission routes. Possible measures to control the spread of antibiotic resistance were identified.

Antibiotic resistance-related topics were analyzed within and across medical, veterinary, and environmental sectors. Existing data were combined with new data to ensure a cross-sectoral approach and to comprehensively assess AMR transmission pathways and mechanisms. For this purpose, samples were collected from humans, animals (including manure), food, and the environment (Figures 1 and 2). The analysis of samples focused on the resistent bacterial strains, main resistance genes and antibiotic residues.

**Figure 1.**
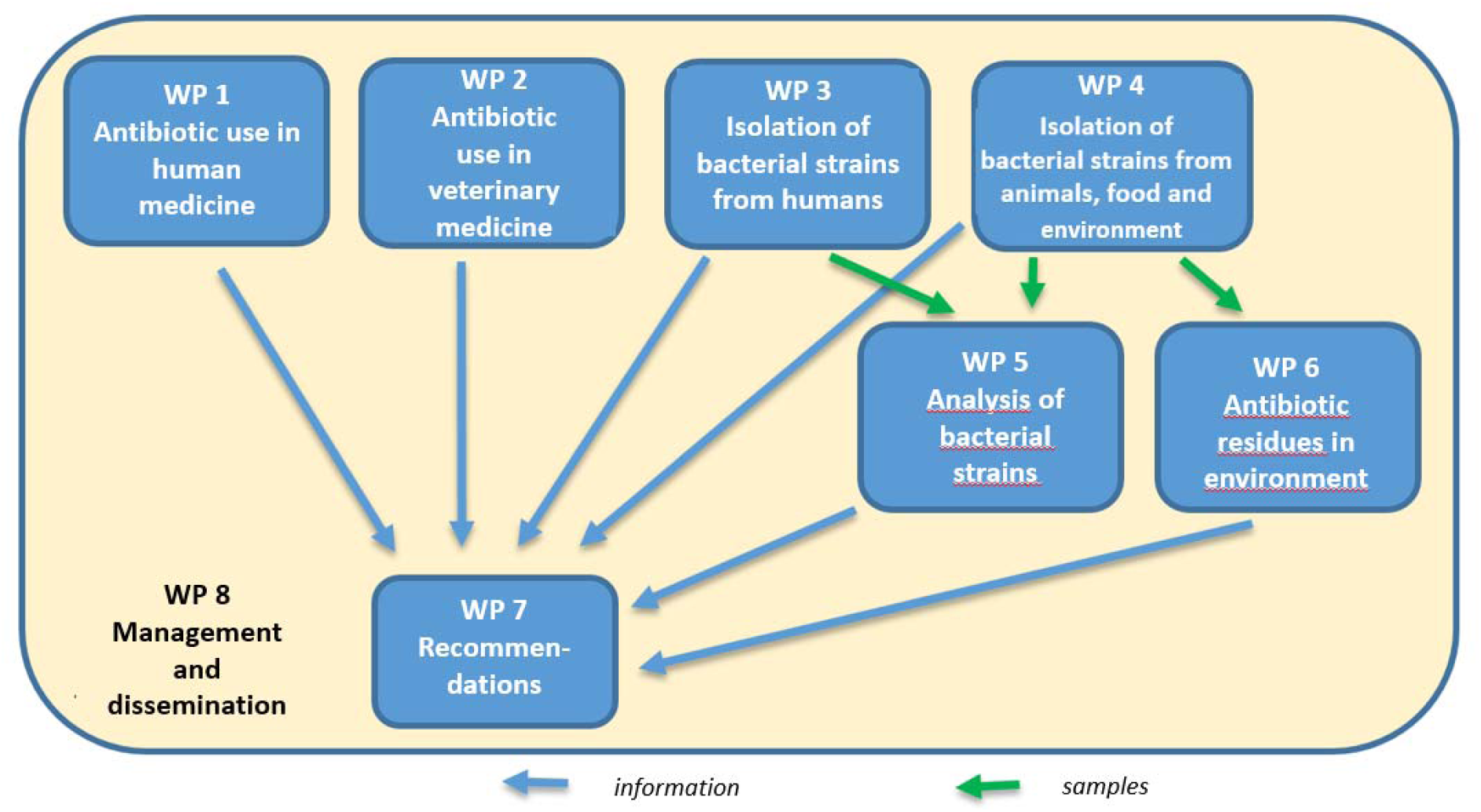
Organization of the work packages (WP) of the AMR-RITA project.

**Figure 2.**
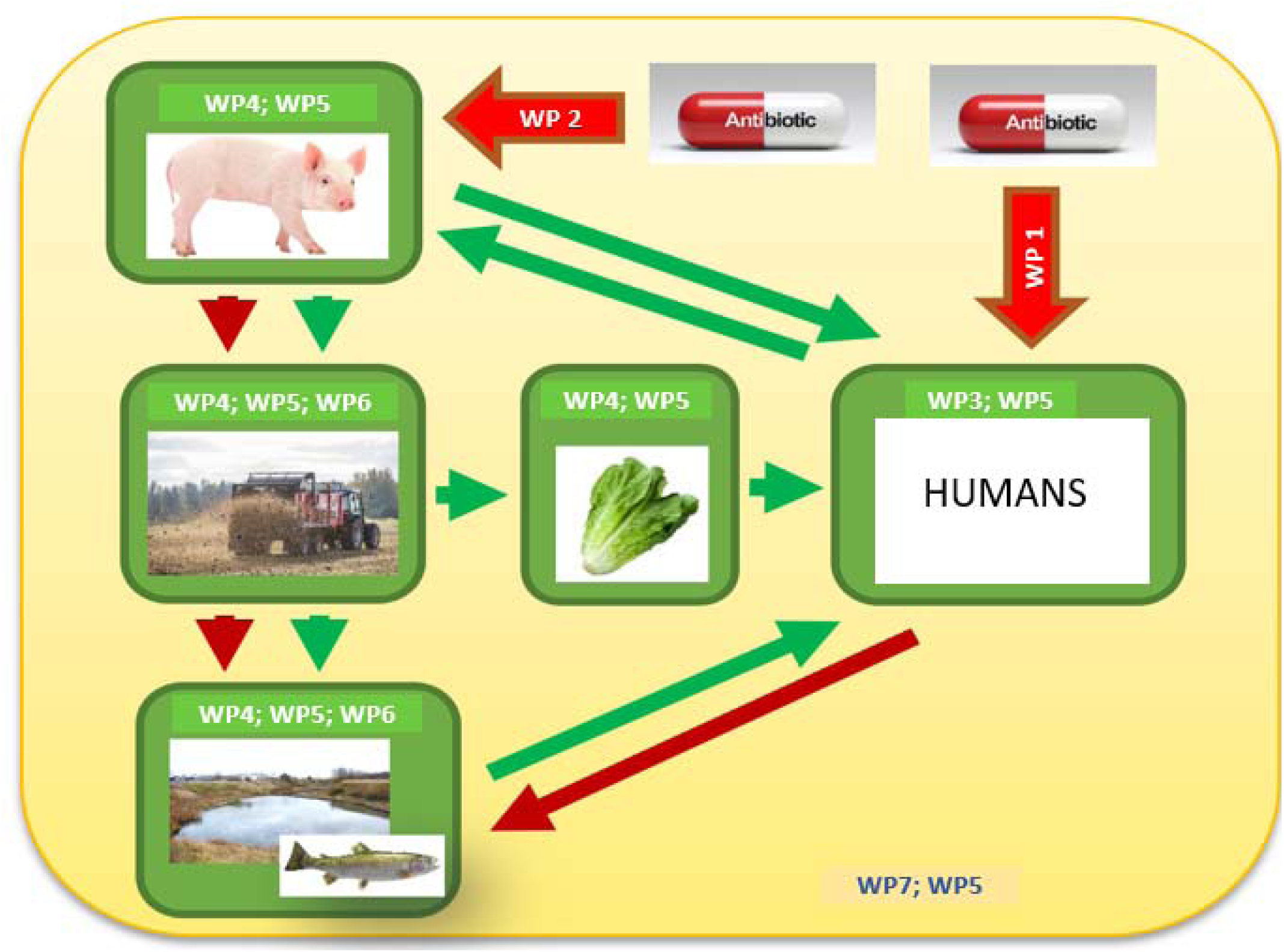
Development and spread of antibiotic resistance in relation to the work packages (WP) of the AMR-RITA project. The red arrows indicate the consumption of antibiotics and the transmission routes of antibiotic residues. Green arrows indicate transmission pathways of antibiotic-resistant bacteria and resistance genes.

Based on the results obtained during the studies, we present a series of evidence-based recommendations for shaping AMR policy in the country. The study also highlighted potential bottlenecks that may hinder the implementation of scientific recommendations.

## 3. Antibiotic Use in Veterinary Medicine

In Estonia, over fifty different antimicrobial agents are used for the veterinary purposes annually. According to the data from the Estonian State Agency of Medicines, the most sold groups of veterinary antibiotics in 2021 were tetracyclines, penicillins, and pleuromutilins. The sold quantity of antimicrobial agents decreased by 5.6% in 2021 compared to the previous year, amounting to 6.4 tons (Figure 3). The most sold active ingredients were doxycycline (1320 kg), tiamulin (1209 kg), and amoxicillin (828 kg) (Table 1). Unfortunately, it is not possible to specify antibiotic use by animal species.

**Figure 3.**
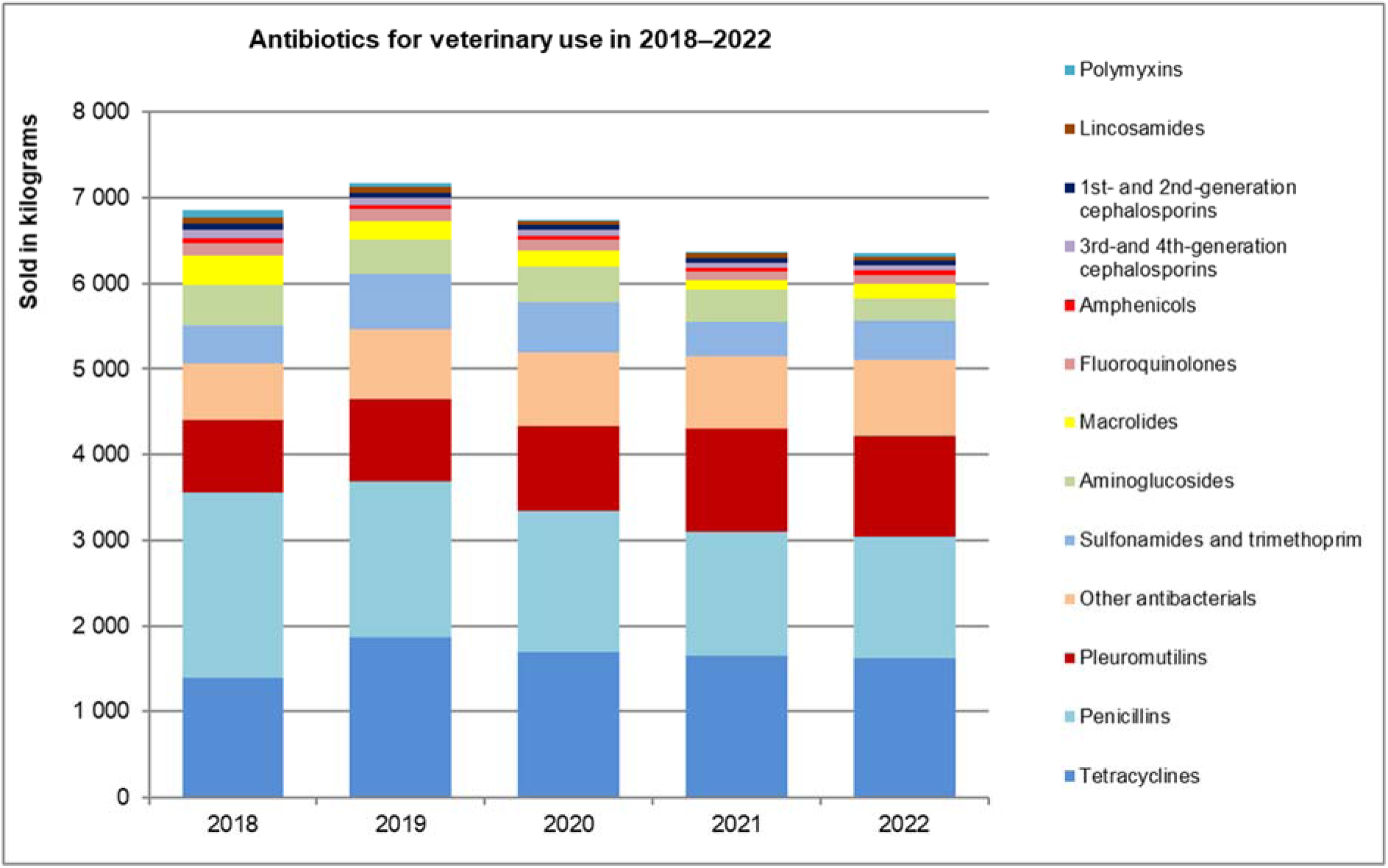
The use of antibiotics in veterinary medicine by group, considering the active substance amount sold in 2018–2022 (source: Estonian State Agency of Medicines).

**Table 1.** Sales of antibiotics for veterinary purposes, highest-selling active ingredients in 2017-2021 (Active substance sold; quantity in kilograms) (source: Estonian State Agency of Medicines).

| Active substance | 2017 | 2018 | 2019 | 2020 | 2021 |
| --- | --- | --- | --- | --- | --- |
| Doxycycline | 1380 | 1042 | 1509 | 1359 | 1320 |
| Tiamulin | 565 | 856 | 956 | 997 | 1209 |
| Amoxicillin | 1701 | 1321 | 1064 | 7050 | 828 |
| Monensin | 526 | 546 | 709 | 767 | 732 |
| Benzylpenicillin | 683 | 742 | 697 | 571 | 553 |
| Dihydrostreptomycin | 353 | 339 | 285 | 312 | 255 |
| Oxytetracycline | 178 | 242 | 245 | 220 | 241 |
| Sulfamethoxazole | 205 | 218 | 393 | 402 | 217 |

### 3.1. Antibiotics are used in the treatment of cattle diseases

Antibiotic use data were collected over a two-year period (2018-2020) in 51 cattle herds, and the treatment records for the last 12 months were analyzed. In total, 31,432 cows were kept in the study farms. The number animals kept in the study farms studied represented 38.7% of all Estonian dairy cows, making this a highly representative dataset for antibiotic treatment of Estonian dairy cattle. In the studied cattle herds, a total of 60,345 antibiotic treatment courses were conducted, averaging 56 treatment courses per 100 cows. Antibiotic courses were most frequently prescribed for the treatment of clinical mastitis (n=17,408, CI 95% 28.2-29.3) and for dry period treatment (n=17,079, CI 95% 27.4-28.6). These two reasons accounted for 57.1% of all antibiotic treatment courses. Uterine infections were treated in 12.2% (n=7,385, CI 95% 12.7-13.3) of treatment cases, and lameness in 10.9% (n=6,554, CI 95% 10.6-11.1) of treatment cases. Monensin for ketosis treatment was used 8,357 times (Figure 4).

**Figure 4.**
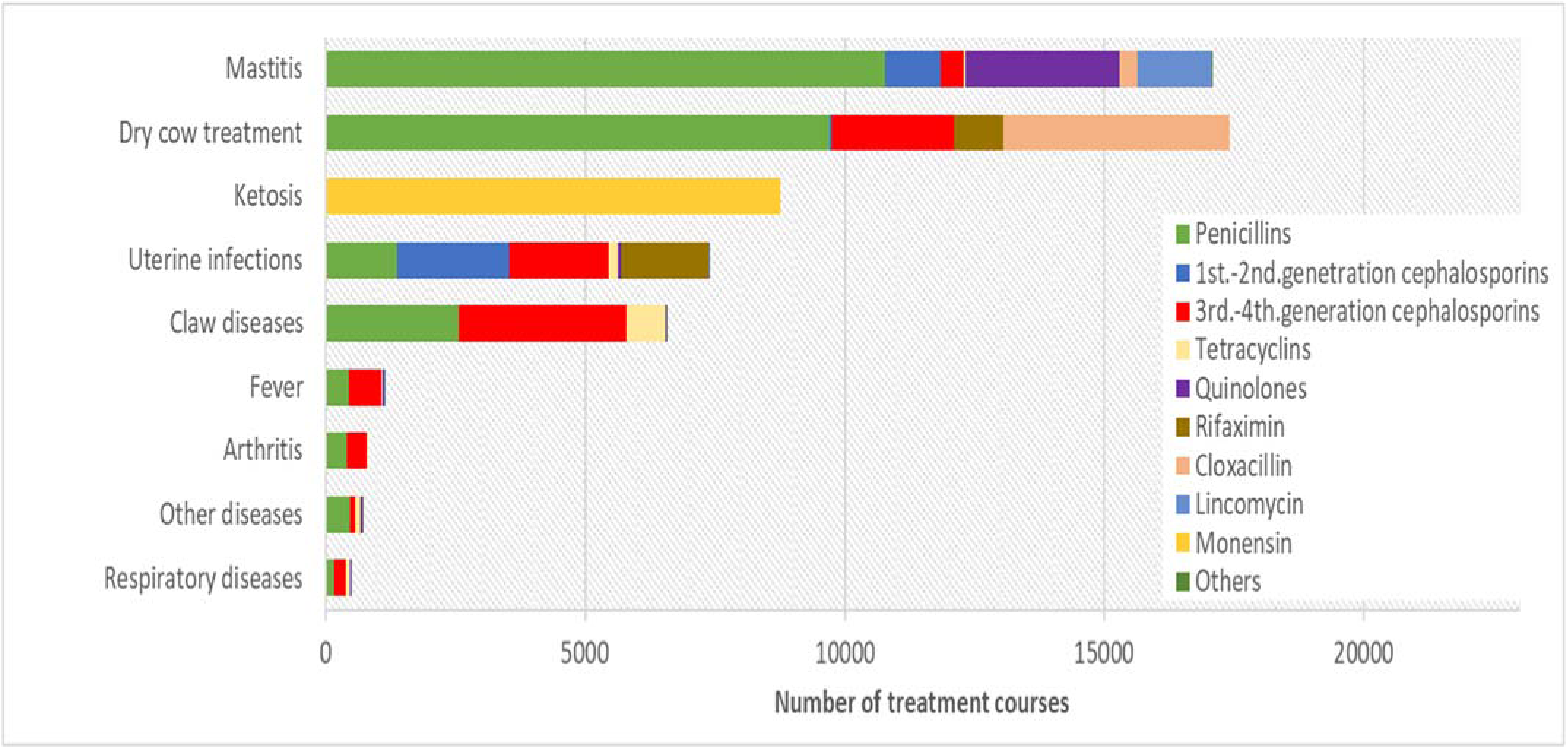
Use of different antibiotics in the treatment of cattle diseases.

Figure 4 shows that procainebenzyl penicillin was most commonly used for the treatment of clinical mastitis and dry cow treatment in cattle. The use of active ingredients was equally distributed in uterine infection treatment, but more than half of the cases of lameness were treated with 3rd and 4th generation cephalosporins.

In total, 58 different drugs containing 25 different active ingredients were used for the treatment of cattle diseases. Each drug contained one active ingredient or a combination of several different active ingredients (Table 2).

**Table 2.**
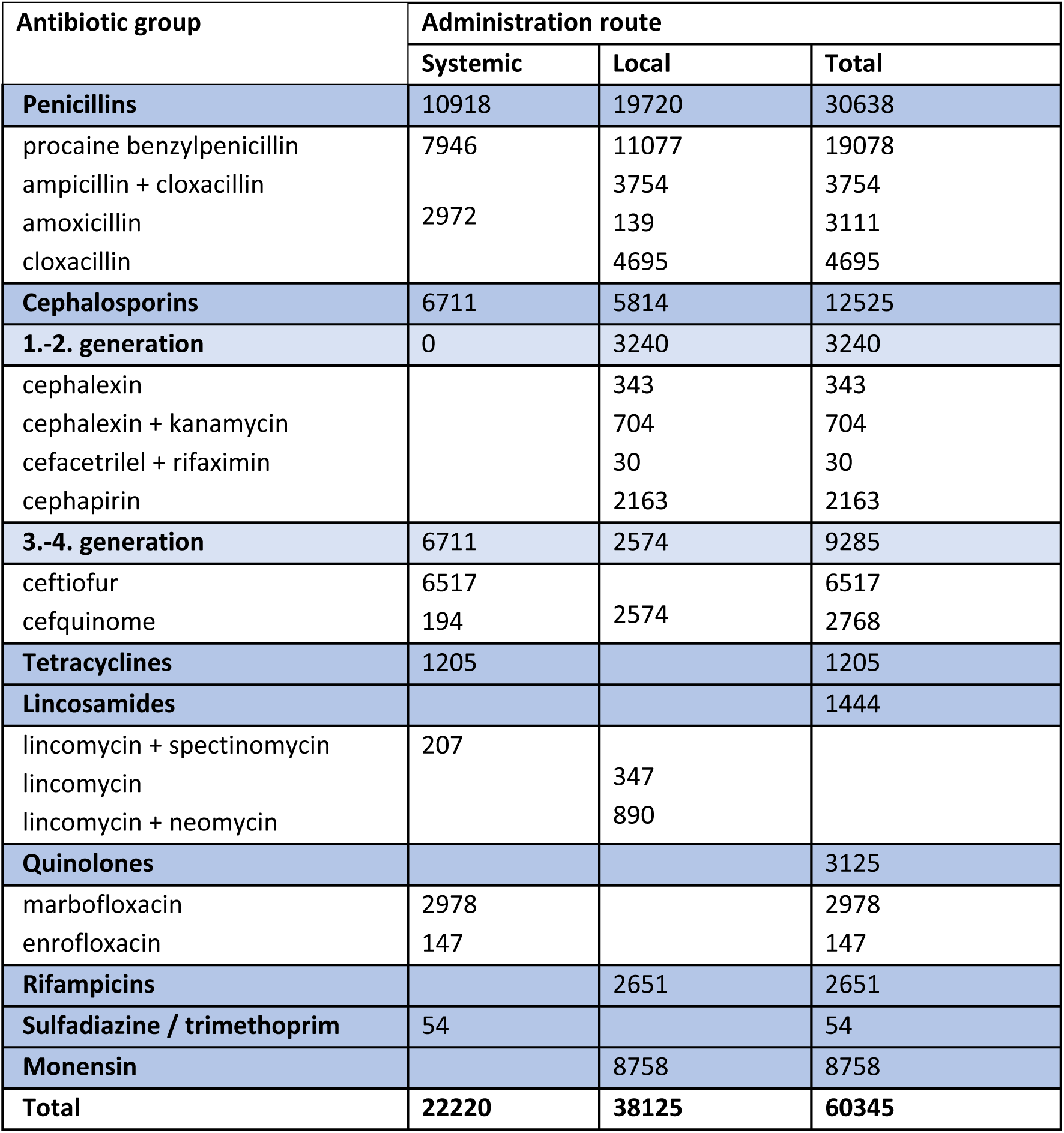
Number of treatment courses conducted with different active ingredients in 51 Estonian cattle herds.

Antibiotics were administered as systemic treatment a total of 22,220 times (37%) and as local treatment 38,125 times (63%). Excluding intraruminal monensin (n=8,785), locally administered antibiotics were used in 56.9% of cases. Veterinarians most commonly chose systemic treatment with prokain benzyl penicillins (35.7%), followed by ceftiofur (29.3%) and marbofloxacin (13.4%). Critically important antibiotics for human medicine (quinolones, 3rd and 4th generation cephalosporins) were used in systemic treatment in 9,836 treatment courses, accounting for 44.2% of all systemic treatment courses.

Data on use of critically important antibiotics for human medicine were collected over a two-year period (2018-2020), with data analysis conducted for the last 12 months. In the last 12 months, at least one cow in 47 of the study herds (92.1% of all study herds) was treated with 3rd or 4th generation cephalosporins or quinolones. Quinolones were used in 40 herds (78.4%). The quantity of active ingredients used (mg/PCU) varied between herds. The median value for the use of 3rd and 4th generation cephalosporins was 1.53 (SD = 2.5) mg/PCU and for quinolones, the median value was 0.9 (SD = 2.6) mg/PCU. The use of cephalosporins exceeded the EU median value (0.2 mg/PCU) in 38 herds, and the use of quinolones was higher than the EU median value (1.1 mg/PCU) in 24 herds.

Figure 5 demonstrates that 19 of the study herds used both cephalosporins and quinolones in quantities exceeding the EU median values, but in eight herds, the use of these antibiotics was lower than the EU median values. In the remaining herds, either cephalosporins or quinolones were used in quantities exceeding the median values. In total, 51 herds were included in the study.

**Figure 5.**
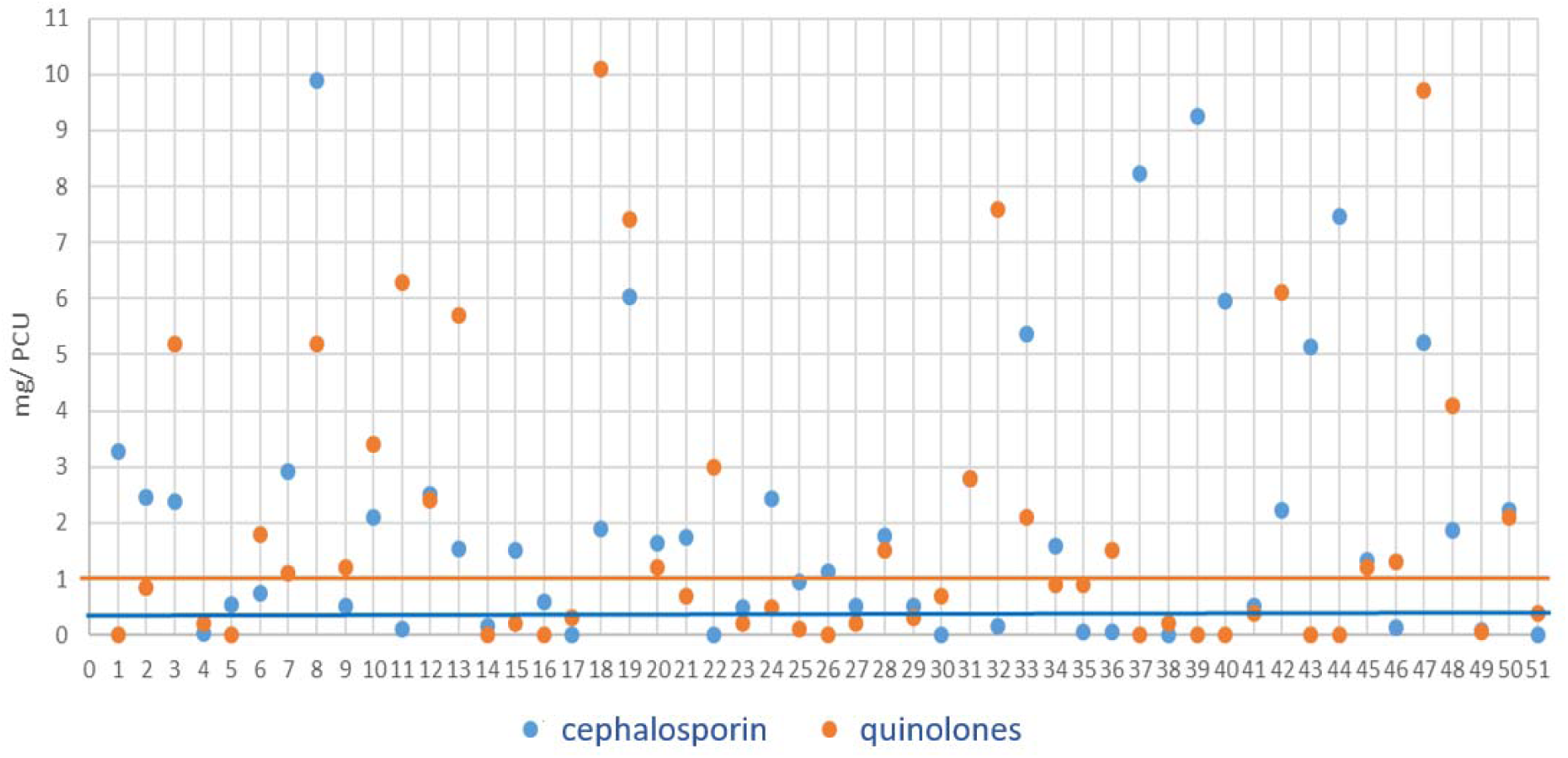
Amounts of cephalosporins and quinolones used in 51 study herds during 12 months. Lines represent the median values of the quantities used in the EU countries (ESVAC report 2020), which are 0.2 mg/PCU for cephalosporins and 1.1 mg/PCU for quinolones.

### 3.2. Use of Antibiotics in the Treatment of Swine Diseases

Data on the use of antibiotics for swine diseases were collected over a period of 12 months from 12 swine farms between 2018 and 2020. The selection of active ingredients varied between 2 and 6, with an average of four active ingredients used per farm. Oral antibiotics (doxycycline, tiamulin, colistin) were mainly used to treat piglet diarrhea. Injectable active ingredients (procaine benzylpenicillin, amoxicillin, tetracycline, enrofloxacin, ceftiofur, tiamulin) were used to treat joint, lung, and uterine infections in sows. Calculating the quantities of active ingredients was limited, because half of the farms did not register treatment data electronically. Consequently, it was not possible to perform an analysis of active ingredient quantities per mg/PCU (population correction unit). The distribution of active ingredients across farms is described in Figure 6.

**Figure 6.**
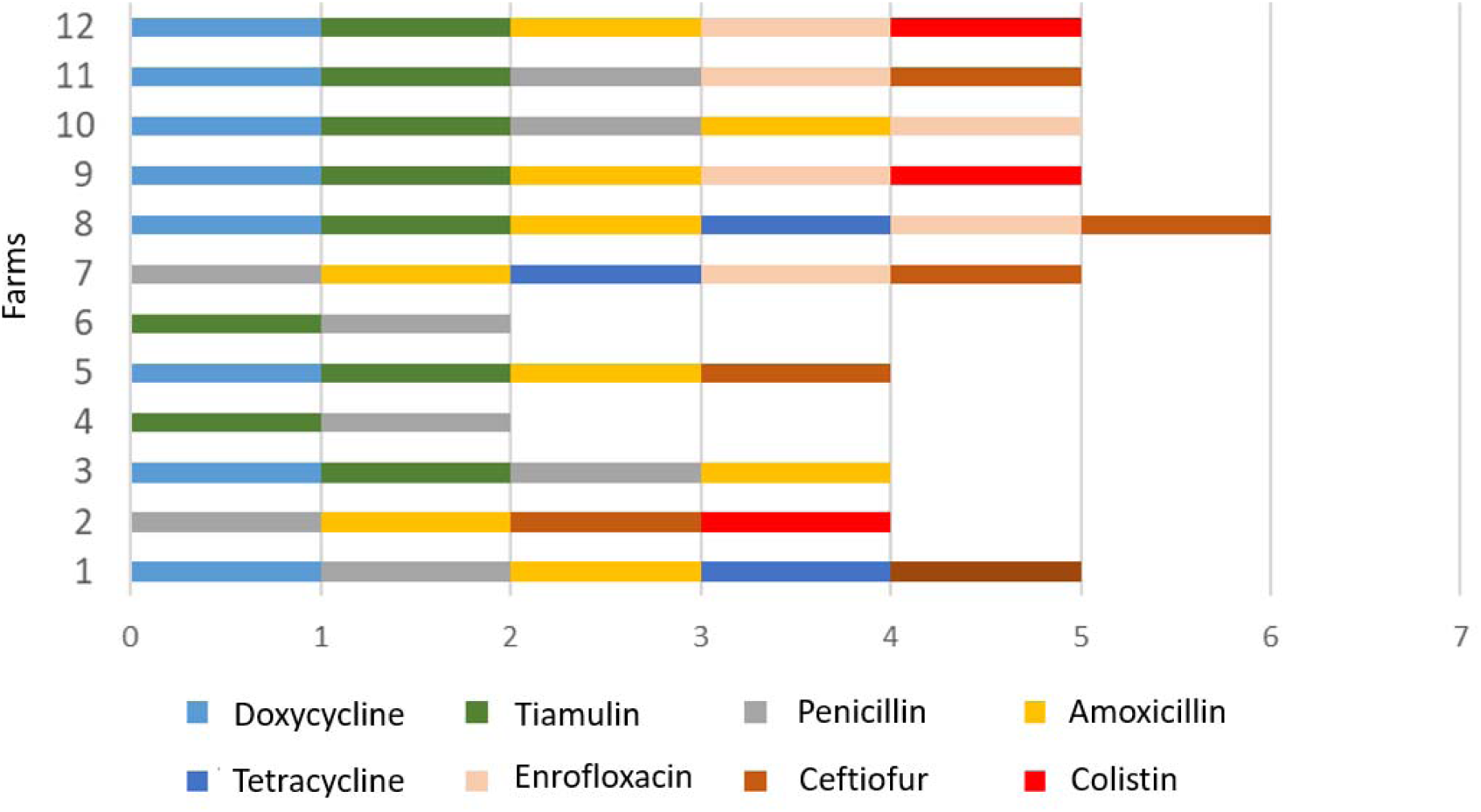
Number of active substances used by pig farms (n=12).

### 3.3. Impact of Cephalosporin Use on *E. coli* Antibiotic Resistance

A study on extended-spectrum beta-lactamase (ESBL)-producing *E. coli* was conducted on 46 cattle farms, and ESBL-positive *E. coli* isolates were found in 14 farms (30.4% of the studied farms). Farms with ESBL-positive *E. coli* had a 2.1 times higher use of 3rd and 4th generation cephalosporins (median 1.76 mg/PCU) compared to ESBL-negative farms (median 0.68 mg/PCU). Herds that used 3rd and 4th generation cephalosporins above the median of 1.5 mg/PCU had a 17% higher probability (p=0.091) of finding ESBL-positive *E. coli* microbes.

For pigs, ESBL/AmpC-type beta-lactamase-producing *E. coli* strains were detected in eight cases (66.7% of the studied swine). In two ESBL-positive pig herds, ceftiofur was used for intramuscular treatment in sows. In six farms where ESBL-positive *E. coli* microbes were found, cephalosporins had never been used for swine treatment. Given that there are approximately 40 pig farms in Estonia, conclusive conclusions cannot be drawn regarding the relationship between cephalosporin use and the presence of ESBL-positive *E. coli* due to the small sample size. The most common beta-lactamases were blaTEM-1B, blaTEM-52C, and blaCTXM-1.

## 4. Antibiotic Use in Human Medicine

Between 2017 and 2021, according to data from the Estonian Agency of Medicines, the most widely used antibiotic class in human medicine in Estonia was the beta-lactams. They were followed by the macrolides, lincosamides, and streptogramins (Figure 7). The most sold active ingredients in 2021 were amoxicillin/clavulanic acid (2.15 DDD/1000 patients/day), doxycycline (1.33 DDD/1000 patients/day), and amoxicillin (1.29 DDD/1000 patients/day). The most sold active ingredients are listed in Table 3.

**Figure 7.**
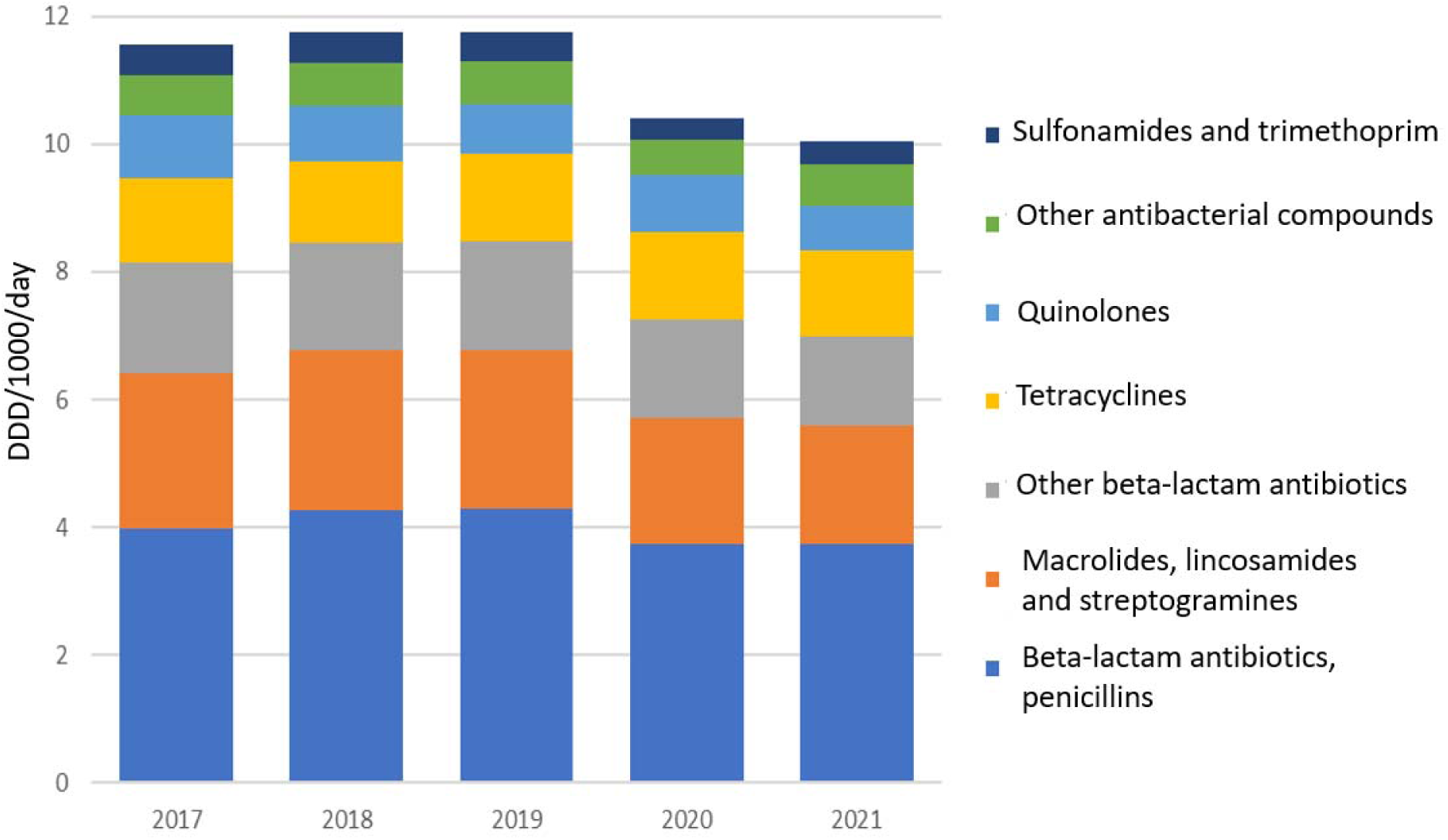
Antibiotic use in Estonia between 2017 and 2021, by ATC groups (defined daily dose per 1000 inhabitants per day). Source: Estonian State Agency of Medicines.

**Table 3.** Sales of antibiotics for human medicine to general and hospital pharmacies, as well as other institutions in Estonia for the years 2017-2021 (source: Estonian State Agency of Medicines).

| Active substance(s) | DDD/1000/day |  |  |  |  |
| --- | --- | --- | --- | --- | --- |
|  | Year 2017 | 2018 | 2019 | 2020 | 2021 |
| amoxicillin + clavulanic acid | 1.84 | 2.13 | 2.18 | 2.06 | 2.15 |
| doxycycline | 1.32 | 1.28 | 1.38 | 1.39 | 1.33 |
| amoxicillin | 1.64 | 1.64 | 1.68 | 1.31 | 1.29 |
| clarithromycin | 1.69 | 1.75 | 1.73 | 1.28 | 1.17 |
| cefuroxime | 1.30 | 1.26 | 1.26 | 1.12 | 1.01 |
| nitrofurantoin | 0.57 | 0.58 | 0.58 | 0.47 | 0.57 |
| ciprofloxacin | 0.68 | 0.67 | 0.59 | 0.62 | 0.54 |
| azithromycin | 0.55 | 0.56 | 0.58 | 0.52 | 0.53 |

### 4.1. Antibiotic Use in Hospitals

In 2016, a point prevalence survey was conducted in 23 Estonian acute care hospitals following a unified ECDC protocol. All regional and central hospitals were represented. Of the patients admitted on the survey day, 25.1% (1059 patients) received at least one antimicrobial drug. The statistical confidence interval (CI 95%) for this 25.1% is within the range of 23.8% to 26.4%. Both the prevalence of antimicrobial treatment (25.1% vs. 30.5%) and the use of drugs in defined daily doses per 100 patients (36 vs. 46) were lower in Estonia than the European average. Similar to other European countries, antimicrobial treatment was mostly used for the treatment of infections. The most common sources of infection (lower respiratory tract, urinary tract, abdominal cavity) did not differ significantly from the main sources of infection in other countries.

Based on data from the use of antibiotics in Estonian hospitals in 2019, collected from hospital pharmacies or statistical departments, the average total antibiotic usage for the 19 hospitals participating in the study was 51.5 defined daily doses (DDD) per 100 patient days (range 15-102 DDD/100 BD).

Although the use of antimicrobial treatment in Estonian hospitals is lower compared to many other European countries, there are several areas of concern.

#### 4.1.1. Variations in antibiotic consumption and selection in hospitals

In regional hospitals (the two largest clinical centers: North Estonia Regional Hospital and Tartu University Hospital), total consumption of antibiotics was similar, but in central hospitals, there was a significant variation. For example, Ida-Viru Central Hospital’ s total consumption was twice as high (82 DDD/100 BD) as Pärnu Hospital (41 DDD/100 BD). In general and local hospitals, there were six hospitals with higher usage than the Estonian average, with Rakvere Hospital having the highest usage (see Figure 8). Focusing to hospitals where usage is higher or significantly different from the Estonian average is needed. To address this, a better system for continuous monitoring of antibiotic usage is required.

**Figure 8.**
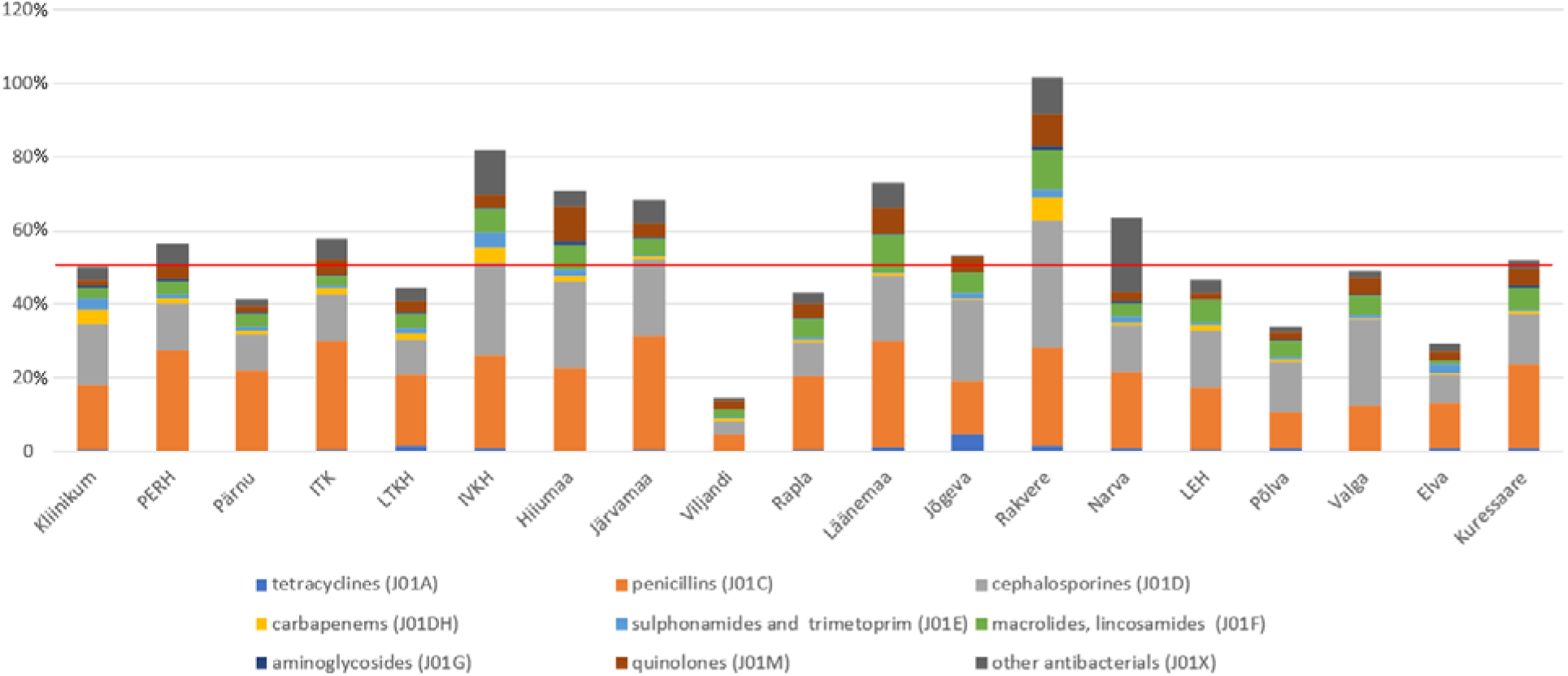
Antibiotic total usage in Estonian hospitals in defined daily doses per 100 patient days in 2019. Sorted by types of hospitals (regional hospitals: Tartu University Hospital (Kliinikum), North Estonia Regional Hospital (PERH); central hospitals: East-Viru Central Hospital (IVKH), East-Tallinn Central Hospital (IKH), West-Tallinn Central Hospital (LTKH), Pärnu Hospital; general and local hospitals: Rakvere Hospital, Läänemaa Hospital, Hiiumaa Hospital, Järvamaa Hospital, Narva Hospital, Jõgeva Hospital, Kuressaare Hospital, Valga Hospital, Southern-Estonia Hospital (LEH), Rapla Hospital, Põlva Hospital, Elva Hospital, Viljandi Hospital), and by total usage. The red line represents the average usage in Estonian hospitals (51.5 defined daily doses per 100 BD).

In the comparison of antibiotic consumption in departments with similar patient profiles, there were both similarities and significant differences. For example, in the Tartu University Hospital and North Estonia Regional Hospital, antibiotic usage in intensive care units varied little and the proportions of drug groups used were similar. However, in intensive care units of central hospitals total consumption varied more. For instance, the total consumption in Ida-Viru Central Hospital’s intensive care unit was half as much (81 DDD/100 BD) as that of Ida-Tallinn Central Hospital (170 DDD/100 BD).

The total antibiotic usage in the neurology departments of the University of Tartu Hospital and North Estonia Regional Hospital was similar (34 and 36 DDD/100 BD, respectively). In central hospitals, the usage in the neurology department of Pärnu Hospital was roughly equivalent to that of regional hospitals (35 DDD/100 BD), but it was significantly higher (172 DDD/100 BD) in Ida-Tallinn Central Hospital.

Although departments with similar patient profiles in regional hospitals usually had similar usage, the total usage in the urology department of the University of Tartu Hospital was more than half less (83 DDD/100 BD) than in North Estonia Regional Hospital’s urology department (183 DDD/100 BD).

The use of drugs in nursing departments varied in total amount. Hospitals with low usage in nursing departments included Põlva and Pärnu (5.9 and 7.7 DDD/100 BD); hospitals with average usage included the Tartu University Hospital and Valga Hospital (both 13.6), as well as Viljandi Hospital (15.2 DDD/100 BD); hospitals with slightly higher usage included Järvamaa Hospital (20.6), North Estonia Regional Hospital (23.8), and Elva Hospital (29.8 DDD/100 BD).

In departments with similar patient profiles, the selection of active ingredients sometimes varied significantly (e.g., in urology or cardiology departments). To determine the reasons for these differences, a more detailed analysis is needed.

#### 4.1.2. The high proportion of antibiotics according to the WHO Access, Watch, Reserve (AWaRe) Classification

In a study of antibiotic use in hospitals, antibiotics were classified according to the ATC classification and the WHO AWaRe classification into three main groups: main use (Access), restricted use (Watch), and reserve antibiotics (Reserve). The classification aims to promote rational antibiotic use and, thereby, reduce the development of antimicrobial resistance. The WHO recommendation is that, by 2023, 60% of antibiotics used in human medicine should belong to the main use category (Klein et al. 2021).

In 2019, 47 different antibiotics were used in Estonian hospitals, of which 19 were classified as Acess according to the AWaRe classification, 21 belonged to the Watch, and 7 to Reserve group. According to the AWaRe classification, the proportion of main use antibiotics in central and regional hospitals varied from 45% to 65% of total use, while restricted use antibiotics were more commonly used in general and local hospitals. Only three out of thirteen general and local hospitals met the WHO goal of having 60% of total antibiotic use classified as Acess group antibiotics.

Among central and regional hospitals, the use of restricted antibiotics was lowest in East-Tallinn Central Hospital and Pärnu Hospital (34%), and highest in East-Viru Central Hospital (54%) (Figure 9).

**Figure 9.**
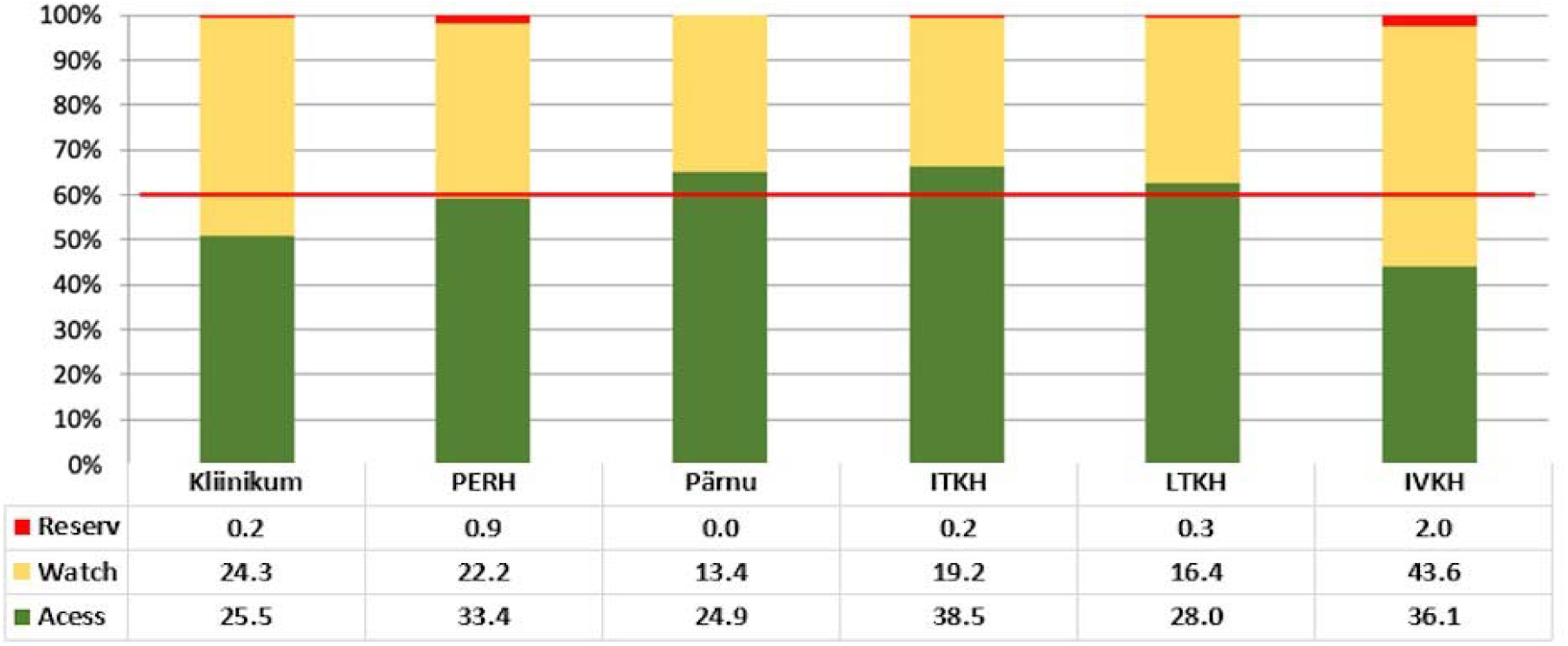
Share of antibiotics of the WHO Access, Watch, Reserve classification used in 2019 from the total use of antibiotics defined in daily doses per 100 BD. Use in regional and central hospitals. The red line marks the WHO target of at least 60% of the total use of antibiotics being from the Access group. Kliinikum - Tartu University Hospital; PERH - Northern Estonia Regional Hospital; ITKH - East-Tallinn Central Hospital; LTKH - Western Tallinn Central Hospital; IVKH - East-Viru Central Hospital.

Among general and local hospitals, Valga Hospital had the highest proportion of restricted group antibiotics (70%), followed by Põlva Hospital (62%) and Viljandi Hospital (61%) (Figure 10).

**Figure 10.**
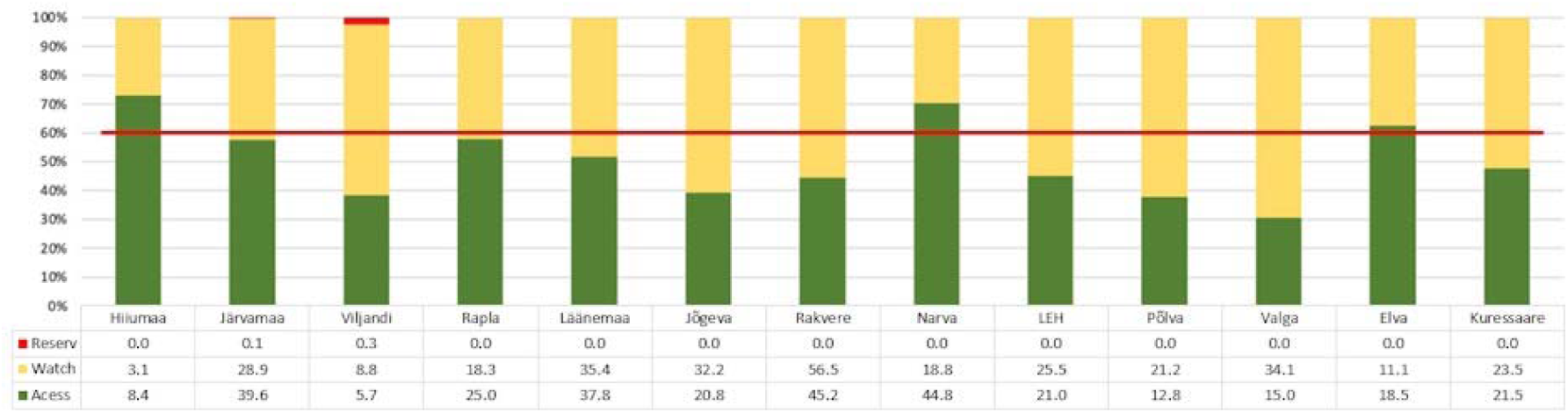
Share of antibiotics of the WHO Access, Watch, Reserve classification used in 2019 from the total use of antibiotics defined in daily doses per 100 BD. Use in general and local hospitals. The red line marks the WHO target of at least 60% of the total use of antibiotics being from the Access group. LEH - Southern Estonia Hospital; other hospitals are named according to the town or county of location.

The use of reserve antibiotics in Estonian hospitals was low, accounting for 0.5-1.6% in regional hospitals and 0.01-2.3% in general and local hospitals.

The most frequently used restricted group antibiotic in all hospitals was cefuroxime, with its proportion of total antibiotic use ranging from 6.5% in Kuressaare Hospital to 47% in Valga Hospital. Cefuroxime extensive use was confirmed in a point prevalence study in hospitals, which showed that cefuroxime ranked second in the treatment of community-acquired lower respiratory tract infections in Estonia and third in uncomplicated urinary tract infections. Surprisingly, fluoroquinolones, which belong to the AWaRe restricted group, were also frequently prescribed, primarily for urinary tract and, surprisingly, surgical site infections. The European Medicines Agency has issued a warning regarding the serious and potentially long-lasting side effects associated with fluoroquinolone use (State Agency of Medicines 2018). Therefore, monitoring the trend of fluoroquinolone use in hospitals is essential. Even in the AWaRe classification, fluoroquinolones are not recommended as first-line treatment.

It is necessary to analyze the reasons for the use of restricted antibiotics. It is crucial to determine whether it is based on existing guidelines or due to their absence, and how many of these antibiotics could be replaced by main use antibiotics in treatment guidelines. Furthermore, hospital doctors need training since the AWaRe classification is new and requires introduction. The AWaRe classification provides infection control doctors in hospitals with a novel opportunity to monitor antibiotic use, set goals, and make plans to promote more rational antibiotic use. The national goal could be to have 60% of hospital antibiotic use classified as main use category antibiotics. Compliance with the AWaRe classification could be included in the Health Insurance Fund’s quality indicators.

#### 4.1.3. Lack of Antimicrobial Treatment Guidelines and Infection Control Doctors in Hospitals

A point prevalence study conducted in 2016 revealed that 43.5% of Estonian hospitals lacked antimicrobial treatment guidelines, and 75% of general, local, and specialized hospitals had no antimicrobial treatment consultant, usually an infection control doctor. According to the 2019 hospital data report, nine of the hospitals participating in the study lacked an infection control doctor, who, according to legislation (https://www.riigiteataja.ee/akt/13253198), must be qualified in infectious diseases. The development of antibiotic use policy is one of the tasks of an infection control doctor. Adhering to guidelines can shorten the duration of antimicrobial treatment and hospital stay and reduce patient mortality (Versporten et al. 2018). A systematic review and meta-analysis showed that guideline-based antimicrobial treatment reduced patient mortality by 35% (Schuts et al. 2016). An infection control doctor’s tasks should also include limiting the use of drugs in the restricted group of the AWaRe classification, thus promoting more rational antimicrobial treatment (Pollack et al. 2016). An infection control doctor can also contribute to better treatment outcomes in various infectious diseases (Bai et al. 2015, Mejia-Chew et al. 2019).

Infection control doctors can also help with appropriate perioperative prophylactic antimicrobial treatment, as recommended by the WHO. In the prevalence survey conducted within the framework of this project, it was revealed that in Estonian hospitals, perioperative prophylaxis is performed for more than one day in a quarter of patients, which is not in accordance with the treatment guidelines. In most cases, so-called extended prophylaxis was performed in oncological, orthopedic and urological operations, and the problem is most pronounced in regional, general, local and special hospitals in Estonia. 10% of all antimicrobials administered on the study day were used for medical prophylaxis. Surprisingly, 61.5% of the drugs used for prophylaxis were registered in general, local and special hospitals, where there should not be more immunocompromised patients than in regional or central hospitals. This result, as well as the use of 25 different active substances (of which only 6.2% was trimethoprim/sulfamethoxazole), points to the fact that the indication for prescribing these drugs was unclear, and therefore the use of antimicrobial therapy was not justified.

A significant deficiency in the field of guidelines was also indirectly indicated by the 2019 hospital data report, according to which there are large variations in the antibiotic use of both hospitals and departments of the same type. At the national level, it is necessary to pay attention to the availability of antibiotic treatment guidelines and training in hospitals and the presence of an infection control doctor in all hospitals.

#### 4.1.4. Preference for intravenous drugs over oral ones

The results of the current prevalence survey showed that 78.7% of all drugs were administered intravenously and only 12.5% of drugs administered on the study day had been changed from intravenous to oral form. Many intravenous drugs are more expensive than oral drugs, and the costs of administration, preparation and storage are added (Mertz et al. 2009). Oral treatment has several advantages: no risk of catheter infections related to antimicrobial therapy, the more comfortable for patient, reduction of treatment costs, lower nurses workload, shorter length of hospital stays (Davis et al. 2005, Barlam et al. 2016). One method to optimize the use of antimicrobial therapy is to encourage switching from intravenous to oral treatment, following the relevant instructions.

#### 4.1.5. Inadequate documentation of antimicrobial therapy at the medical records

One of the indicators of the quality of antimicrobial therapy is the rationale for the use of antimicrobial therapy documentation in the medical records (Pulcini et al. 2019). Documenting the justification for antimicrobials use allows all healthcare workers dealing with the patient to understand the purpose of the treatment, methods of administration (intravenous or oral), duration and need for de-escalation. In the current prevalence survey, there was documented rationale for antimicrobial treatment in 84.2% of cases. Also, 48-72 h after the start of antimicrobial treatment, it should be assessed whether the treatment is appropriate and whether it is possible to switch from a wider spectrum of action to a narrower one. Such a method allows to avoid unnecessary long-term antimicrobial therapy, reduce drug-related side effects and *Clostridioides difficile* infection (Pollack et al. 2016).

#### 4.1.6. Quality of antibiotic use data

Antibiotic use in hospitals has been previously analyzed mainly on a hospital-by-hospital basis and no national comparison has been made so far. There is no national system of assesing the antibiotic use in hospitals available for data collection and analysis.

One of the goals of the study was to determine how data on antibiotic use is collected and analyzed in hospitals. Based on the results of the current analysis the aim was to evaluate the possibilities for harmonizing the collection of antibiotic use data in hospitals. The collected data are inconsistent because of different format used by different hospitals and lack of experience of smaller hospitals when collecting and analysing the data of antibiotic use. The composition and structure of the data also depended from the pharmacy program or accounting program used by the respective hospital It also turned out that some hospitals do not analyze and report the data about antibiotic use for perioperative antibiotic prophylaxis or for the treatment of tuberculosis. Because manual data collection is laborious, inaccurate, and not sustainable, the collection of antibiotic use data should be automated in the future.

### 4.2. Outpatient use of antibiotics

Based on the reports of the European Center for Disease Prevention and Control (ECDC), Estonian outpatient use of antibiotics in the last 20 years has been relatively stable and one of the lowest in Europe. For example, in 2020, the total ambulatory antibiotic use in Estonia was 8.8 defined daily dose per 1000 inhabitants per day. In European Union / European Economic Area the corresponding average indicator was 15 (Figure 11) (ECDC, 2021).

**Figure 11.**
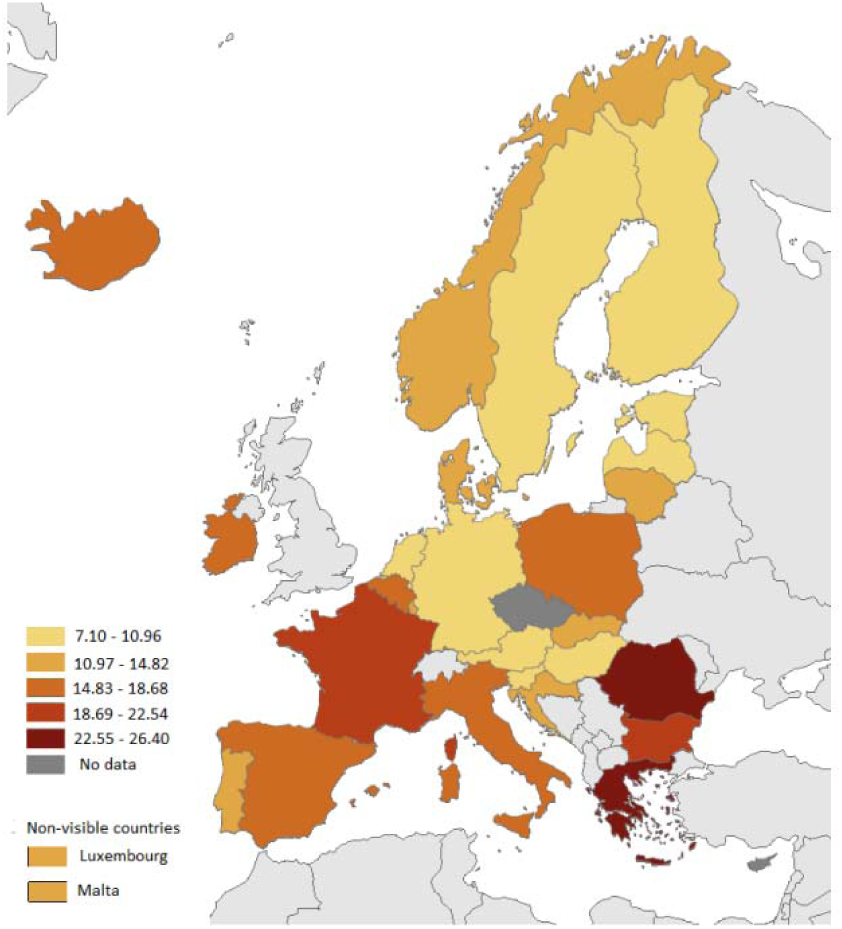
Total ambulatory use of systemic antibiotics (ATC classification group J01). defined in daily doses per 1000 inhabitants per day of the European Union / European Economic Area countries in 2020. (European Centre for Disese Prevention and Control)

A comparison of outpatient antibiotic use from 2018 showed that the general frequency of use of antimicrobials in Estonia is similar to Finland, Sweden, Latvia and Denmark (respectively 10.2; 13.2; 10.8; 11.4 and 13.7 DDD/1000 per day). However, there are significant differences in the drug groups used. For example, significantly less penicillin is used in Estonia, Latvia and Finland than in Sweden and Denmark. Cephalosporins are also used more in Finland and Estonia than in Sweden and Denmark. Tetracyclines are used the least in Estonia and the most in Finland. Estonia ranks first in the use of macrolides. Despite the low use of antibiotics, broad-spectrum antibiotics are a concern in Estonia as these antibiotics have increased in the proportion of total usage. We assessed ambulatory antibiotic use in Estonia in 2008-2018, based on ESAC-Net ambulatory antibiotic treatment quality indicators using the data provided by Estonian Agency of Medicines. A total of 12 indicators were analyzed, of which two indicators had changed significantly during 11 years. The data indicated an adverse change in the broad-spectrum and narrow-spectrum penicillins, cephalosporins and macrolide use ratio. In 2008 broad-spectrum penicillins, cephalosporins and macrolides were used 5.4 times more than narrow spectrum penicillins. In contrast, in 2018 the ratio was 16 times bigger. Ratio of the combination of penicillins with beta-lactamase inhibitors (ATC classification group J01CR) to the total use of all antibacterial substances (J01) also changed significantly. In 2008, J01CR subgroup use of the J01 group was 8.3%, and in 2018 the same figure was more than twice as high, i.e. 19.1%. Therefore, we also analyzed ambulatory antibiotic use in more detail using The Health Insurance Fund’s prescription drug database. We found that ambulatory antibiotic use in Estonia remained low in the years 2014-2018 and the number of antibiotic prescriptions per 1,000 inhabitants per year has rather a downward trend (Table 4). However, we identified several potential problem areas, dealing with which will help to make antibiotic use even more optimal.

**Table 4.** Total number of purchased antibiotic prescriptions (PAP) and number per 1000 inhabitants by year in the years 2014-2018.

| Year | PAP total number | PAP per 1000 inhabitants |
| --- | --- | --- |
| 2014 | 581 025 | 442.0 |
| 2015 | 575 166 | 437.5 |
| 2016 | 552 897 | 420.2 |
| 2017 | 548 923 | 416.7 |
| 2018 | 561 401 | 424.7 |
| Total | 2 819 412 | 428.2 |

#### 4.2.1. Prescribing antibiotics for diagnoses that are not indicated for antibacterial treatment

Antibiotics were most often prescribed for respiratory infections (43.5%). Of them in turn, 49.5% of the prescriptions (608,342 purchased antibiotic prescriptions) were prescribed with respiratory diagnoses for which the guideline for ambulatory antibiotic treatment of the Estonian Society of Infectious Diseases recommends antibiotic use should be avoided (acute respiratory infections, sinusitis, bronchitis), as the disease is mostly caused by a virus (Estonian Society of Infectious Diseases 2018). Thus, it should be possible to reduce current antibiotic use with respiratory indications alone by at least 20%.

#### 4.2.2. Widespread and steadily increasing use of broad-spectrum antibiotics

During the study period, the use of penicillins combined with a beta-lactamase inhibitor has continuously increased (APC 8.1%, CI 95% 7.9-8.3). The highest use was in the age group 0-9 years of life (Figure 12).

**Figure 12.**
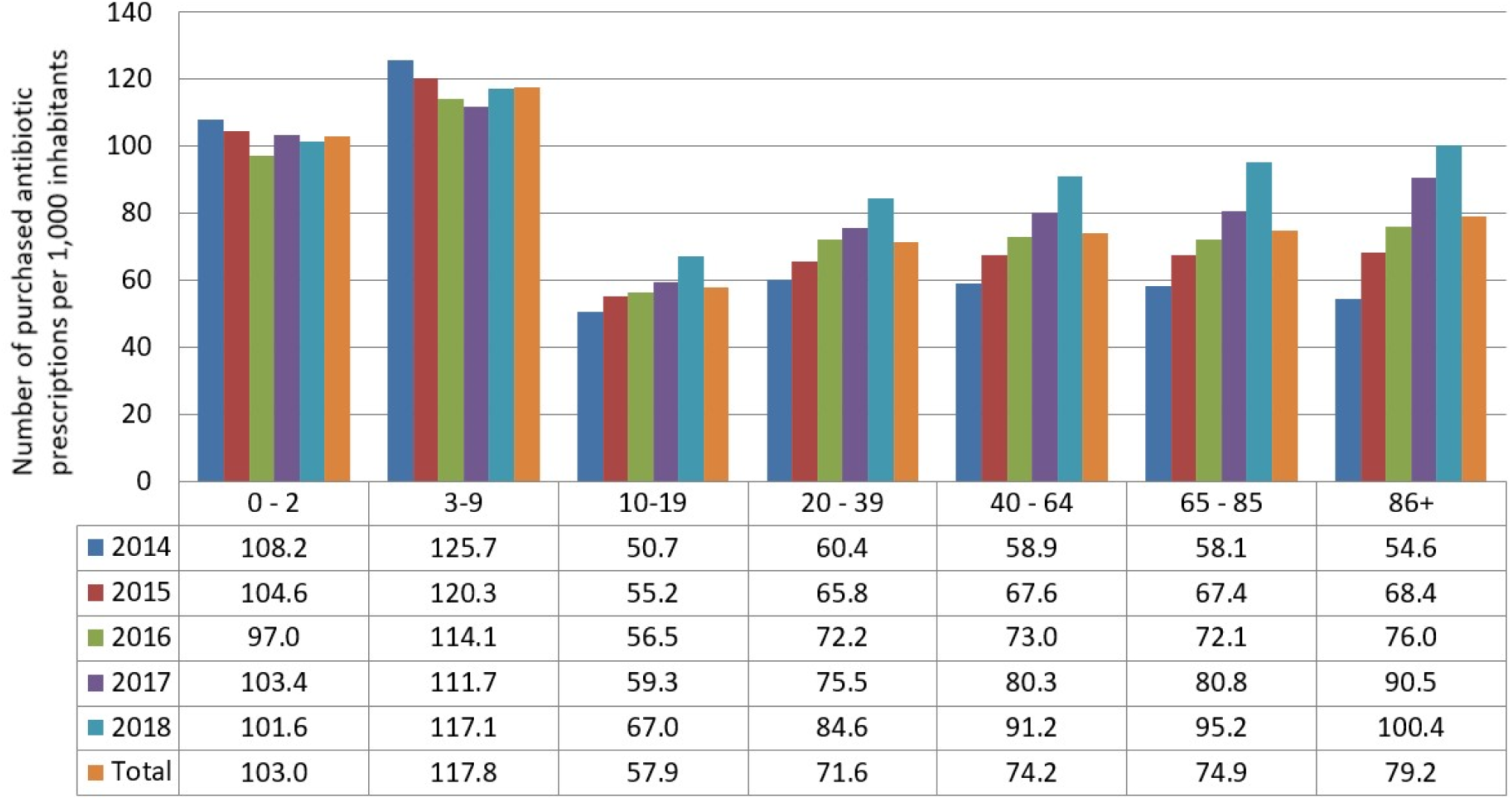
Penicillins combined with a beta-lactamase inhibitor. Number of purchased antibiotic prescriptions per 1000 inhabitants by age group.

The most common indications for which penicillin combinations with a beta-lactamase inhibitors were used are infections of the respiratory tract and teeth and jaws (Figure 13). Unfortunately, these usage patterns have an upward trend.

**Figure 13.**
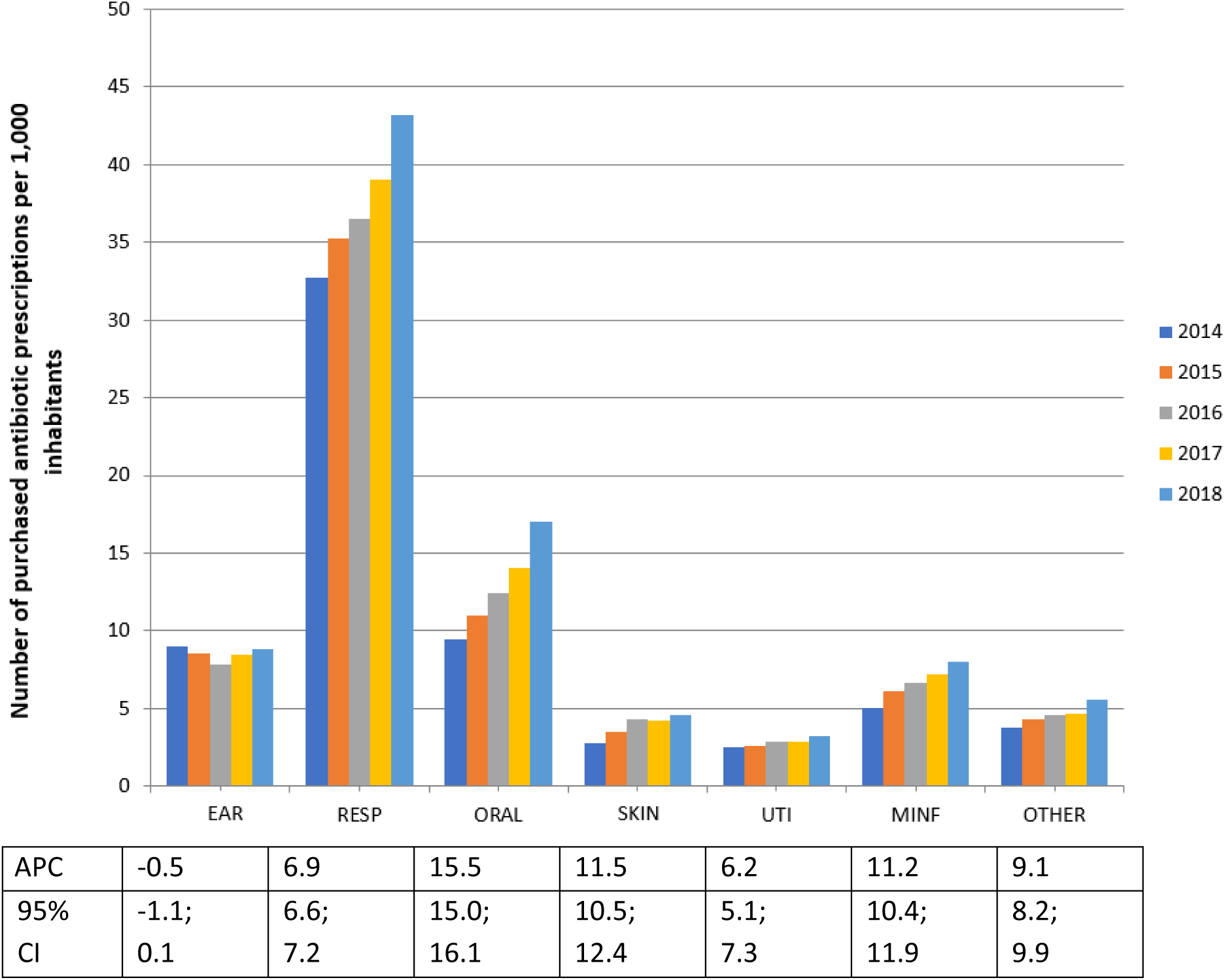
Penicillins combined with a beta-lactamase inhibitor. Number of purchased antibiotic prescriptions per 1,000 inhabitants. Distribution between diagnosis groups: EAR - ear infections; RESP - respiratory infections; ORAL - oral, infections of the salivary glands and jaws; SKIN – infections of the skin, subcutaneous tissue and mammary gland; UTI – urinary tract infections; MINF – non-infectious diagnoses; CI – confidence interval; APC – average percentage change per year.

The combinations of beta-lactams with inhibitors were prescribed for nearly one-third of tooth and jaw infection indications. There is no antibacterial treatment guide for oral and dental infections in Estonia. Based treatment instructions in other countries (e.g. Great Britain and the United States), broad-spectrum antibiotics (beta-lactamase inhibitor combined penicillins, 2nd generation cephalosporins, macrolides and lincosamides) should be avoided as a first choice (NICE 2020, Lockhart et al. 2019).

Although suspected respiratory viral infections are mostly treated with macrolides, the increase in usage of penicillins combined with a beta-lactamase inhibitor is a problem also for this diagnoses group.

#### 4.2.3. The selection of the antibiotic was not based on the recommendations of the treatment guidelines

Although the guidelines given in the guidelines of the Estonian Society of Infectious Diseases for the treatment of pneumonia recommendto use either amoxicillin or amoxicillin/clavulanic acid (dependent of the risk factors) (Estonian Society of Infectious Diseases 2018), in almost half of the cases (43.8%) macrolide was preferred (Figure 14).

**Figure 14.**
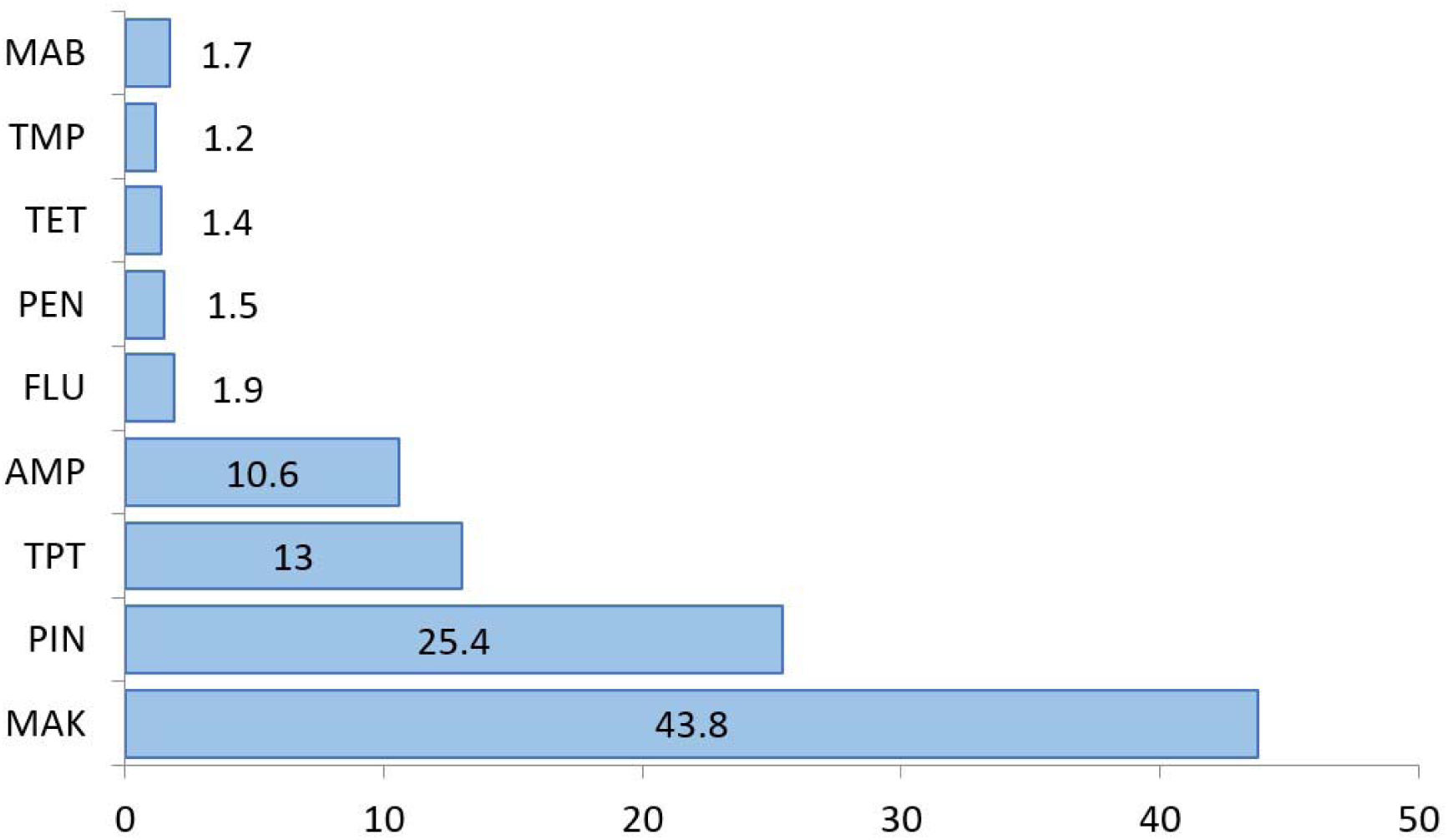
Antibiotic prescriptions purchased with pneumonia diagnosis in 2014-2018. MAB - other antibiotics; TMP – trimethoprim/sulfamethoxazole; TET – tetracyclines; PEN – penicillins; FLU - fluoroquinolones; AMP – aminopenicillins; TPT - 2nd generation cephalosporins; PIN – penicillins combined with a beta-lactamase inhibitor; MAK – macrolides.

For treatment of skin and soft tissue infections 39.6% of antibiotic choices followed the guidelines (34.2% had used 1st generation cephalosporin and 5.4% clindamycin). Unfortunately, in 31.4% of cases, an inappropriately broad spectrum antibiotic was used, namely penicillin combined with a beta-lactamase inhibitor (22.5% of cases) or 2. generation cephalosporin (8.9% of cases). At the same time, the penicillin-inhibitor group has been used during the study period with a constant upward trend.

Although the guideline recommends avoiding fluoroquinolones in the treatment of cystitis, in 29.8% of cases it is still prescribed to the patient (Figure 15). Regular auditing of antibiotic use, feedback and continuous training of doctors are expected to improve adherance to the guidelines.

**Figure 15.**
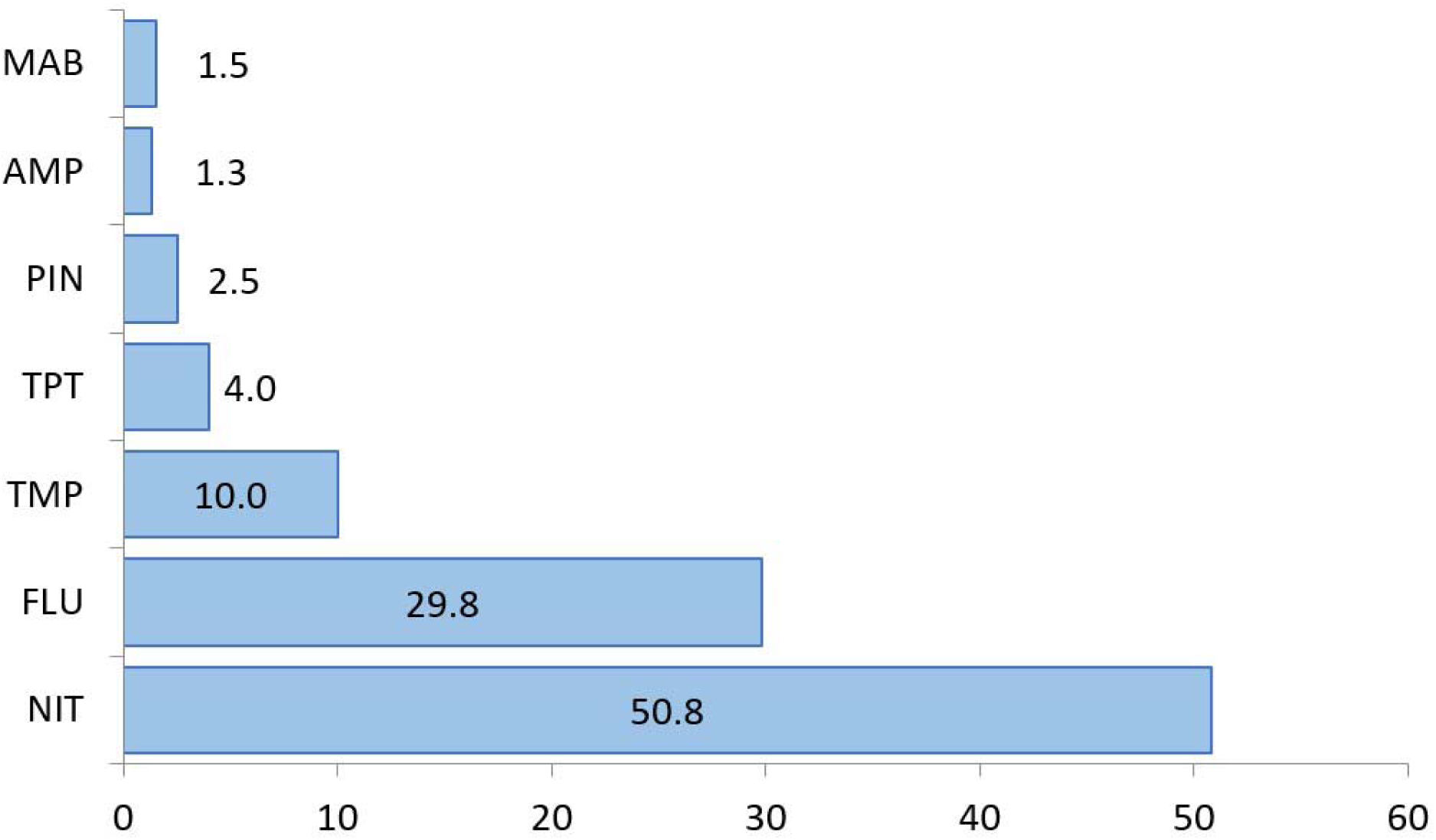
Antibiotic prescriptions purchased with cystitis diagnosis in 2014-2018. MAB - other antibiotics; AMP – aminopenicillins; PIN – penicillins combined with a beta-lactamase inhibitor; TPT - 2nd generation cephalosporins; TMP – trimethoprim/sulfamethoxazole; FLU - fluoroquinolones; NIT - Nitrofurantoin.

#### 4.2.4. Seasonal variations in antibiotic use

Variation in the seasonal use of antibiotics (increased use in the winter months as compared to the summer months) is an important ESAC-Net indicator of the quality of ambulatory use of antibiotics. In our study large seasonal differences emerged. Based on the prescription data, the difference between antibiotic prescriptions purchased during the highest period (1st quarter) and the lowest period (3rd quarter) is almost 1.5 times (Figure 16).

**Figure 16.**
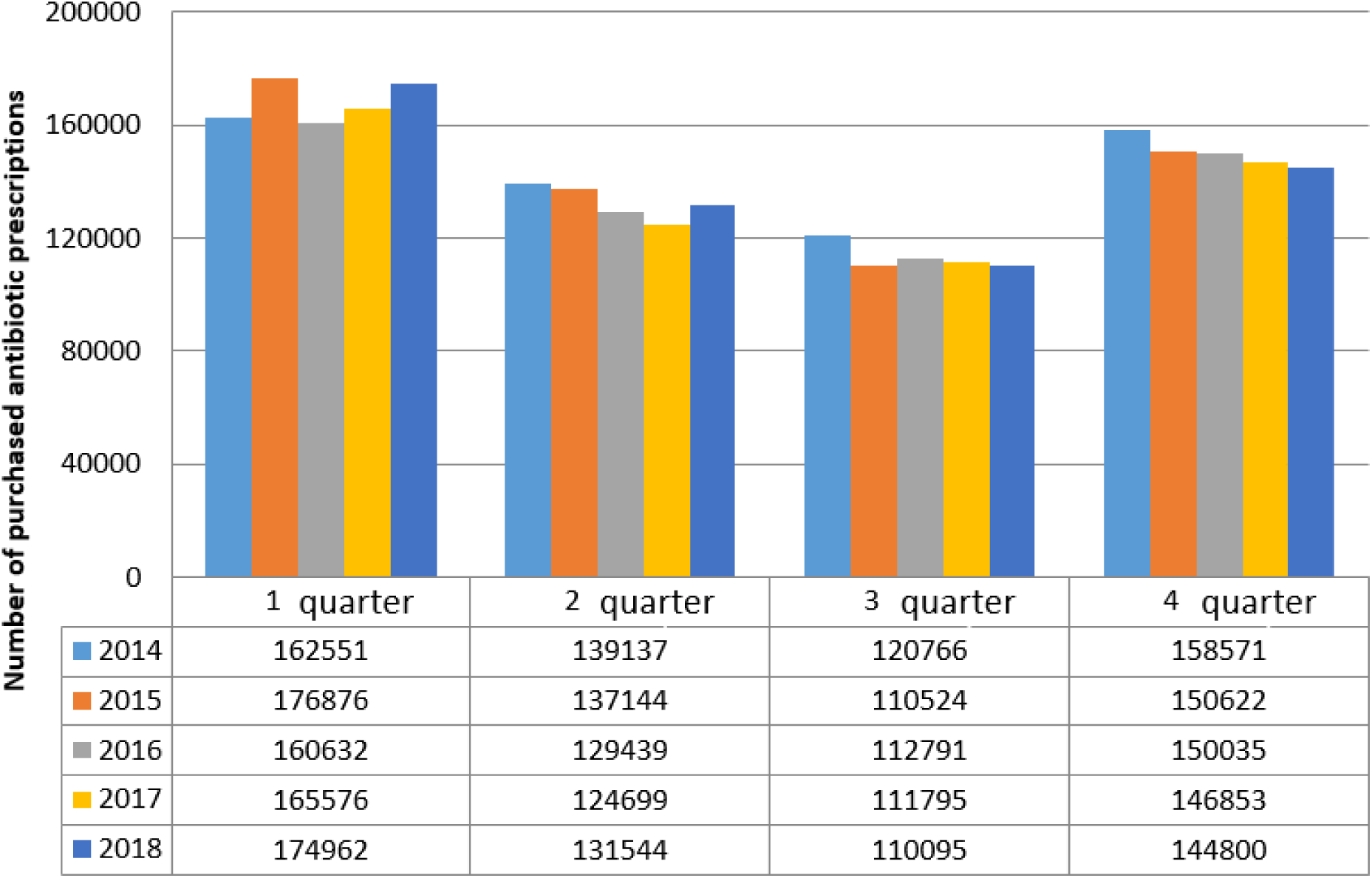
Distribution of the number of purchased antibiotic prescriptions between different quarters of the year.

This phenomenon is not unique to Estonia, but has also been described in numerous previous reports (Suda et al. 2014). It has been found that the seasonality of antibiotic prescribing is caused the increase frequency of respiratory infections in the winter months (Durkin et al. 2018, Suda et al. 2014). Because this coincides with the peak season of viral infections, then it can be assumed that part of the antibacterial treatment is also prescribed to treat viral infections during this period. However, compared to other Nordic countries, the the seasonal variability in Estonia is extremely high - in the winter months, the estimated overuse in Estonia is 34%. In Denmark, this figure is 8.6 and in Finland 16.9%. An audit would help to determine whether antibiotics have been prescribed for the right indications.

#### 4.2.5. High antibiotic use among children

The highest number of purchased antibiotic prescriptions per 1000 inhabitants was in age groups 0-2 and 3-9 (608.9 and 625.5, respectively). The reports of other countries also describe the largest antibiotic use in the age group up to 3 years that is connected with frequent respiratory tract infections. Equivalent number of purchased antibiotic prescriptions per 1000 population in age group 3-9 is, however, rather exceptional (Hicks et al. 2015, King et al. 2020, SVA 2018). However, it is known that in this age group there are also more respiratory viral infections, which is why an audit is necessary to clarify whether antibiotics are prescribed for the correct indications in this age group (Pattemore 2008).

#### 4.2.6. Large regional differences in antibiotic use

There are large county differences in the use of antibiotics. When in Tartu County the number of purchased antibiotic prescriptions per 1000 inhabitants was 513.6, then with the lowest use in the county - Raplamaa - the corresponding indicator was 269.7. A similar regional difference also characterized broad-spectrum antibiotics (penicillins combined with a beta-lactamase inhibitor) use (Figure 17). The analysis made does not allow to explain the given differences and therefore it is necessary conducting a regional audit.

**Figure 17.**
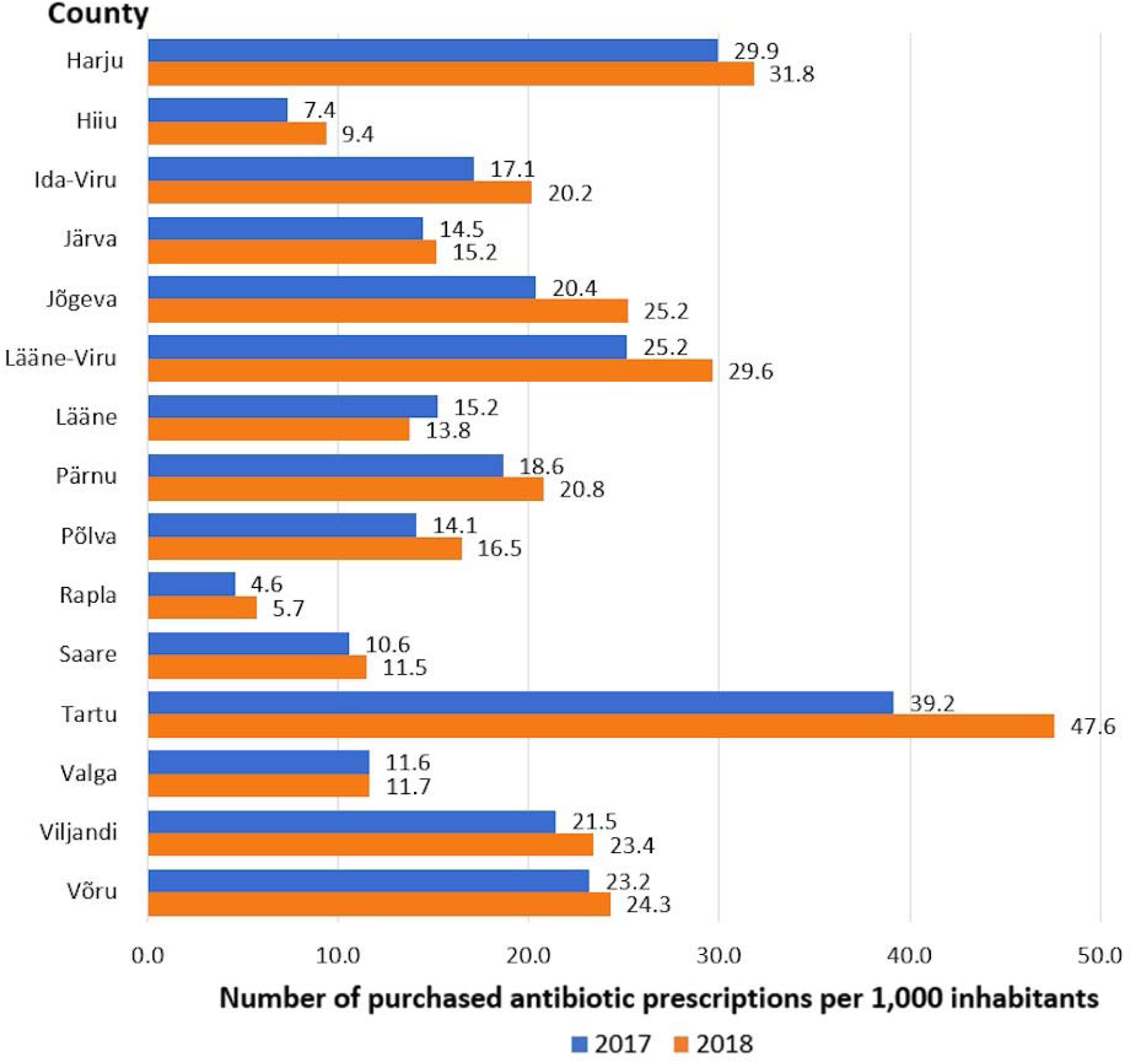
Purchased penicillins combined with a beta-lactamase inhibitor. Number of antibiotic prescriptions per 1,000 inhabitants by county in 2017 - 2018.

#### 4.2.7. Documentation errors

Marking the doctor’s specialty and healthcare service area on the prescription has improved year by year. However, there are significant deficiencies in noting the diagnosis. In 7% of the prescriptions, antibacterial treatment was prescribed for non-infectious cause.

Of the purchased antibiotic prescriptions, 4.1% (n=116,199) were for an unspecified urinary tract infection. The use of this diagnosis has increased year by year. However, the inaccuracy of the diagnosis does not allow to give clear recommendations for the choice of antibiotics. There are also differences in prescription practices, some physicians prescribing nitrofurantoin as primary choice and some fluoroquinolones.

### 4.3. Prevention of infections caused by resistant microbes

Analysing bloodstream infection caused by ESBL-positive *E. coli* and *K. pneumoniae* in hospital patients, we found risk factors that may affect the treatment of patients and antibiotic prescribing practices.

4.3.1. Colonization or infection with an ESBL-positive strain of *E. coli* or *K. pneumoniae* during the previous year

Observations on this risk factor are similar to the results of several other studies where previous carriage or infection with ESBL-positive enterobacteria is an important risk factor for the development of resistant bloodstream infection (van Aken et al. 2014, Freeman et al. 2012, Ben-Ami et al. 2009). In Sweden a population-based study found that people with isolated ESBL-positive enterobacteria in their urine or feces have the risk of subsequent bloodstream infection 61 and 32 times higher, respectively, when compared to people who have not had this pathogen before (Isendahl et al. 2019). Choice of adequate treatment of a bloodstream infection in the hospital is facilitated by displaying data about ESBL-positive E. coli or K. pneumoniae on the first page of the digital health record for at least up to 1 year after the isolation of the given strain from the patient.

#### 4.3.2. Antibiotic treatment within the previous 6 months

Antibiotic use has been implicated as a risk factor for ESBL-positive enterobacteriaceae in many studies (Tumbarello et al. 2006). In our study, antibiotic treatment within the previous 6 months was found to be an important risk factor for ESBL-positive *E. coli* and *K. pneumoniae* bloodstream infection. Therefore it is also important to collect and analyze consistently resistance data of ambulatory microbiological cultures. Based on this, adequate empiric antibacterial therapy can be recommended in case of suspicion of resistant strains (e.g. previous colonization known) and thereby risk of bloodstream infection reduced. Consistent analysis of antibiotic use data is important both in outpatient and hospital settings. This will assist in avoiding overuse of cephalosporins and thereby preventing the development of resistance.

#### 4.3.3. Staying in a nursing home during the previous 6 months (*E. coli*); hospital and healthcare acquired infection (*K. pneumoniae*)

Our study confirmed that the risk factor for ESBL-positive *K. pneumoniae* is hospital and healthcare-acquired infection. In our study, K. pneumoniae ST307 dominated. This ST has been associated with hospital outbreaks previously (Peirano et al. 2020). At the same time, sequencing data of our study strains (high divergence of nuclear genomes) exclude clonal spread. The reason for the lack of clonal spread may be the contact isolation requirements used in our hospitals for patients with ESBL-positive *K. pneumoniae* infection or colonization. Therefore, it is necessary to continue to treat such patients in hospital contact isolation zone as also recommended by the European Society of Clinical Microbiology and Infectious Diseases guide (Tacconelli et al. 2012). In the present study, one risk factors for acquiring ESBL-positive *E. coli* was previous stay in a nursing home. In a recent systematic review and meta-analysis, Bezabih et al observed an increase in carriage of ESBL-positive *E. coli* outside the hospital (Bezabih et al. 2021). During recent years, studies have paid more attention to the role of nursing homes in the spread of multi-resistant microorganisms (Vink et al. 2020). A systematic review and meta-analysis by Flokas et al described that the prevalence of ESBL-positive enterobacteria in nursing homes is 18% in Europe and 13% in North America (Flokas et al. 2017). Duval et al found that carriage of ESBL positive enterobacteria among people entering nursing homes and the rate of acquisition of this pathogen there are more frequent than in intensive care units (Duval et al. 2018). These studies suggest that nursing homes may have a significantly higher load of ESBL-positive enterobacteria as compared to the hospitals. In addition, mathematical models in a study in Netherlands showed that nursing homes might play a central role in the emergence and persistence of nation wide outbreaks of multi-resistant Gram-negative microorganisms (van den Dool et al. 2016). Therefore, it is important to prevent the spread of resistant microorganisms also in nursing homes. It is not possible to isolate clients in nursing homes, but care should be taken to follow standard infection control requirements (e.g. hand hygiene, correct surface cleaning and disinfection, use of personal protective equipment). It is necessary to deal with nursing home infection control topics at the national level (guidelines, training, networking with infection control services of central hospitals).

#### 4.3.4. An invasive device (bladder catheter, epicystostomy or nephrostomy) in the urinary tract for 48 hours before taking a positive blood culture (*K. pneumoniae*)

A risk factor for ESBL-positive *K. pneumoniae* in the present study was an invasive device in the urinary tract (bladder catheter, epicystostomy, or nephrostomy) within 48 hours before taking a positive blood culture. A Swedish population-based study found that a person previously colonized with ESBL-positive enterobacteria and having accompanying urological problem has hazard ratio for developing ESBL-positive bloodstream infection 3.4 times higher than control group (95% CI 2.47–4.69) enterobacteria the (Isendahl et al. 2019). In a study by Duffy et al., it was observed that a quarter of patients with health-care acquired ESBL-producing enterobacterial infections had a bladder catheter in the previous two days before giving positive microbiological test. It was concluded that ensuring proper care of the catheter (including removal of unnecessary catheters) can help to reduce infections by ESBL-positive enterobacteria(Duffy et al. 2022). Urinary tract infections, especially bladder catheter-related infections, are one of the most frequent healthcare-acquired infections (Cassini et al. 2016). The invasive agent in the urinary tract promotes the colonization of the urinary tract with bacteria and thus increases risk of infection (Bonten et al. 2021). In a systematic review on *E. coli* bacteremia in high income countries, Bonten et al observed that more than half of the cases were related with urinary tract infection; the risk of bloodstream infection increased 3 or 6 times when the hospitalized patient had either a bladder catheter or an epicystostomy, respectively. They concluded that to reduce *E. coli* bacteremia, urinary tract infections should be prevented (Bonten et al. 2021). To prevent bladder catheter related urinary tract infections, the knowledge of healthcare workers and nursing home staff should be increased. Bladder catheters are often used for the wrong indication (Laan et al 2020). Insertion of a bladder catheter for the correct indication and reduction of the duration of catheterization are related with decrease in infections (Fakih et al. 2012). The best remedies for prevention are the presence of relevant guides and supplies for inserting and maintaining bladder catheters in hospitals and nursing homes; bladder catheter removal reminders, surveillance of bladder catheter related urinary tract infections and training of staff (Lo et al. 2014, Hooton et al. 2010, Laan et al. 2020).

## 5. Antibiotic residues in the environment

Antibiotic residues, like other drug residues, can reach the environment. This can happen, for example, when manure or composted sewage sludge is used as fertilizer. Antibiotic residues can also enter into the water cycle through sewage treatment plants. In addition to promoting the emergence and spread of antibiotic resistance, antibiotic residues released into the environment can damage the entire ecosystem.

The current study investigated soil samples taken from fields near 5 farms and water samples from drainage ditches adjacent to the fields. Three farms are located in Tartu County, one in Jõgeva County and one in Lääne-Viru County. There are no ditches or other water bodies in the vicinity of one of the farms as it is in a karst area. All fields received regular application of liquid manure from the farm. Samples were collected from January 2020 to August 2021, at least once in each season. In one of the five farms, additional samples were simultaneously taken from farm animals (pig faeces) and liquid manure.

The development of antibiotic residue analysis methodologies was based on antibiotic usage and environmental stability data. The determination limits of the analytical methodologies were set according to the EMEA/CVMP recommendations of 100 ng/l for water samples, 10 μg/kg for soil and 100 μg/kg for manure samples. Since there is no official detection limit for sewage compost, the detection limit for manure was used.

Determining antibiotic residues turned out to be difficult for technical reasons. In the case of water samples, all antibiotic residues of interest were determined successfully (amoxicillin, azithromycin, benzylpenicillin (penicillin G), cefuroxime, ciprofloxacin, clarithromycin, doxycycline, enrofloxacin, florfenicol, marbofloxacin, norfloxacin, ofloxacin, oxytetracycline, sulfadimethoxine, sulfamethoxazole, tiamulin); 3 non-antibiotic pharmaceuticals were also detected (diclofenac, carbamazepine, triclosan). In the case of soil and sewage sludge samples, it was only possible to determine fluoroquinolones and tetracyclines.

We detected fluoroquinolones and tetracyclines in liquid manure and uncomposted sewage sludge in comparable concentrations. We found enrofloxacin in the liquid manure of the farm under observation 10 μg/kg, oxytetracycline 30 to 124 μg/kg, and doxycycline 30 to 480 μg/kg. In sewage sludge ciprofloxacin was in concentrations between 5 and 74 μg/kg and ofloxacin 23 μg/kg. Composting reduces the content of drug residues; the efficiency of the process depends on the technology used. Based on the literature, fluoroquinolones are known to be stable in the environment (Thiele-Bruhn 2003). However, these compounds can degrade during composting of sewage sludge. Initially fluoroquinalones can be in liquid manure and raw sewage sludge in a comparable quantity. However, during composting of the sewage sludge the concentration drops up to ten times and less residues reach the field as compared to liquid manure.

Antibiotics were detected in the surface water (ditch next to the field) on only one field of the five fields under study. Amoxicillin was detected at a concentration of 190 ng/l. This antibiotic had been used in the particular farm located close to the field. Thus, in this study we did not establish that antibiotics in liquid manure would quickly reach surface water if applied onto the field. At the same time the question needs more detailed research, as the amount of antibiotic use in the farm, liquid manure application time, rainfall and soil properties must be considered.

We also monitored the situation in aquaculture. A total of 5 different fish farms were studied in different parts of Estonia. The collection of samples was carried out in the early spring of 2020 and again at the end of the summer. Expected misuse of antibiotics could take place during spring-summer, when a new generation of fish is introduced. We did not observe antibiotic residues in the aquaculture effluent. We also did not detect elevated levels of antibiotic-resistant bacteria. At the same time, we recorded that the farms under observation did not use antibiotics during the study period. Thus, cases where antibiotics are actually used in aquaculture may cause concern.

In addition to antibiotic residues, we determined in surface water concentrations of some non-antibiotic pharmaceuticals that accumulate in the environment. The high levels of diclofenac and carbamazepine in surface water are of concern as we found these substances from several samples (Table 5). These are medicines for only human use, therefore they move into the environment through sewage treatment plants and/or sewage sludge compost. It also might indicate the potential risk that people do not handle unused medicines correctly. Consistent outreach activities are probably needed to improve the situation. Pharmacies are currently obliged to take back unused medicines, but that is additional work for them. It should be discussed how to make taking back medicines more attractive to pharmacies.

**Table 5.** Carbamazepine and diclofenac in environmental samples.

| Sample | Location | Time | carbamazepine<br>CAR [ng/l] | diclofenac<br>DIC [ng/l] |
| --- | --- | --- | --- | --- |
| 3 | Aquaculture 2, filter | 17.02.2020 | 0 | 61 |
| 7 | Aquaculture 4, filter | 17.02.2020 | 0 | 656 |
| 15 | Field 1, ditch | 20.01.2020 | 47 | 25 |
| 16 | Field 1, ditch | 20.01.2020 | 43 | 36 |
| 17 | Field 1, ditch | 20.01.2020 | 50 | 27 |
| 19 | Field 3, ditch | 28.04.2020 | 0 | 133 |
| 21 | Field 1, ditch | 28.04.2020 | 50 | 30 |
| 25 | Field 3, ditch | 28.08.2020 | 0 | 204 |
| 30 | Field 1, ditch | 01.03.2021 | 62 | 0 |
| 34 | Field 1, ditch | 19.04.2021 | 34 | 0 |

In summary, we conclude that drug residues reach the environment both through waste water treatment process (treated waste water, sewage sludge compost) and liquid manure. Finding out more precise parameters of the processes need further studies. In the case of sewage sludge, the efficiency of different composting technologies must be considered. In the case of liquid manure, the degradation of medicines during storage and the effect of adding composting steps should be studied. More information is needed about the transfer of drug residues from the soil to the environment. Here, it is also important to consider soil properties and rainfall. Special attention needs to be given to karst areas as there drug residues and microorganisms can more easily spread into groundwater.

The current study was based on observing functioning production and processing systems. It turned out that sometimes it is not possible to obtain accurate information from the farms. For example, information about liquid manure storage and spreading times is often not recorded in detailed manner. A more detailed study of the key issues influencing the release of medicines into the environment would be necessary by using an experimental approach - the use of experimental reactors and experimental fields.

## 6. Antibiotic resistance of microbes

### 6.1. Phenotypic resistance of microbial strains isolated from human clinical material

According to the Infectious Diseases Information System (NAKIS), 1,443 notifications were recorded in Estonia in 2020 on drug-resistant infectious agents. The most frequent findings were *E. coli* ESBL+ (932 notification), *K. pneumoniae* ESBL+ (286 notifications) and *S. aureus* MRSA (174 notifications). The same microbes were the most frequent findings in 2018 and 2019 as well (Health Board 2021). More detailed analysis of the infections was performed in current study.

#### Collection of microbial strains

The study was carried out in eight health care institutions participating in the European Antimicrobial Resistance Surveillance Network (EARSNet) (Tartu University Clinic, Northern Estonian Regional Hospital, Western Tallinn Central Hospital, Pärnu Hospital, East-Tallinn Central Hospital, East-Viru Central Hospital, Narva Hospital, SYNLAB Estonia).

Hospitalized patients and outpatients with ESBL-positive isolates were included in the study of *E. coli*, *K. pneumoniae* complex (*K. pneumoniae, K. variicola, K. quasipneumoniae subsp. quasipneumoniae, K. quasipneumoniae subsp. similipneumoniae*), methicillin-resistant *S. aureus* (MRSA) and vancomycin-resistant *E. faecium* (VRE). For each patient with a resistant strain, a patient with a sensitive microbial strain from the same medical institution or in ambulatory setting a consecutive patient with a sensitive microbial strain (ESBL-negative *E. coli, K. pneumoniae*, methicillin-susceptible *S. aureus* or vancomycin-susceptible *E. faecium*) was incuded. *Enterobacteriaceae* from hospitalized patients were isolated from blood, and strains from ambulatory patients were isolated from urine. *S. aureus* and *E. faecium* strains were isolated from various clinical materials (e.g. blood, urine, wound). During the study period (2019-2022) 202 pairs of resistant and sensitive strains, i.e. 404 strains, were collected (Table 6).

**Table 6.** Number of microbial strains collected.

| Bacterium | Clinical strains (n=234) | Ambulatory strains (n=170) |
| --- | --- | --- |
| *Staphylococcus aureus* | 22 | 8 |
| *Enterococcus faecium* | 28 | 2 |
| *Escherichia coli* | 120 | 102 |
| *Klebsiella pneumoniae* | 64 | 58 |

#### Phenotypic resistance of microbial strains

ESBL-positive *E. coli* and *K. pneumoniae* strains isolated from ambulatory and hospital materials were more resistant to fluoroquinolones, aminoglycosides, trimethoprim and trimethoprim/sulfamethoxazole (Tables 7 and 8). According to the literature, from the ESBL positive enterobacteria up to 90% are resistant to fluoroquinolones, 70% to aminoglycosides and 77-88% to trimethoprim/sulfamethoxazole (Azargun et al. 2018, Wiener et al. 2016, Khan et al. 2020, Xercavins et al. 2020, Teklu et al. 2019). Resistance of ESBL-positive strains to other groups of antibiotics is associated with specific ESBL genes (CTX-M, SHV, TEM) (Morosini et al. 2006, Canton et al. 2012). CTX-M genes often cluster in the same genetic element (plasmid, transposon, etc.) as the genes that give resistance to fluoroquinolones, aminoglycosides, trimethoprim/sulfamethoxazole and fosfomycin (Canton et al. 2012).

**Table 7.** Resistance of *E. coli* strains isolated from urine cultures of outpatients and blood cultures of hospital patients. Resistance (%) of ESBL-positive and ESBL-negative strains accordin to the criteria of the European Committee for Antibiotic Testing.

| Group of antibacterial agents | Antibacterial compound | Resistant strains (%) |  |  |  |
| --- | --- | --- | --- | --- | --- |
|  |  | Ambulatory *E. coli* strains (n=102) |  | Hospital *E. coli* strains (n=120) |  |
|  |  | ESBL neg | ESBL pos | ESBL neg | ESBL pos |
| Penicillins | Ampicillin | 11.8* | 100* | 63.8 | 100 |
|  | Mecillinam | 3.9 | 9.8 | 7.3 | 7.1 |
|  | Amoxicillin/clavulanic acid | 17.6* | 78.4* | 55.2* | 77.6* |
|  | Piperacillin | 11.8* | 96.1* | 60.3* | 79.3* |
|  | Piperacillin/tazobactam | 2* | 23.5* | 13.3 | 29.3 |
| Cephalosporins | Cefalexin | 2* | 100* | 31* | 100* |
|  | Cefuroxime | 45.1* | 100* | 35.8* | 99.2* |
|  | Ceftazidime | 0* | 98* | 10* | 84.17* |
|  | Ceftriaxone | 0* | 98* | 11.7* | 81.7* |
|  | Cefixime | 0* | 100* | 12.5* | 84.2* |
|  | Cefepime | 0* | 96.1 | 10* | 75* |
| Carbapenems | Ertapenem | 0 | 2 | 0 | 1.7 |
|  | Imipenem | 0 | 0 | 0 | 0 |
|  | Meropenem | 0 | 0 | 0 | 0 |
| Monobactams | Aztreonam | 0* | 43* | 10* | 24* |
| Fluoroquinolones | Norfloxacin | 0* | 78.4* | 32.8* | 58.6* |
|  | Levofloxacin | 0* | 52.4* | 29.8* | 50* |
|  | Ciprofloxacin | 0* | 66.7* | 29.8* | 52.7* |
| Aminoglycosides | Gentamicin | 15.7* | 49* | 46.6 | 60.3 |
|  | Tobramycin | 9.8* | 49* | 43.1* | 65.5* |
|  | Amikacin | 0* | 13.7* | 12.1 | 20.7 |
| Tetracyclines | Tigecycline | 7.8 | 5.9 | 11.1 | 7.9 |
| Other | Fosfomycin | 0 | 2 | 1.7 | 1.7 |
|  | Nitrofurantoin | 0 | 3.9 | 0 | 1.7 |
|  | Trimethoprim | 15.7* | 54* | 32.8 | 44.8 |
|  | TMP/SMX | 8.2* | 50* | 31 | 39.7 |

\* $p \leq 0.05$ (comparison of susceptible and resistant strains); TMP/SMX – trimethoprim/sulfamethoxazole

**Table 8.**
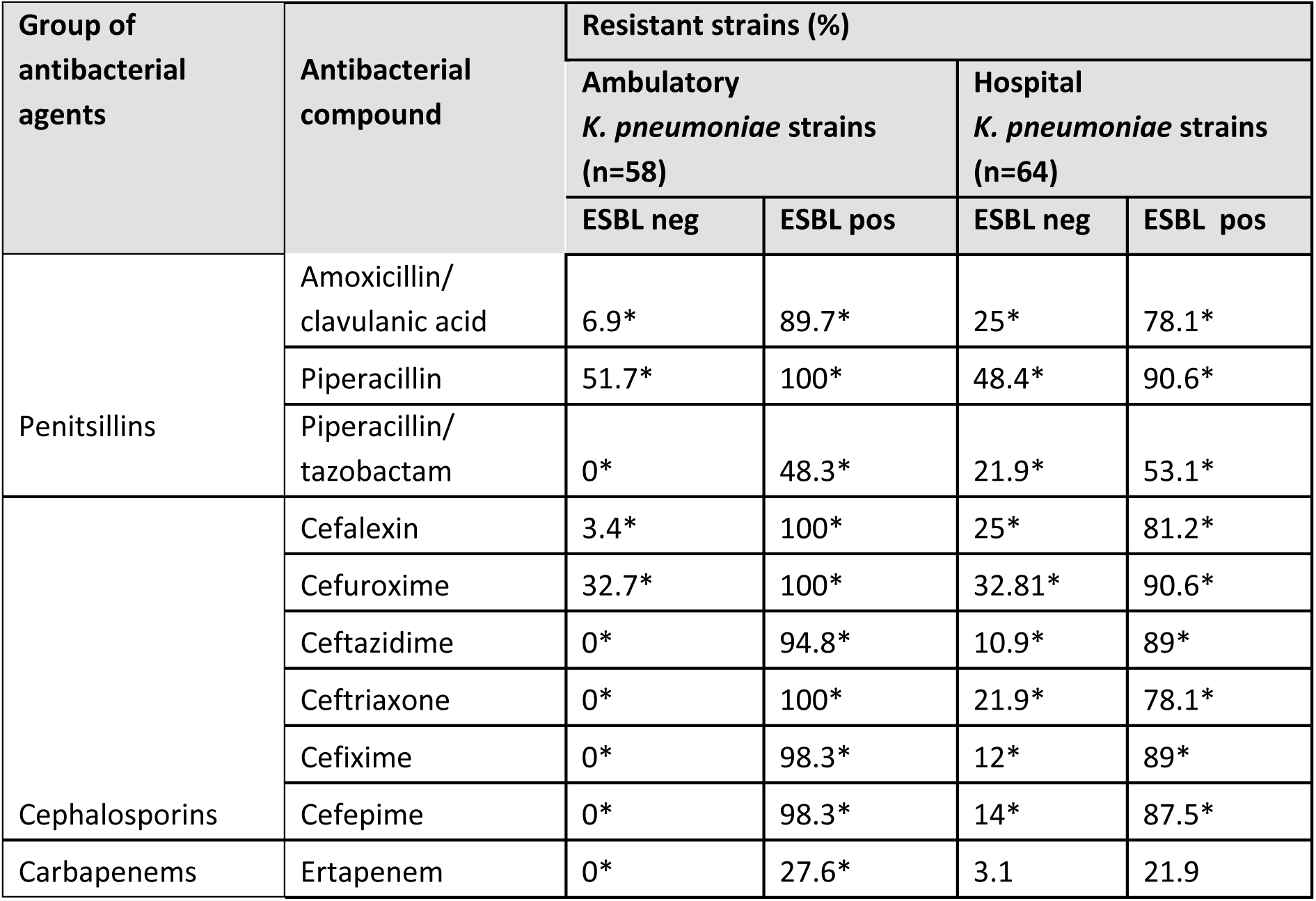
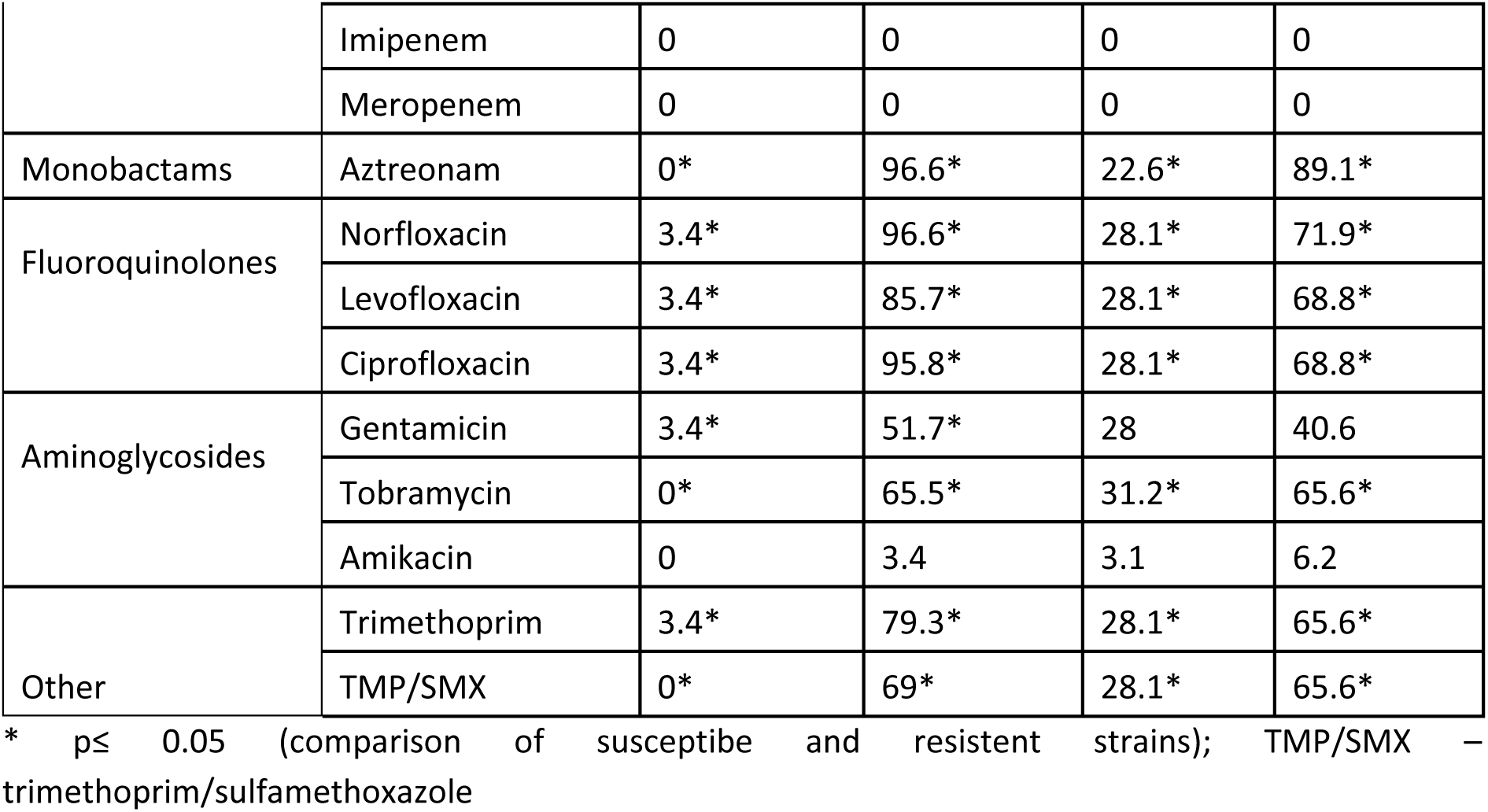
Resistance of *K. pneumoniae* strains isolated from urine cultures of outpatients and blood cultures of hospital patients. Resistance (%) of ESBL-positive and ESBL-negative strains accordin to the criteria of the European Committee for Antibiotic Testing.

| Group of antibacterial agents | Antibacterial compound | Resistant strains (%) |  |  |  |
| --- | --- | --- | --- | --- | --- |
|  |  | Ambulatory *K. pneumoniae* strains (n=58) |  | Hospital *K. pneumoniae* strains (n=64) |  |
|  |  | ESBL neg | ESBL pos | ESBL neg | ESBL pos |
| Penicillins | Amoxicillin/clavulanic acid | 6.9* | 89.7* | 25* | 78.1* |
|  | Piperacillin | 51.7* | 100* | 48.4* | 90.6* |
|  | Piperacillin/tazobactam | 0* | 48.3* | 21.9* | 53.1* |
| Cephalosporins | Cefalexin | 3.4* | 100* | 25* | 81.2* |
|  | Cefuroxime | 32.7* | 100* | 32.81* | 90.6* |
|  | Ceftazidime | 0* | 94.8* | 10.9* | 89* |
|  | Ceftriaxone | 0* | 100* | 21.9* | 78.1* |
|  | Cefixime | 0* | 98.3* | 12* | 89* |
|  | Cefepime | 0* | 98.3* | 14* | 87.5* |
| Carbapenems | Ertapenem | 0* | 27.6* | 3.1 | 21.9 |
|  | Imipenem | 0 | 0 | 0 | 0 |
|  | Meropenem | 0 | 0 | 0 | 0 |
| Monobactams | Aztreonam | 0* | 96.6* | 22.6* | 89.1* |
| Fluoroquinolones | Norfloxacin | 3.4* | 96.6* | 28.1* | 71.9* |
|  | Levofloxacin | 3.4* | 85.7* | 28.1* | 68.8* |
|  | Ciprofloxacin | 3.4* | 95.8* | 28.1* | 68.8* |
| Aminoglycosides | Gentamicin | 3.4* | 51.7* | 28 | 40.6 |
|  | Tobramycin | 0* | 65.5* | 31.2* | 65.6* |
|  | Amikacin | 0 | 3.4 | 3.1 | 6.2 |
| Other | Trimethoprim | 3.4* | 79.3* | 28.1* | 65.6* |
|  | TMP/SMX | 0* | 69* | 28.1* | 65.6* |

\* $p \leq 0.05$ (comparison of susceptible and resistant strains); TMP/SMX – trimethoprim/sulfamethoxazole

3/4 of ESBL-positive enterobacteria also proved to be resistant to amoxicillin/clavulanic acid and 1/3 of *E. coli* and nearly half of *K. pneumoniae* strains also to piperacillin azobactam. Therefore, it would be important to reduce the use of penicillin / beta-lactamase inhibitor combinations in human medicine.

Resistance of ESBL-negative enterobacteria isolated from ambulatory urine samples to methicillin, 3.-4. generation cephalosporins, fluoroquinolones, nitrofurantoin and to trimethoprim/sulfamethoxazole was low. Methicillinam and nitrofurantoin are suitable for the treatment of ambulatory urinary infections. An alternative would be trimethoprim or trimethoprim/sulfamethoxazole, as resistance of ESBL-negative enterobacteria was below 20% (Gupta et al. 2011). From hospital strains more than 20% of ESBL-negative strains were resistant to the fluoroquinolones, trimethoprim and trimethoprim/sulfamethoxazole. If the resistance to trimethoprim/sulfamethoxazole is above 20% and fluoroquinolone resistance above 10%, then the use of these antibiotics is not recommended in the empiric treatment of urinary tract infections (Gupta et al 2011).

Compared to methicillin-susceptible *S. aureus* strains, MRSA strains had higher MICs for beta-lactams, fluoroquinolones and aminoglycosides (Table 9). Table 8 uses MIC values because for most prescribed antibiotics Gram-positive cocci lack EUCAST criteria to differentiate sensitive and resistant strains.

**Table 9.** Comparison of minimal inhibitory concentrations (MICs) of Gram-positive microorganisms (methicillin-sensitive *S. aureus* vs. methicillin-resistant *S. aureus* and vancomycin-susceptible *E. faecium* vs. vancomycin-resistant *E. faecium*).

| Group of antibacterial agents | Antibacterial compound | MIK<sub>50</sub> |  | MIK<sub>50</sub> |  |
| --- | --- | --- | --- | --- | --- |
|  |  | Methicillin-susceptible *S. aureus* | Methicillin-resistant *S. aureus* | Vancomycin susceptible *E. faecium* | Vancomycin resistant *E. faecium* |
| Betalactams | Penicillin | 0.250" | 0.250" | 0.250 | 0.25 |
|  | Ampicillin | 1 | 1 | 16 | 16 |
|  | Oxacillin | 0.250" | 2" | 2 | 2 |
|  | Moxalactam | 16" | 16" | 16 | 16 |
|  | Cefoxitin | 2" | 8" | 8 | 8 |
|  | Ceftriaxone | 0.250" | 0.5" | 1 | 1 |
|  | Imipenem | 1" | 1" | 8 | 8 |
| Amino-glycosides | Kanamycin |  |  | 32 | 32 |
|  | Gentamicin | 1" | 2" | 4 | 4 |
|  | Tobramycin | 1" | 2" | 4 | 4 |
| Fluroquinolones | Ciprofloxacin | 0.5" | 0.5" | 4 | 4 |
|  | Levofloxacin | 0.5" | 0.5" | 4 | 4 |
|  | Moxifloxacin | 0.250" | 0.250" | 2 | 2 |
| Glycopeptides and lipopeptides | Vancomycin | 1 | 1 | 0.5" | 8" |
|  | Teicoplanin | 0.5 | 0.5 | 0.5" | 0.5" |
| Macrolides, lincosamides, streptogramins | Erythromycin | 0.250" | 4" | 4 | 4 |
|  | Clindamycin | 0.250" | 0.250" |  |  |
|  | Pristinamycin |  |  | 0.5 | 0.5 |
|  | Quinupristin / Dalfopristin | 0.5 | 0.5 | 1" | 0.5" |
| Tetracyclines, oxazolidinones | Tetracycline | 0.5 | 0.5 | 2" | 0.5" |
|  | Tigecycline | 0.125 | 0.125 |  |  |
|  | Linezolid | 1 | 1 | 2 | 2 |
| Other | Daptomycin | 0.5" | 0.5" | 1 | 1 |
|  | Fosfomycin | 16 | 16 | 64 | 64 |
|  | Chloramphenicol | 4 | 4 | 6 | 4 |
|  | Fusidic acid | 0.5 | 0.5 | 4 | 4 |
|  | Mupirocin | 0.5 | 0.5 | 2 | 2 |
|  | Nitrofurantoin | 16 | 16 | 64 | 64 |
|  | Trimethoprim | 2" | 2" | 0.5 | 0.5 |
|  | TMP/SMX | 0.5 | 0.5 | 0.5 | 0.5 |

" $p \leq 0.05$ (MIC comparison); TMP/SMX – trimethoprim/sulfamethoxazole

ESBL-negative *E. coli* and *K. pneumoniae* strains isolated from the hospital had higher MICs for penicillin / beta-lactamase inhibitor combinations, 3.-4. generation cephalosporins, aztreonam, fluoroquinolones, aminoglycosides, trimethoprim and trimethoprim/sulfamethoxazole when compared with ESBL-negative strains isolated from outpatient material. Our previous study also showed that enterobacteria isolated from the blood of hospital patients are more resistant than those from urine samples of outpatients (Sepp et al. 2020). There is higher use of antibiotics in the hospital, which causes selection of more resistant strains and colonization of patients with antibiotic resistant strains. When antibacterial treatment for outpatients is prescribed, it is important to consider the previous stay in the hospital, because patients may be colonized with resistant strains.

### 6.2. Phenotypic resistance of microbial strains isolated from animals and food Collection of samples

The collection of samples in the veterinary sector lasted two years (01.09.2019-30.09.2021). Together 1625 samples from different animal species and food were analyzed (Figure 18).

**Figure 18.**
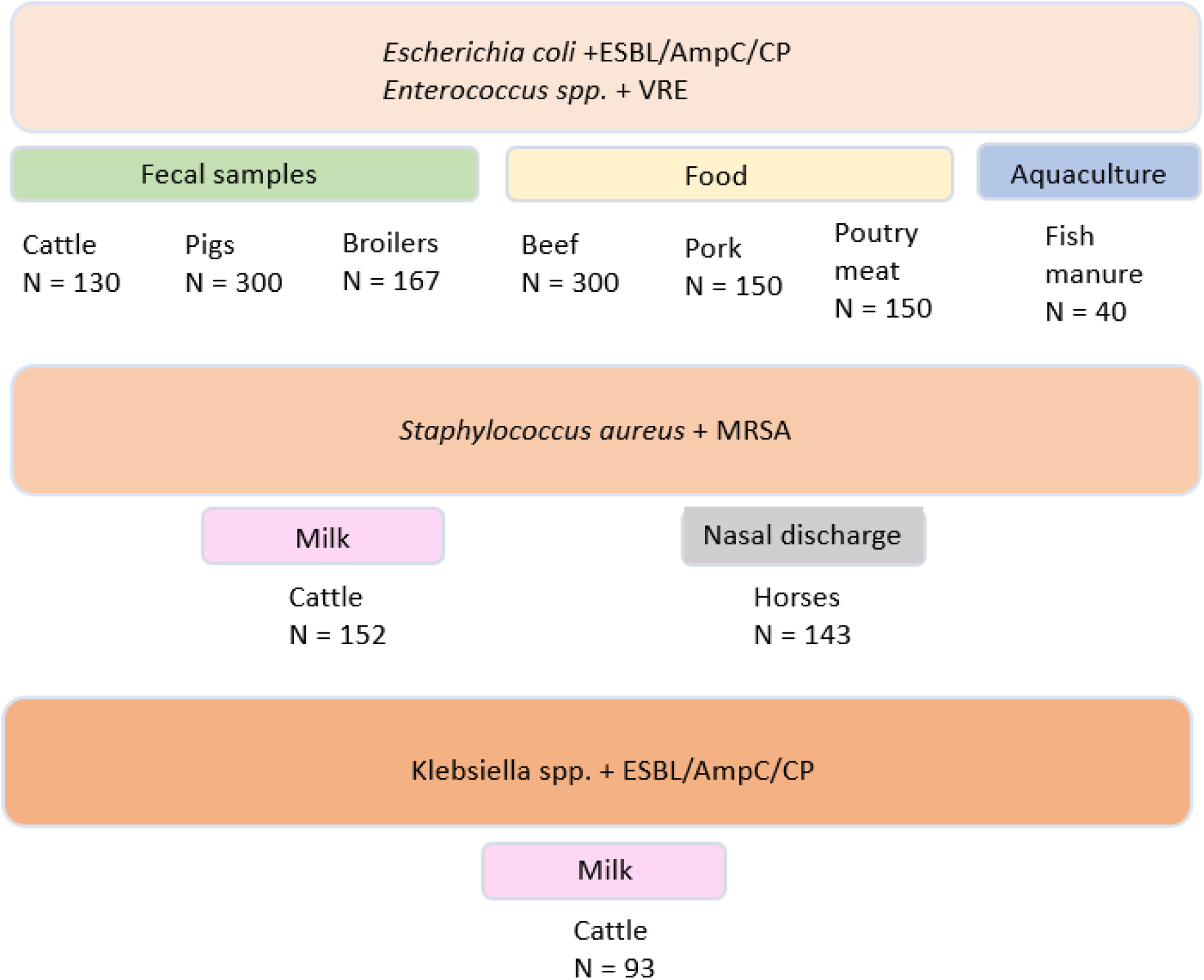
Number, origin and investigated microbial species of samples from the veterinary field.

#### 6.2.1. Antibiotic resistance of *E. coli* in Estonia in 2017-2021 Phenotypic antibiotic resistance of isolates from pigs, broilers and cattle

Antibiotic resistance of ESBL/AmpC-negative *E. coli* isolates from clinically healthy pigs (n=281) has not decreased in three-year comparison (2017 vs 2019 vs 2021). In 2017 11.9% of ESBL- and/or AmpC-negative *E. coli* isolates were multidrug-resistant (simultaneous resistance to three or more active substances); in 2019 the corresponding number was 21.2%, and in 2021 18.4%. Simultaneous resistance to ampicillin, tetracycline and trimethoprim was most common. From all isolates collected during the three study years (n=394) ESBL and/or AmpC producing *E. coli* phenotype was detected in 24 (26.3%) isolates in 2017, 37 (34.2%) isolates in 2019 and 52 (32.9%) isolates in 2021. Thus, the proportion of strains producing ESBL and AmpC beta-lactamases did not change during the period of the study. No carbapenem-resistant strains have been found in pigs.

In the comparison of 2018 and 2020, in *E. coli* isolated from clinically healthy broilers, antibiotic resistance decreased among ESBL- and/or AmpC-negative strains (n=167). In 2018, ESBL- and/or AmpC-negative *E. coli* isolates included 61.2% multidrug-resistant isolates; in 2020 the corresponding number was 19%. From all isolates collected from broilers during the two years (n=278) 111 isolates were found to produce ESBL/AmpC type beta-lactamases. The proportion of ESBL/AmpC producing isolates has decreased, being 44.8% in 2018 and 34% in 2020.

#### Antibiotic resistance of isolates from cattle and swine farm manure

In 2019, from the 115 studied dairy farms 50 *E. coli* strains producing ESBL and/or AmpC type beta-lactamases were found in 50 farms (43.4% of farms). For pigs, manure samples were collected from 15 farms. Strains producing ESBL/AmpC type *E. coli* were isolated in eight pig farms (53.3%).

#### Antibiotic resistance of isolates from pork, broiler and beef

From the pork (n=300) and beef (n=300) samples collected during the two study years, 39 *E. coli* strains producing ESBL and/or AmpC-type beta-lactamases were identified. Beef and pork samples in 2019 eight (2.7%) and in 2021 18 (6%) ESBL/AmpC-positive E. *coli* isolates were found. For pork and beef samples there was statistically significant relationship (p=0.005) between the country of sample origin (domestic vs. imported) and the occurrence of ESBL/AmpC-positive *E. coli*. There were more resistant strains in the imported meat.

130 different sequence types were found in ESBL-positive *E. coli* strains isolated from animals and food (n=310). The six most common sequence types were present in 91 isolates (Figure 19).

**Figure 19.**
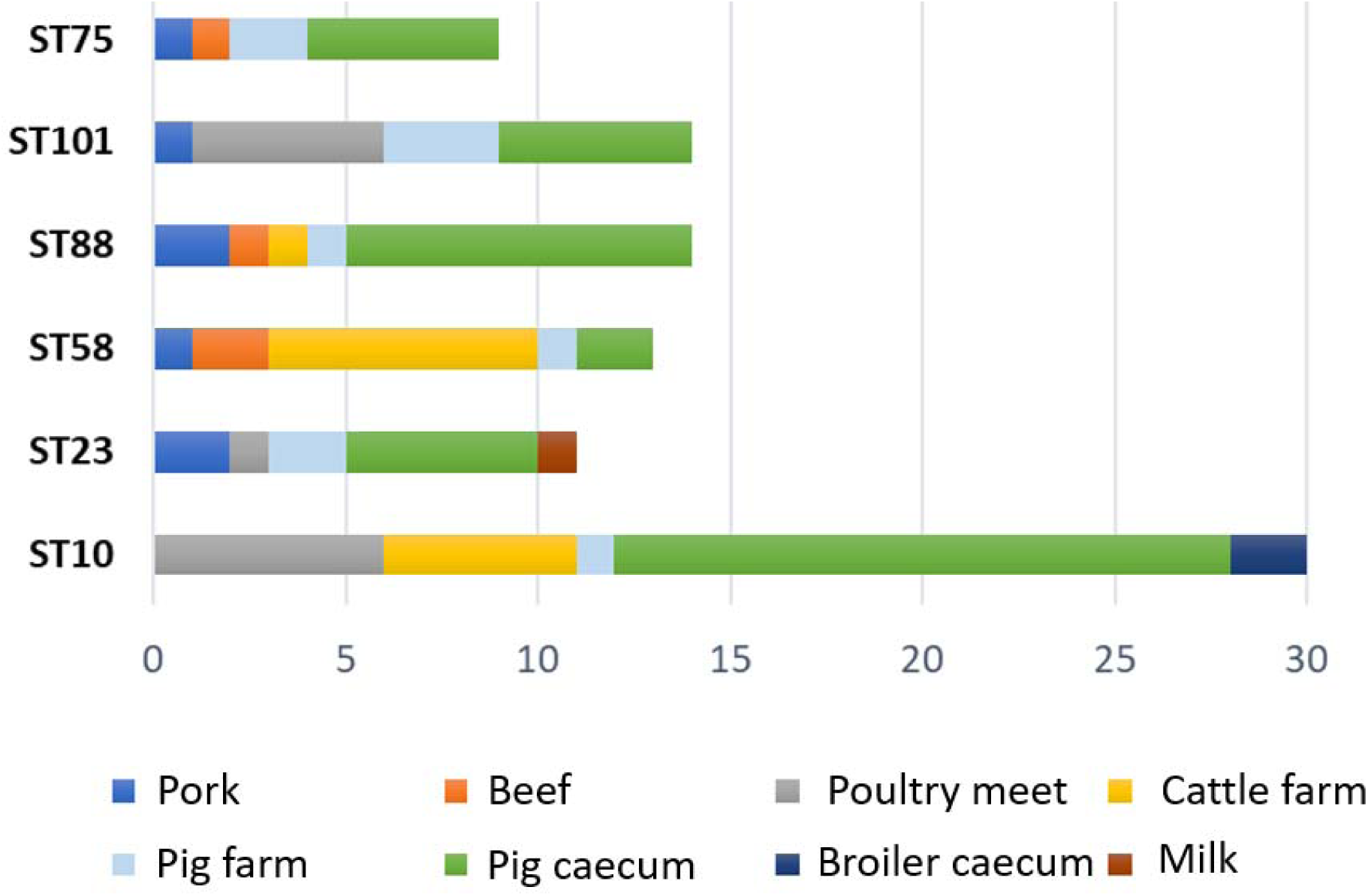
Sequence type distribution of ESBL-positive *E. coli* strains isolated from animals and food.

The distribution of beta-lactamases is characterized in Figure 20. A total of 22 different beta-lactamase types were found in meat samples and 19 in live animals. The most frequent were blaTEM-1B and blaTEM-52C. Among the CTX-M types was the most common blaCTX-M1 in meat and live animals. AmpC-type beta-lactamases were present in both groups of samples, the most frequent being blaCMY-2.

**Figure 20.**
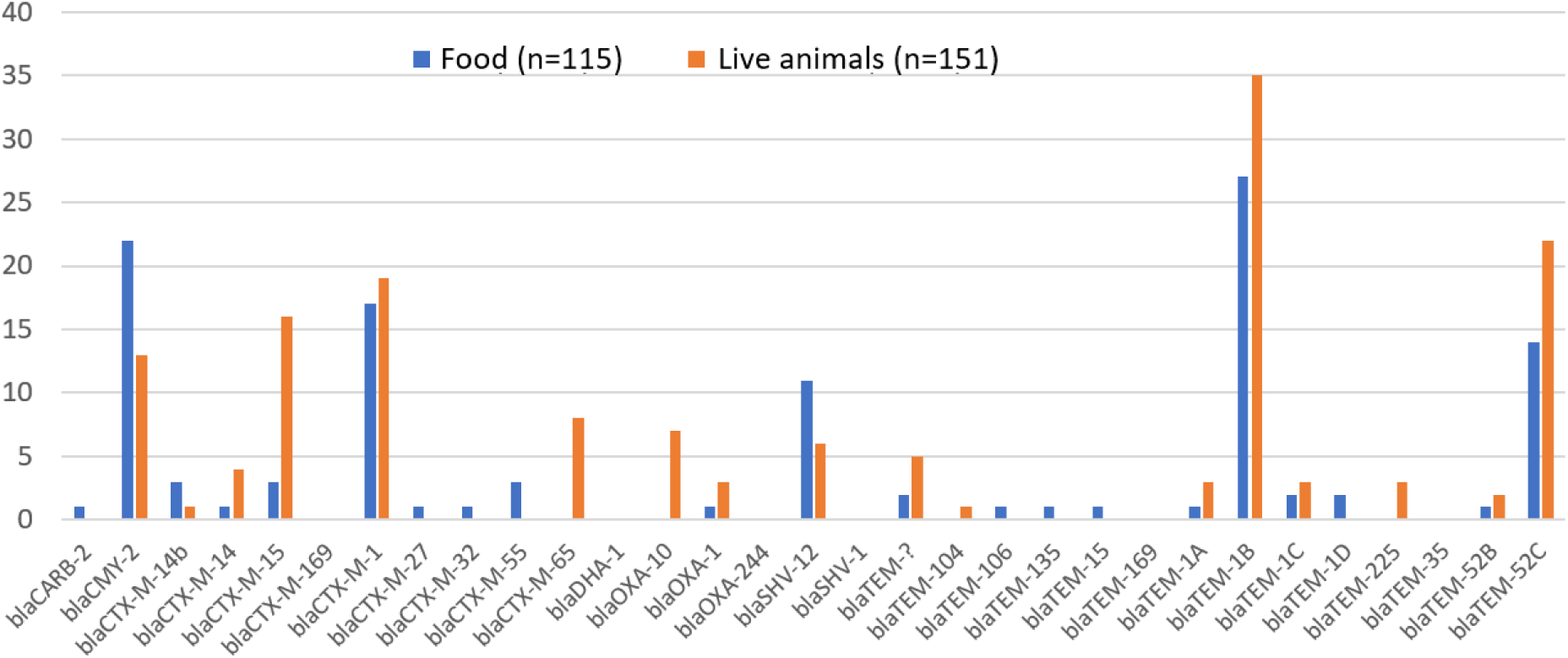
Distribution of different beta-lactamases among *E. coli* isolated from meat and live animals.

#### Prevalence of ESBL/AmpC-type beta-lactamase-producing *E. coli* in nearby countries

Comparative data of neighboring countries were collected on the basis of EU monitoring data (year 2019). In clinically healthy pigs the prevalence of ESBL/AmpC producing *E. coli* varies between countries. The nordic countries the precentage of positive strains was between 2.4% (Finland) and 26.7% (Denmark). In caecal samples of pigs from the Baltic countries and Poland, ESBL/AmpC-positive *E. coli* was present in 23.2 - 54.6% of isolates (Figure 21).

**Figure 21.**
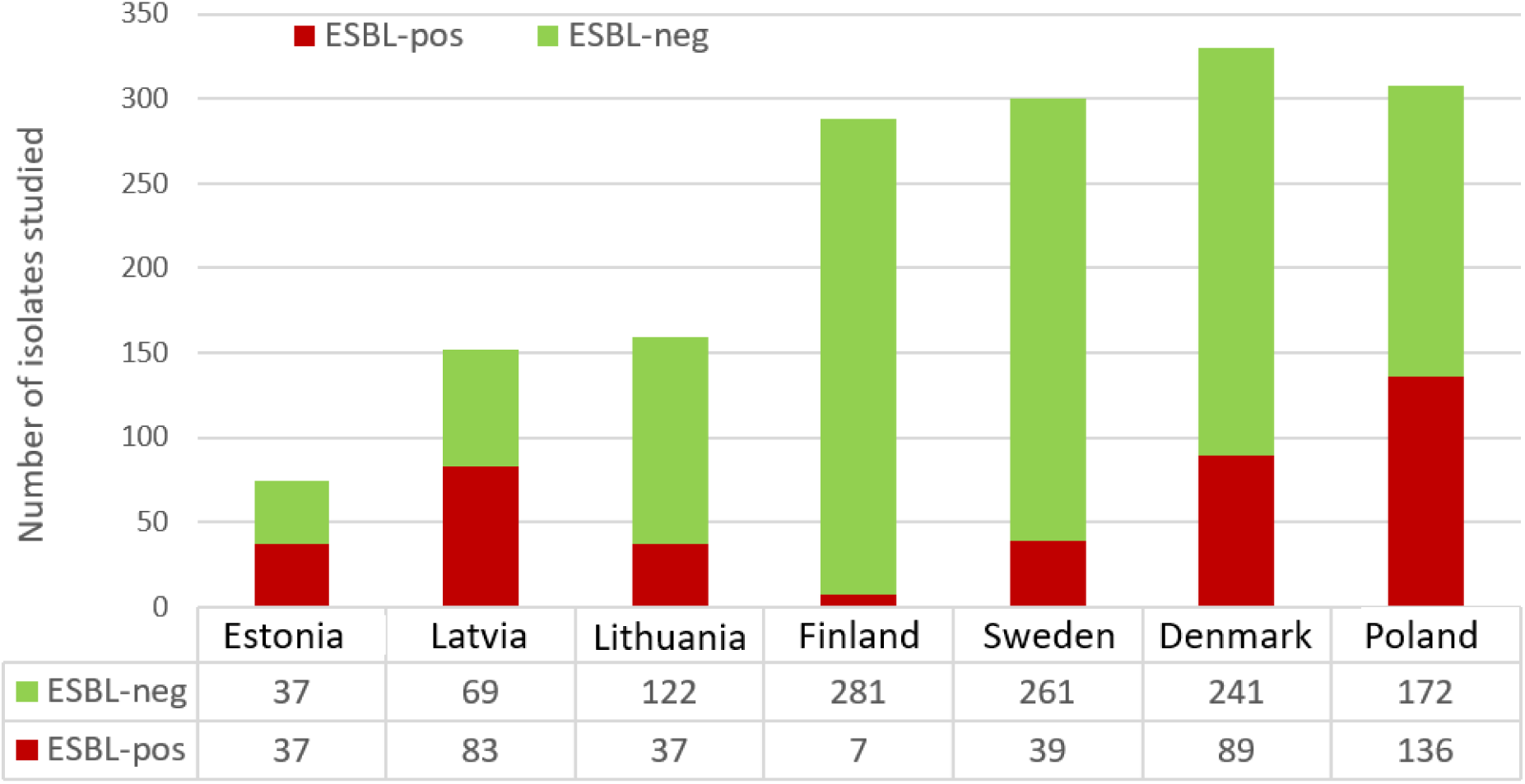
ESBL/AmpC isolated from cecums of clinically healthy pigs from Nordic and Baltic countries. Number of positive *E. coli* isolates in 2019.

Also, the prevalence of ESBL/AmpC positive *E. coli* isolated from ceacal samples of clinically healthy broilers was different between countries, the occurrence in the Nordic countries was significantly lower than in the Baltic countries (Figure 22).

**Figure 22.**
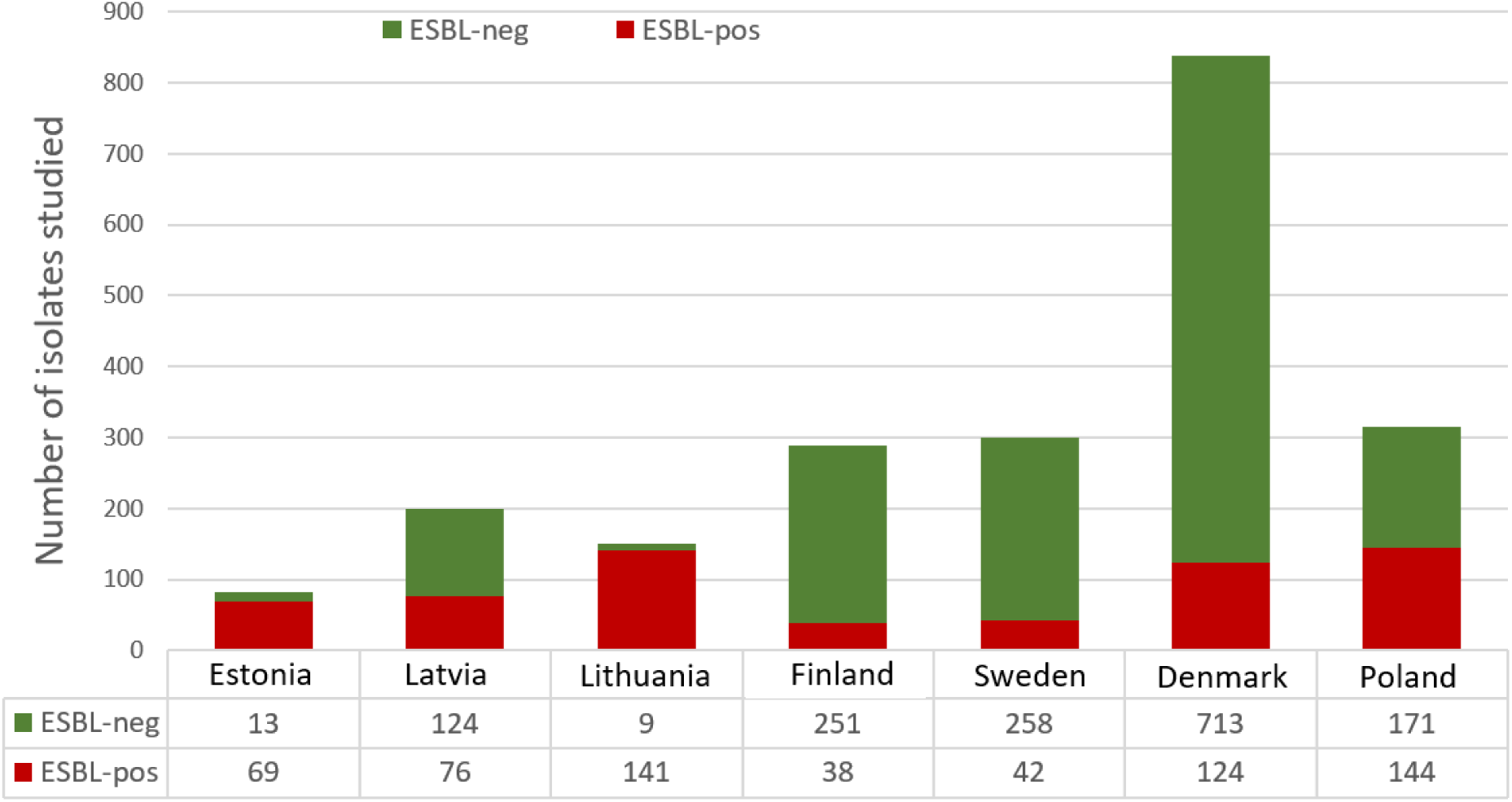
ESBL/AmpC isolated from cecum of clinically healthy broilers from Nordic and Baltic countries. Number of positive *E. coli* isolates in 2019.

#### 6.2.2. Antibiotic resistance of enterococci in Estonia in 2017-2021

##### Phenotypic antibiotic resistance of *Enterococcus* spp from cecum of clinically healthy pigs in 2017 and 2019

In 2017 and 2019, a total of 81 *E. faecalis* and 177 *E. faecium* strains were isolated from the caecum of pigs. Vancomycin-resistant enterococci from cecums of clinically healthy pigs were not isolated during the study period. There has been a decrease in the proportion of ciprofloxacin-resistant *E. faecium* strains in comparison of the two sample collection periods (27.6% vs. 1.8%). Ampicillin, tetracycline and erythromycin resistances have remained relatively stable.

##### Phenotypic antibiotic resistance of *Enterococcus* spp from cecums of clinically healthy broilers in 2018 and 2020

During the two years of the study, a total of 117 *E. faecalis* and 125 *E. faecium* strains were isolated from the caecum of broilers. Vancomycin-resistant enterococci were not detected in the caeca of clinically healthy broilers. The proportion of ciprofloxacin resistant *E. faecium* strains decreased between the two sample collection periods (31.3% vs. 15.6%). For ampicillin, tetracycline and erythromycin resistances the levels remained relatively stable.

#### 6.2.3. Antibiotic resistance in *Staphylococcus* spp. in 2019-2021

##### Occurrence of methicillin-resistant staphylococci in the nasal passages of horses, milk of cattle and organ samples from pigs

For detection of methicillin-resistant *S. aureus* (MRSA), a total of 141 nasal swab samples were collected from horses living in Estonia. For the isolated staphylococci (n=20), the antibiotic susceptibility was determined. Coagulase-negative staphylococci were found the most cases, the dominant species was *S. delphini* (9/16, 45%). This distribution of species is expected in light of our previous data. In total four S. *aureus* isolates were found, two of which proved to be methicillin resistant. The prevalence of MRSA in the horse population studied was 1.4% (CI 95% 0.17-5.03). Both MRSA isolates were from genotypic group ST398. The MRSA-negative staphylococci isolated were often resistant to fusidic acid (50%) and trimethoprim/sulfamethoxazole (18%).

In 2020, 152 refrigerated milk samples were collected from cattle farms with more than 100 cattle. In Estonia there are approximately 200 dairy farms of this size, which means that 75% of the farms were included in the survey. Every farm was sampled with one sample. MRSA with genotype ST398 was detected in eight refrigerated milk samples. Prevalence of MRSA in refrigerated milk was 5.3% (CI 95% 2.7-10.1).

In 2020, 103 samples from clinical pigs were sent to the laboratory for MRSA testing. *S. aureus* was found in total of seven times and two of the cases were MRSA. Both of the samples were from the same farm and both isolates belonged to the ST398.

#### 6.2.4. Antibiotic resistance of isolates from aquaculture

40 fish manure and cleaning filter samples from 12 fish farms were analyzed for vancomycin-resistant enterococci and ESBL/AmpC *E. coli*. No positive cases were observed.

### 6.3. Characterization of bacterial strains based on genomic data

The aim of the study was to analyze the prevalence of *E. coli*, *K. pneumonia* and *S. aureus* strains based on genomic sequences.. Smaller number of strains was collected and analyzed for *E. faecium*, *E. fecalis* and *Salmonella enterica*. Recent transfers (those that could occur during the last 5-10 years) between human-animal-food-environment segments were not observed. The transmissions are therefore not very frequent, and we need a larger data collection to estimate the frequency of transfer more accurately.

#### 6.3.1. Escherichia coli

A total of 174 different *E. coli* sequence types were detected, of which the five most common were ST131 (n=62; 10.8%). ST10 (n=49; 8.6%), ST69 (n=25; 4.4%), ST95 (n=22; 3.8%) and ST73 (n=19; 3.3%). 14 new STs were also identified STs that were not in the PubMLST database. ST131 was the most frequent ST isolated from humans. Among the strains isolated from animals, food and the environment, ST10 was the most common. ST10 was also the most wide spread ST, as it was found in all segments studied (Table 10).

**Table 10.** Common *E. coli* sequence types in different segments.

| Human |  | Animals |  | Environment |  | Food |  |
| --- | --- | --- | --- | --- | --- | --- | --- |
| ST | Number | ST | Number | ST | Number | ST | Number |
| 131 | 61 | 10 | 31 | 10 | 8 | 10 | 6 |
| 95 | 22 | 88 | 14 | 1079 | 3 | 101 | 5 |
| 69 | 21 | 1079 | 13 | 101 | 1 | 752 | 5 |
| 73 | 20 | 58 | 10 | 3232 | 1 | 93 | 4 |
| 404 | 7 | 101 | 9 | 38 | 1 | 115 | 3 |
| 1193 | 6 | 23 | 9 | 877 | 1 | 117 | 3 |
| 12 | 6 | 75 | 7 |  |  | 23 | 3 |
| 10 | 4 | 1611 | 6 |  |  | 57 | 3 |
| 127 | 4 | 164 | 5 |  |  | 58 | 3 |
| 141 | 4 | 542 | 5 |  |  | 746 | 3 |

However, strains with the same ST that occurred in different segments are from the core genome sequences very different, indicating that no recent transfers between different segments have occurred. The most closely related strains isolated from different segments differ on the basis of the core genome by a minimum of about 30 nucleotides. Considering the frequency of evolutionary fixed mutations to be about 4 - 10 nucleotides per year (Duchêne et al., 2016; Lagos et al., 2022), it places the possible transfer to a period 3-5 years ago.

The analysis showed that the ESBL phenotype is relatively evenly distributed on the phylogenetic tree. However, ST131, the most prevalent ST in humans, is almost entirely ESBL-positive. This is a problem because these are mainly invasive strains.

When compared to the period 2010-2014 we see that ST131 continues to dominate in clinical strains (Fig 23). However, the proportion of ST10 has increased significantly in strains from the environment and animals (Figures 24 and 25).

**Figure 23.**
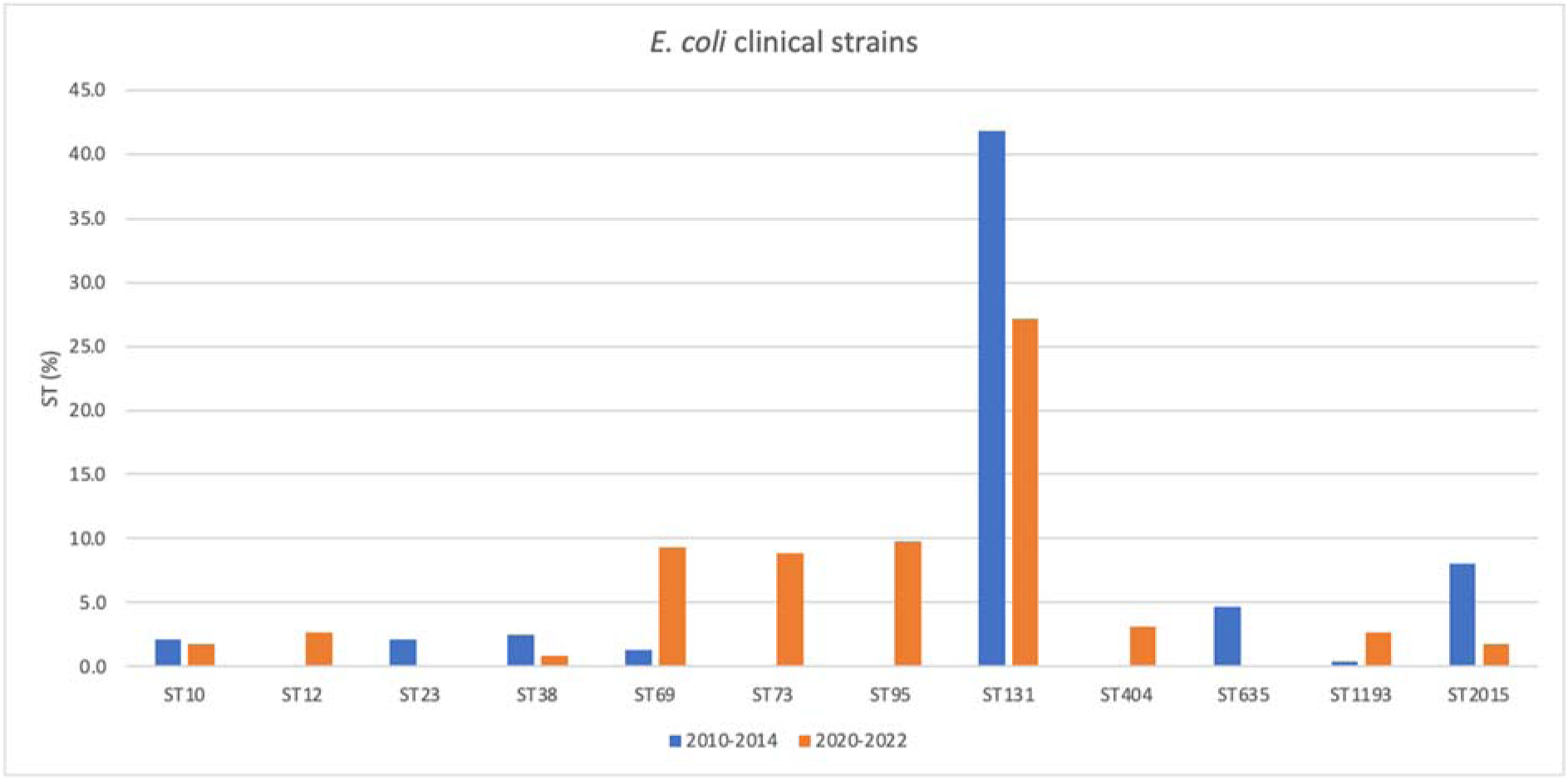
Comparison of ST distribution of *E. coli* clinical strains collected at different time periods.

**Figure 24.**
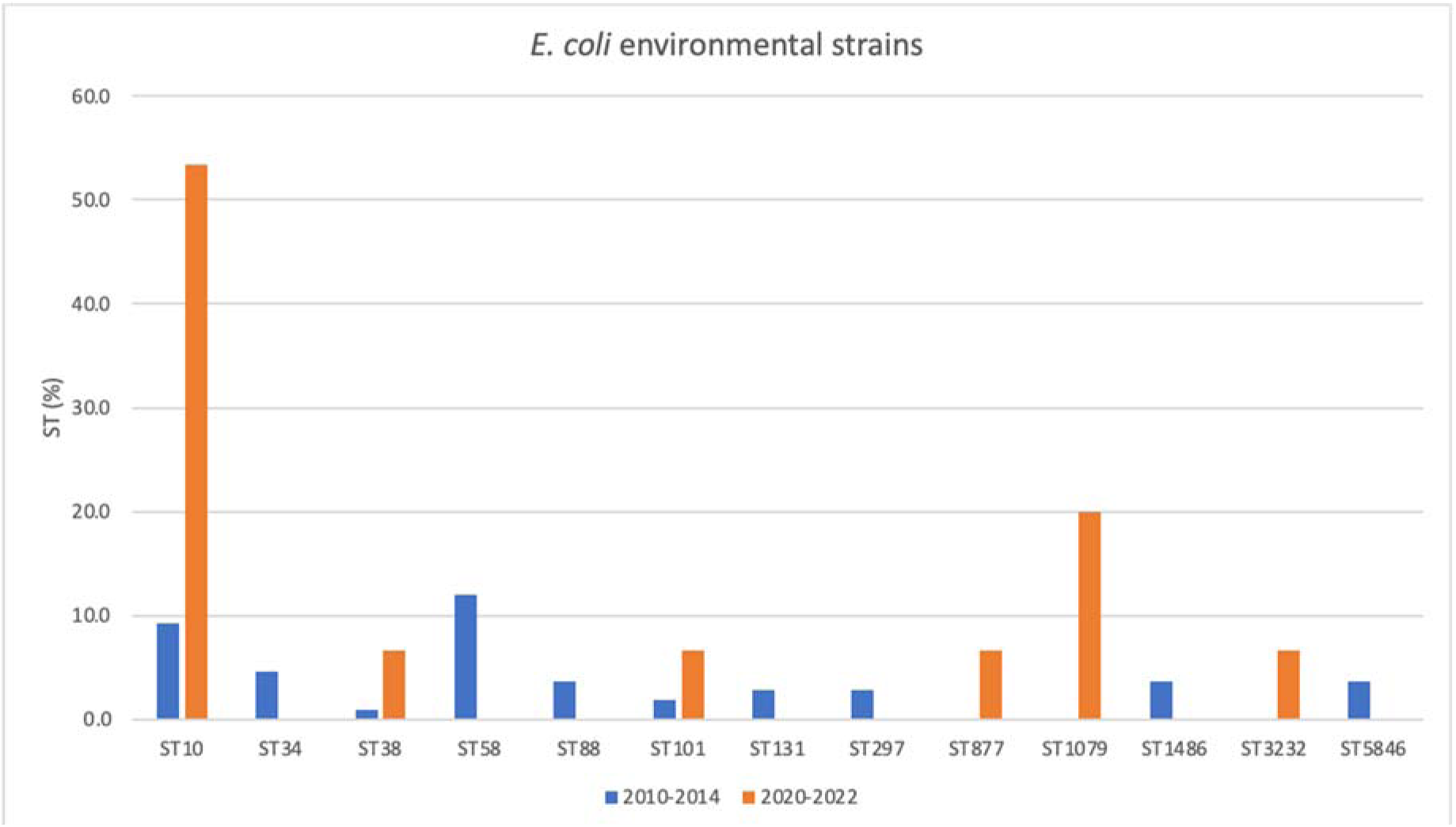
Comparison of *E. coli* environmental strains collected at different time periods.

**Figure 25.**
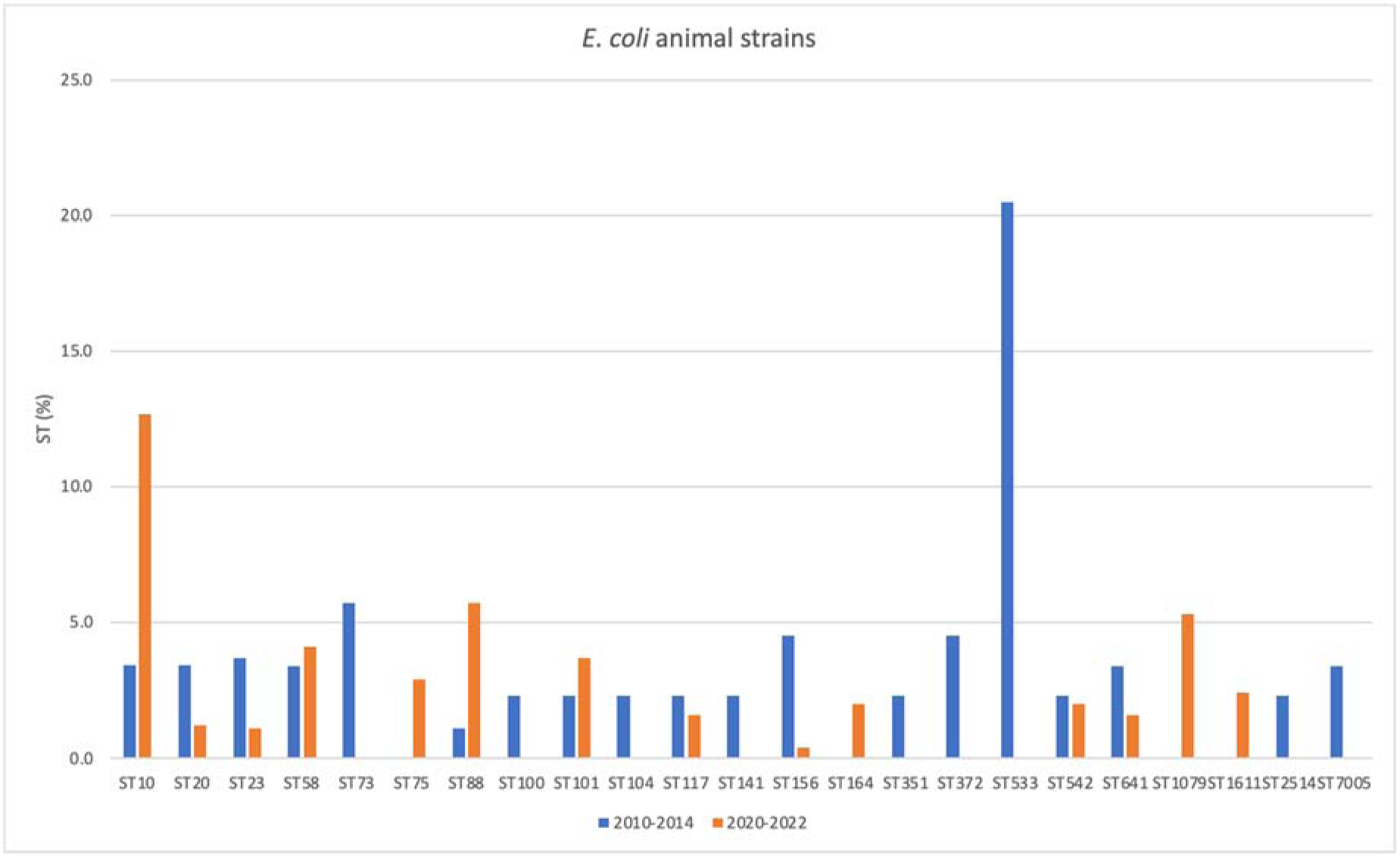
Comparison of animal-derived *E. coli* strains collected at different time periods.

Plasmid-borne *mcr* genes that confer resistance to colistin are seen as a significant threat (Xu et al. 2022). No such genes were identified in the present study, although there has been a colistin-resistant strain of *E. coli* previously isolated in Estonia (Brauer et al. 2016).

#### 6.3.2. Klebsiella pneumoniae

A total of 69 different STs were detected, of which the five most common were ST307 (n=20; 14.7%), ST15 (n=12; 8.8%), ST395 (n=6; 4.4%), ST86 (n=6; 4.4%) and ST219 (n=5; 3.7%).

All these STs were present among human strains. Five new STs not in the PubMLST database were identified.

ESBL-positive ST307 is common in North America (Long et al. 2017), South America (Andrade et al. 2018) and in Europe (David et al. 2019). ST307 was predominantly ESBL-positive in our study. Previously, in the period 2010-2014 ST307 was not found in Estonia. It seems to be a worldwide widespread cluster that has now also reached Estonia (Figure 26).

**Figure 26.**
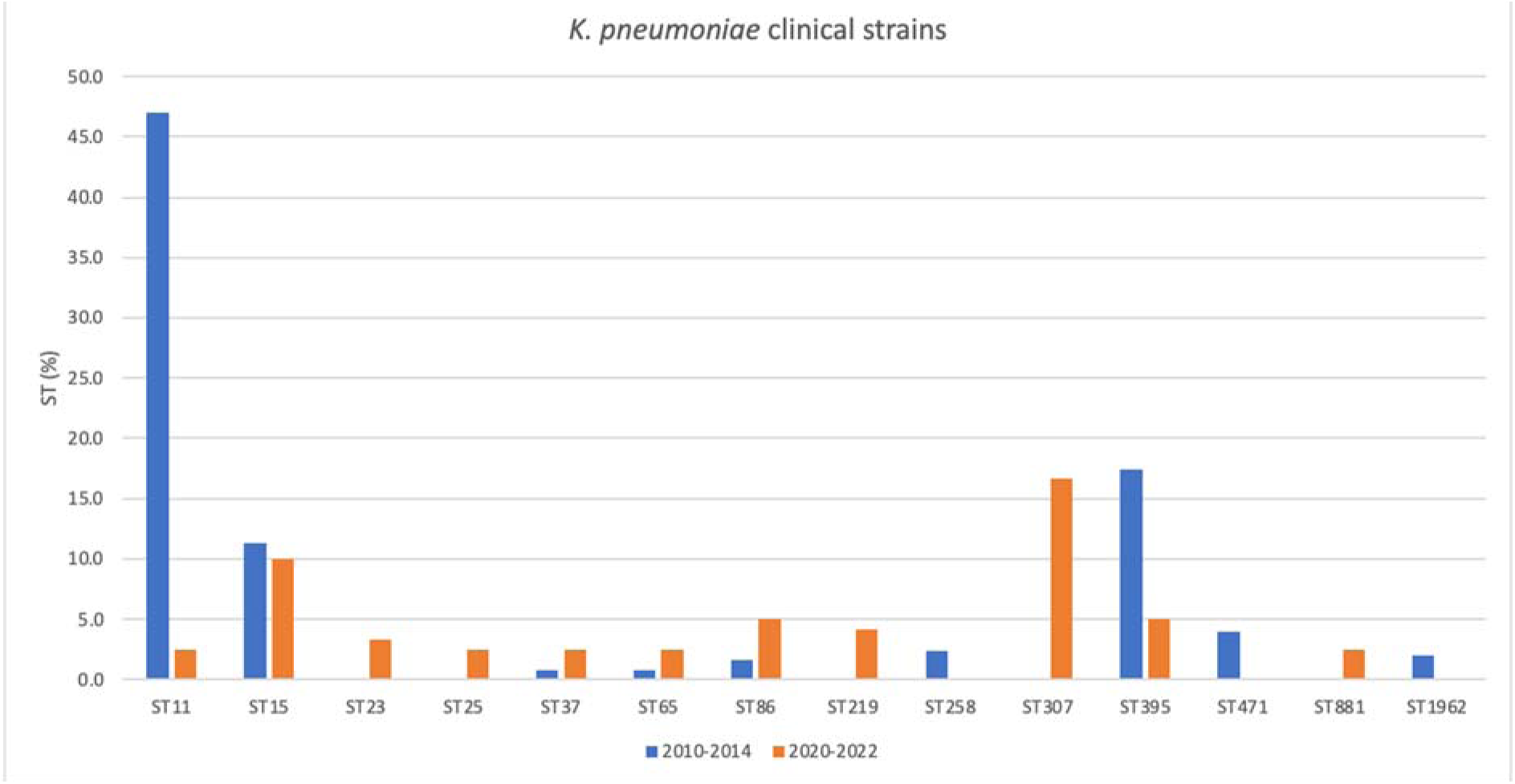
Comparison of *K. pneumoniae* clinical strains collected at different time periods.

*K. pneumoniae* is characterized by the presence of specific sequence types in humans and animals. ST111 (n=3; 2.2%) and ST187 (n=2; 1.5%) were the only ones found in both segments but the large differences in the core genome exclude a recent transfer. According to the literature, *K. pneumonia* has potential for zoonotic transmission, but the strains are still poorly characterized and we do not know the extent of transmission (Hu et al 2021). Based on the data of this work, we did not detect very frequent transmission. The *mcr* genes that spread on plasmids were bnot found.

#### 6.3.3. Staphylococcus aureus

A total of 16 different *S. aureus* STs were identified, all of which were found to be specific either for animals or humans. The three most common sequence types were ST88 (n=28; 43.1%), ST398 (n=12; 18.5%) and ST45 (n=5; 7.7%). The ST88 strains were found only in humans; all ST398 strains were isolated from animals.

ST88 is a MRSA cluster originally spread in Africa (Kpeli et al. 2017). In the current study it was also observed that it is predominantly methicillin-resistant cluster. All ST88 strains were from one outbreak in Tallinn hospitals. ST398 is a common ST found in livestock. The ST can sometimes be transmitted to humans, but usually does not cause disease in humans (Lienen et al. 2021).

#### 6.3.4. Vancomycin-resistant enterococci (VRE)

Based on the number of previous VRE findings in clinical samples, it was expected in the start of the project that the prevalence of VREs is increasing rapidly. Fortunately, this has not happened and we only isolated a small number of strains. Data from earlier period have been published in the article Aun et al. 2021.

Strains with the same ST that would have spread in different segments and would indicate transmission were not found. For *E. faecium* 14 STs were found and 4 for *E. facealis*. Three of them are new, which are not currently in the PubMLST database. Among VRE+ *E. faecium* strains, the most common was vancomycin-resistant ST117 (n=12; 33.3%).

#### 6.3.5. Salmonella enterica

All collected *Salmonella enterica* subsp. enterica serovar Mbandaka strains belonged to ST314 and were isolated from animals. *Salmonella enterica* subsp. *enterica* serovar Typhimurium isolated from the environment was from ST34. ST314 has been associated with very high levels of antibiotic resistance (Wang et al. 2021), but no resistance was detected in strains isolated in Estonia.

### 6.4. AMR trends of ambulatory important microbes in 2008-2018 and comparison with neighboring countries

Data from Latvia, Finland, Sweden and Denmark, which were collected in EARS-Net, were used for the analysis of resistance trends.

Resistance of invasive *E. coli* strains to aminopenicillins (ampicillin) is stably high in Estonia, but slightly decreasing:if in 2012 48% of strains were resistant, then in 2018 43% of strains were resistant (Figure 26). Resistance to 3rd generation cephalosporins (ceftazidime, cefotaxime) ranged from 7.4 to12.2% and has remained stable (Figure 27).

**Figure 27.**
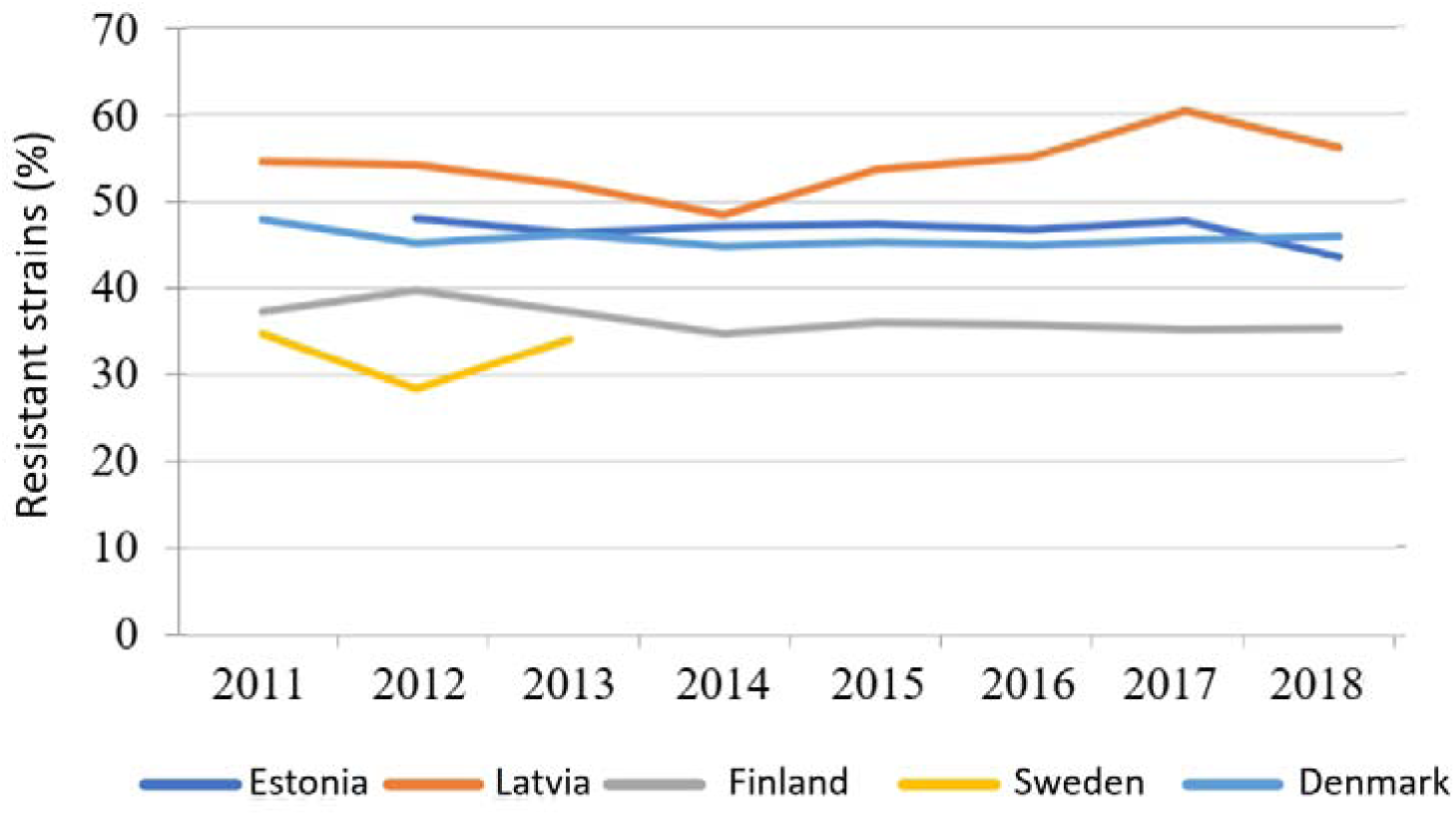
Resistance of *E. coli* to aminopenicillins.

Resistance to fluoroquinolones (ciprofloxacin) is relatively stable (12-17%)(Figure 28). In 2018, 6.2% of *E. coli* strains were resistant to aminoglycosides. In 2018, no carbapenemase-producing E. *coli* strains was reported in the countries under study. In comparison to neighboring countries, in 2018, the highest number of resistant *E. coli* strains was reported in Latvia and the lowest number in Finland (Figures 27-29). Of the observed countries, only Latvia had resistance of invasive strains to 3rd generation cephalosporins higher than the European Union average (the EU average was in 2018 was15.1%). In Denmark, the resistance of *E. coli* strains is low, but the rise of resistance to fluoroquinolones has been noticeable in recent years (ERAS-Net 2019).

**Figure 28.**
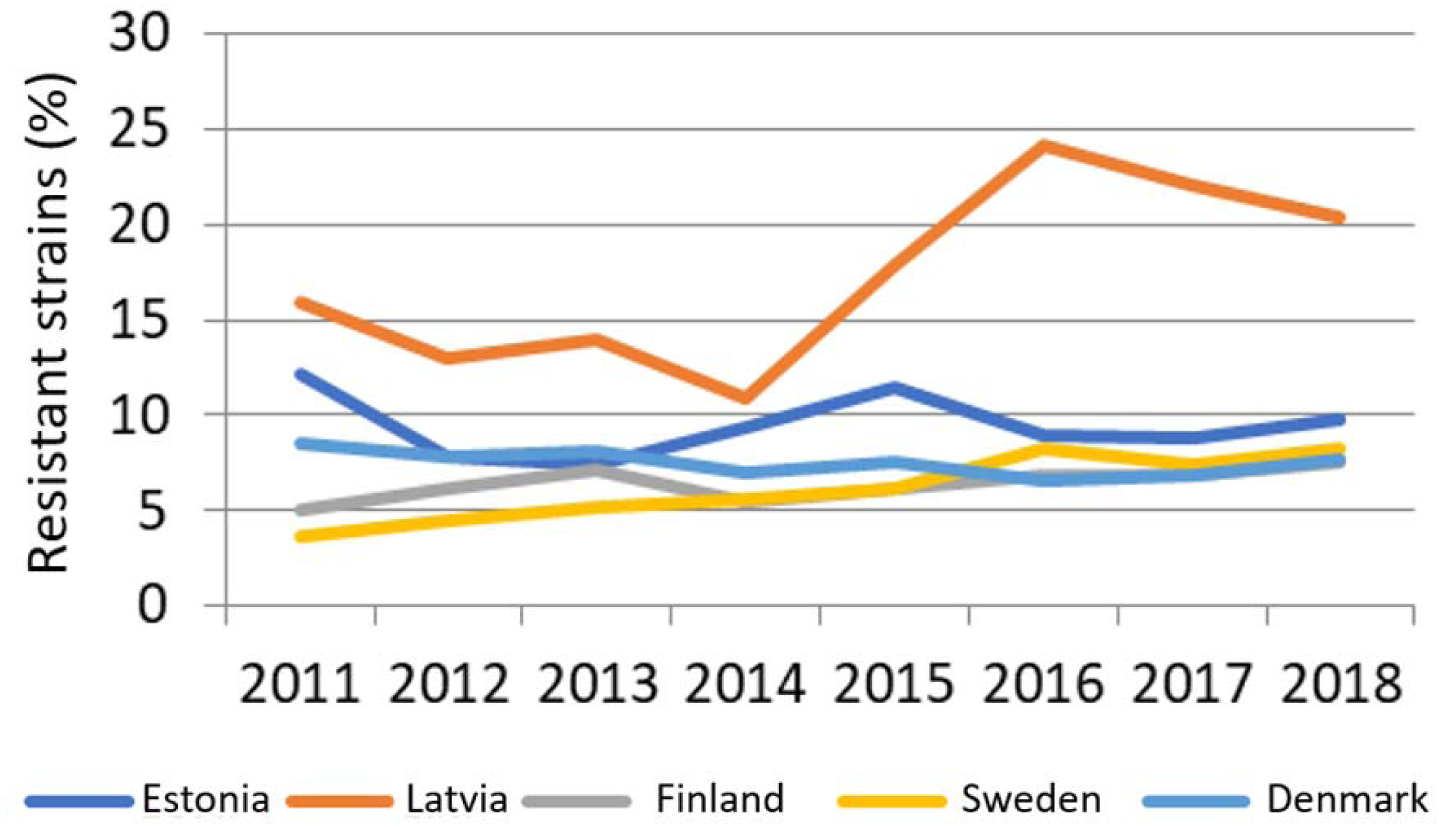
Resistance of *E. coli* to third-generation cephalosporins.

**Figure 29.**
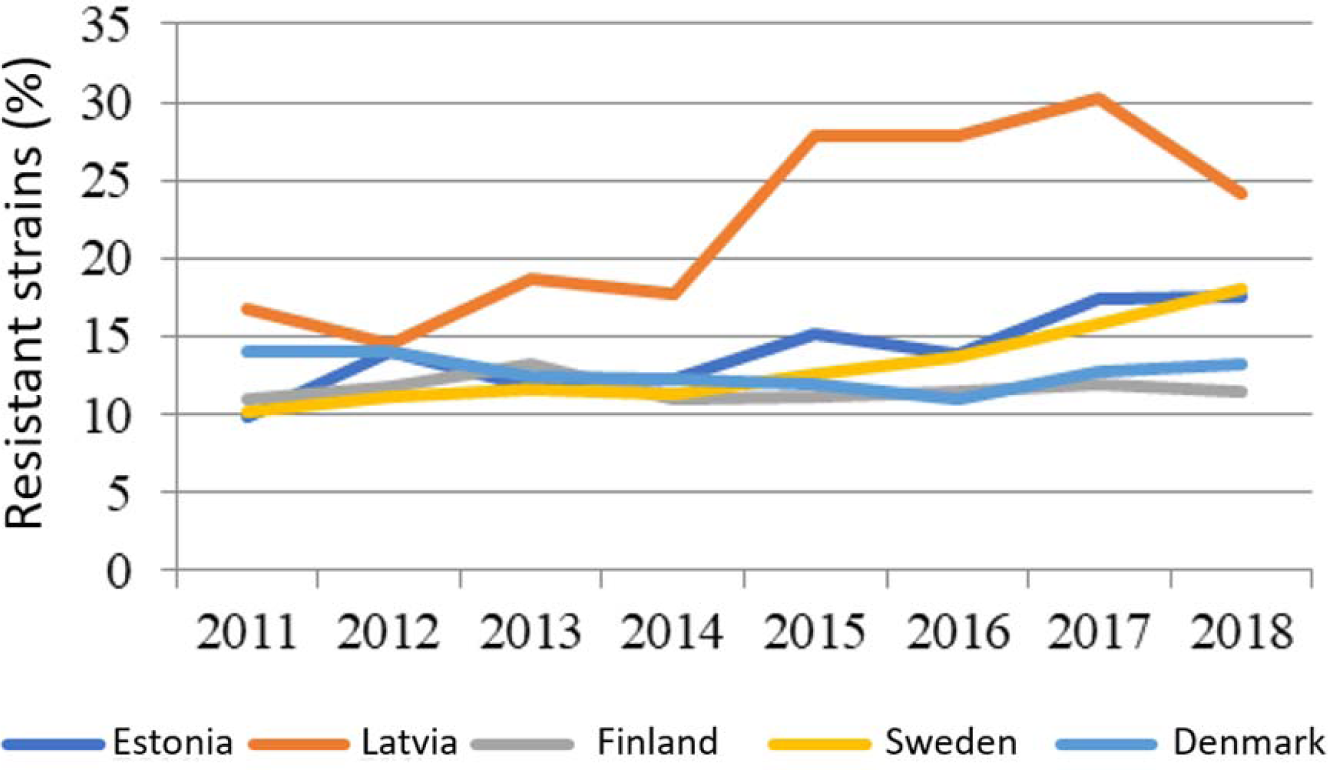
Resistance of *E. coli* to fluoroquinolones.

According to the BEEP (Baltic ESBL Epidemiology Project) study conducted in 2012, ESBL producing *E. coli* strains are more common in Latvia (on average 4.7% in outpatients and 12% in hospitals) and St. Petersburg, Russia (12% and 11.9% respectively). However, differences in ESBL incidence between institutions were large, especially in Russia (ambulatory 0-42%; hospital 5.7-53%).

Resistance of invasive *K. pneumoniae* strains to 3rd generation cephalosporins (ceftazidime and/or cefotaxime) in Estonia has decreased almost four times compared to 2011 (39.5% vs. 13.6%)(Figure 30). Also, in Latvia, in recent years the resistance of *K. pneumonia* to 3rd generation cephalosporins has decreased (EARS-Net 2019). Resistance to fluoroquinolones (ciprofloxacin) remains high (21% in 2018), but decrease in resistance is noticeable (ERAS-Net 2018). In Finland and Denmark, an increase in resistance to fluoroquinolones has been observed in recent years (Figure 31). (EARS-Net 2018, ERAS-Net 2019).

**Figure 30.**
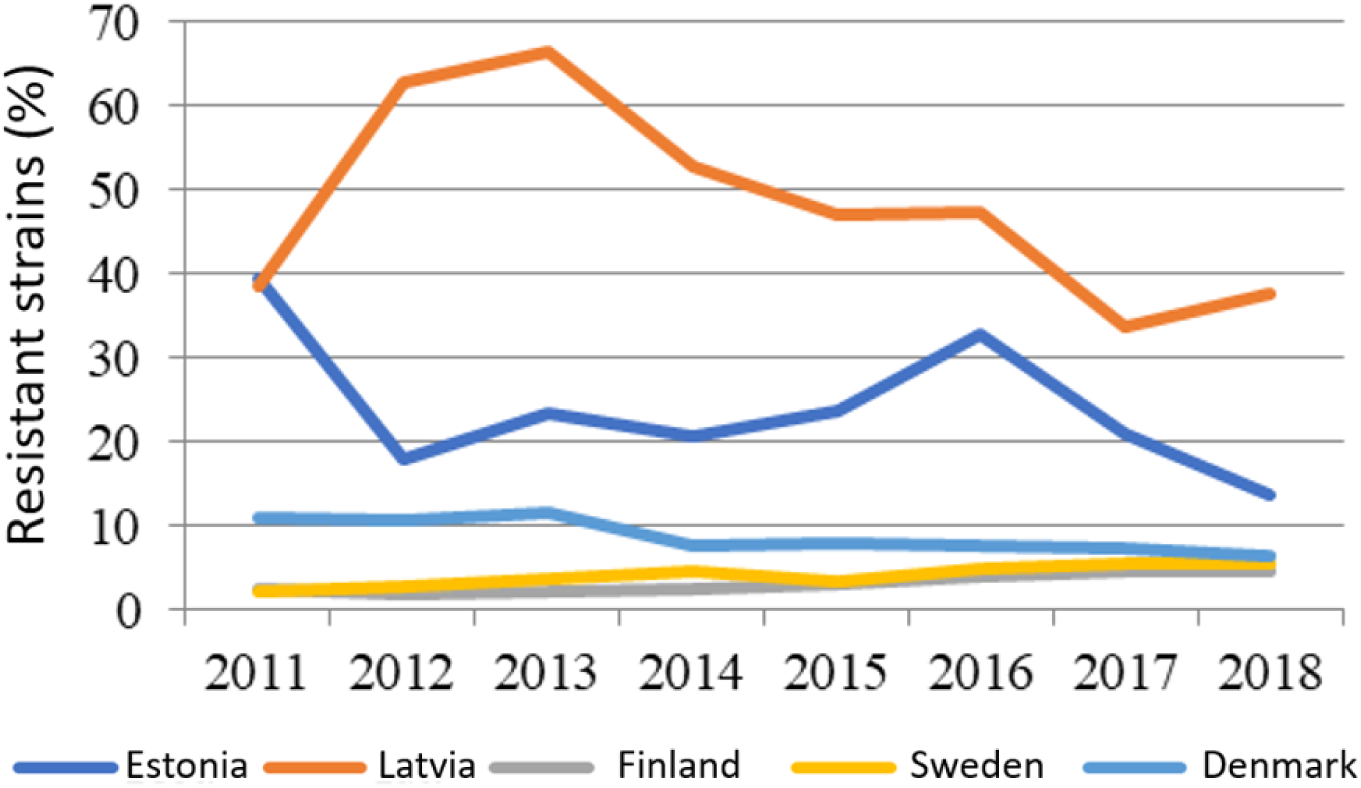
Resistance of *K. pneumonia* to third-generation cephalosporins.

**Figure 31.**
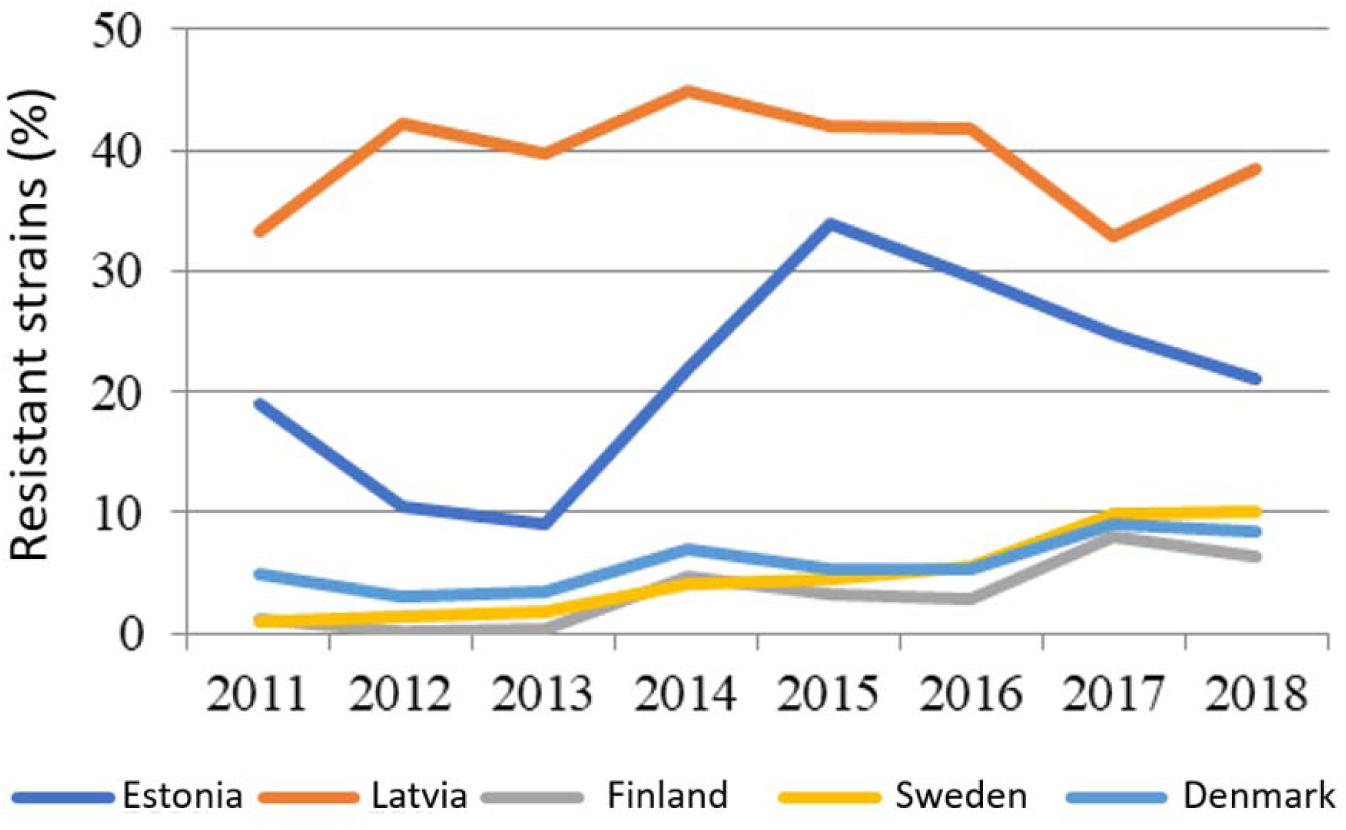
Resistance of *K. pneumonia* to fluroquinolones.

Resistance to aminoglycosides (amikacin, gentamicin) is higher in Estonia than in comparison countries (10% in 2018), but the trend is towards decrease, similar to Latvia (EARS-Net 2019). In 2011-2018, majority of *K. pneumoniae* strains, both in Estonia and in the reference countries were sensitive to carbapenems. This is a good indicator as the European average *K. pneumoniae* resistance rate was 7.5% in 2018.

Resistance of *E. faecalis* to gentamicin and macrolides remains at a high level (25% in 2018, EU average 27.1%). The rate of resistance is slightly higher in Latvia (32% in 2018) and lower in Sweden and Denmark (approx. 12% in both)(Figure 32). All *E. faecalis* strains analyzed in Estonia so far have been sensitive to vancomycin, but in 2018 one strain was resistant to linezolid.

**Figure 32.**
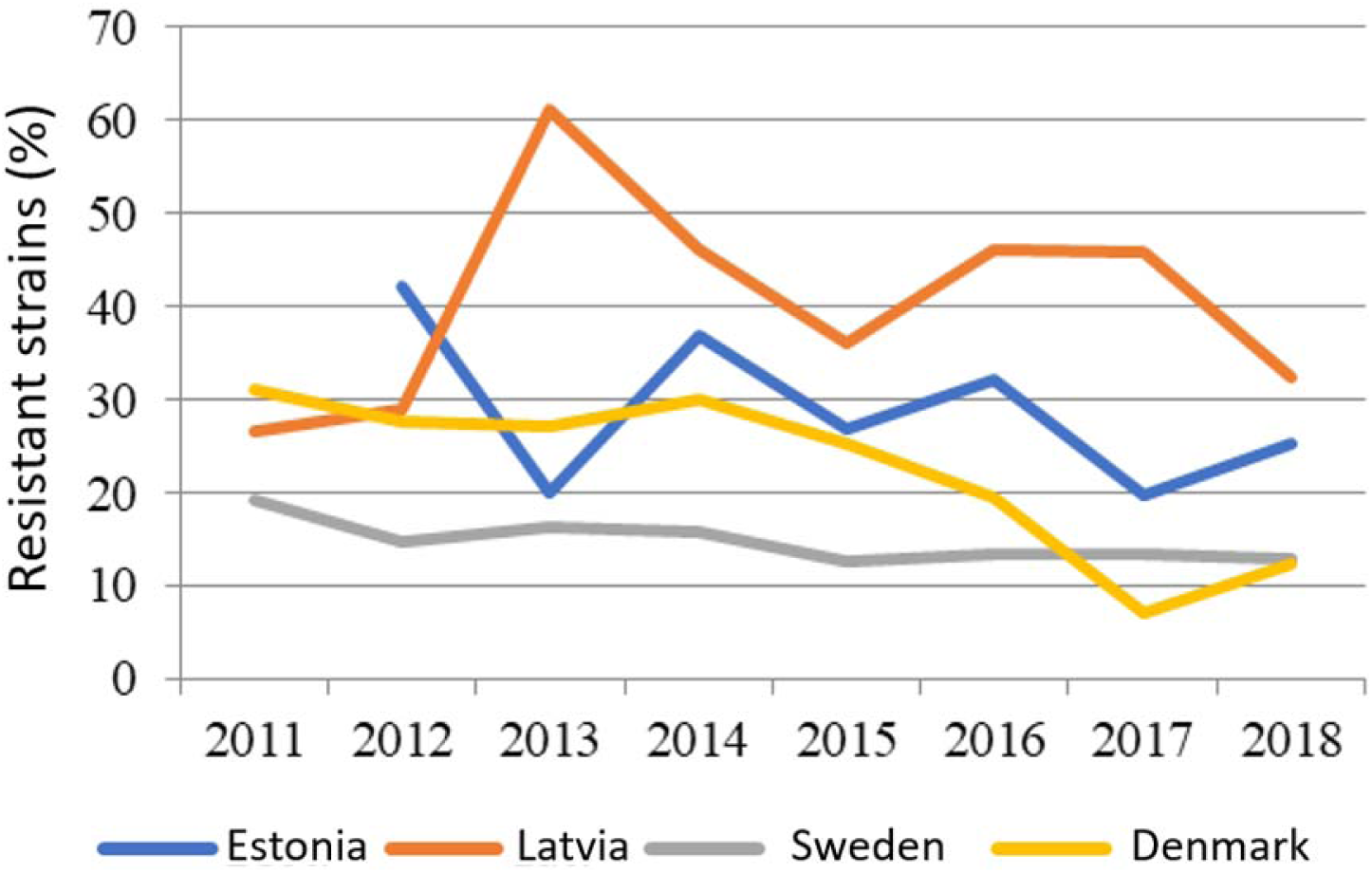
Resistance of *Enterococcus faecalis* to gentamicin.

The proportion of invasive methicillin-resistant *S. aureus* (MRSA) in Estonia in 2011-2018 was 1.7-3.5% (Figure 33). All the strains studied in Estonia were vancomycin sensitive, but in 2018 four strains resistant to linezolid were detected. MRSA is also rare in the comparison countries, but somewhat more frequent in Latvia (5.7% of strains in 2018).

**Figure 33.**
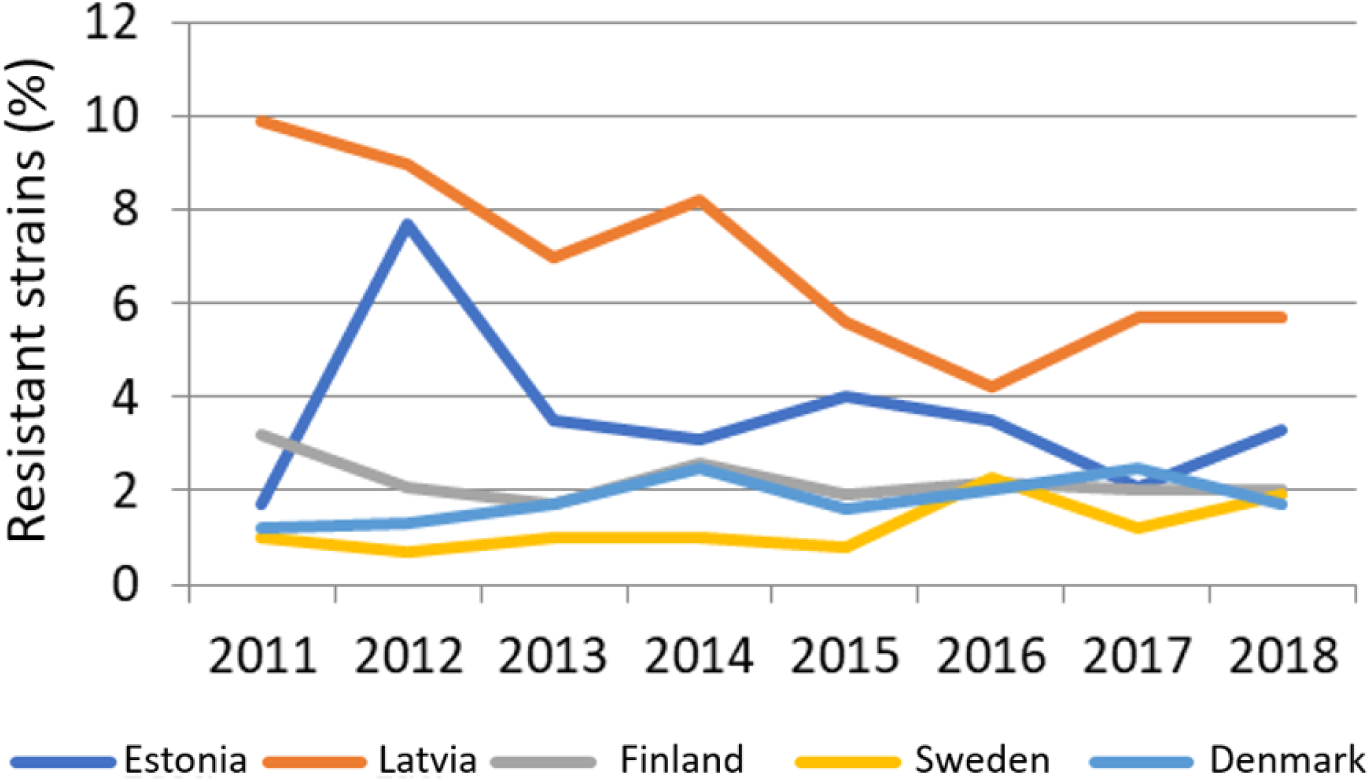
Resistance of *S. aureus* to methicillin, i.e. proportion of MRSA.

In Estonia, the resistance of *Streptococcus pneumoniae* to penicillin is low, and in 2018 no invasive strain was resistant (Figure 34). In Finland, Sweden and Denmark, the proportion of penicillin-resistant strains was lower (1%), but higher in Latvia (4.3%). Resistance to macrolides (erythromycin) is not high in Estonia, although in recent years it had an increasing trend. All strains of *S. pneumoniae* in Estonia were sensitive to 3rd generation cephalosporins (ceftriaxone, cefotaxime) and moxifloxacin. Similar results were also obtained in the reference countries, but in Finland the resistance to macrolides is higher (Figure 35).

**Figure 34.**
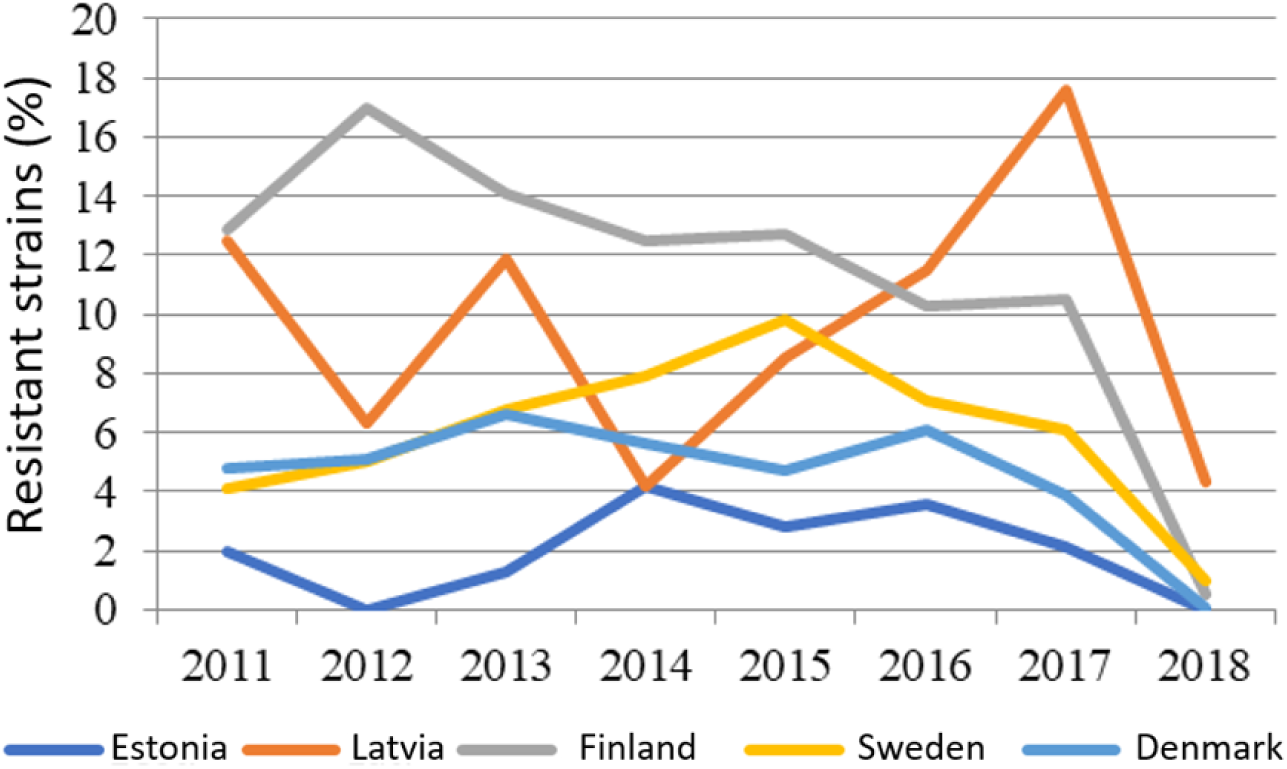
Resistance of *S. pneumoniae* to penicillin.

**Figure 35.**
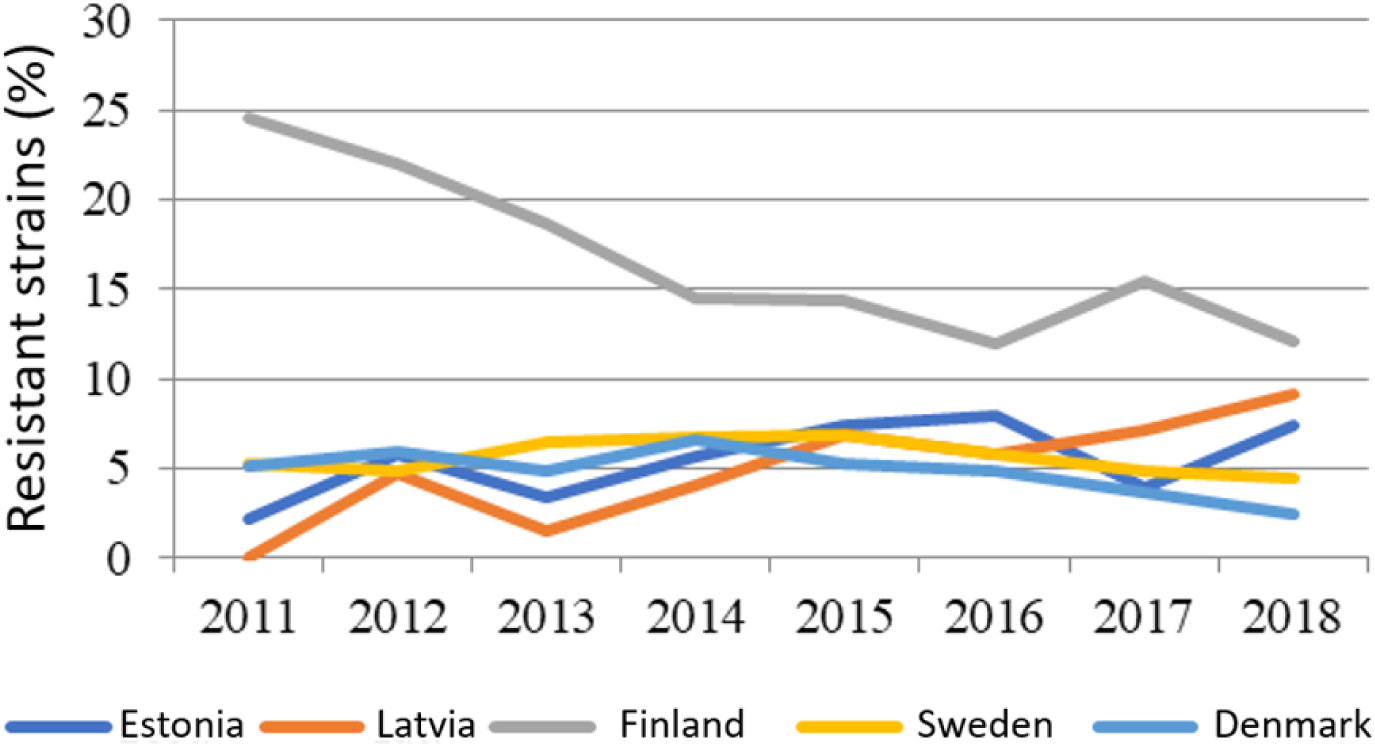
Resistance of *S. pneumoniae* resistance to macrolides.

In summary, the level of resistance in Estonia is relatively low, similar to the Scandinavian countries. Analyzing the prevalence of invasive hospital strains of drug-resistant microbes in Estonia and Europe (EARS-Net reports), it is seen that the resistance level of Gram-positive pathogens (such as methicillin-resistant *S. aureus*, macrolide- and/or penicillin-resistant pneumococcus) has stabilized and even decreased in several countries. The prevalence of these resistant microbes in Estonia is generally low. However, for resistant Gram-negative bacteria (especially ESBL-positive *K. pneumoniae*) an upward trend is observed in Estonia and all Europe.

### 6.5. Summary of factors influencing the development of antibiotic resistance

Antibiotic resistance can be natural or acquired. By natural resistance is meant the natural ability of some bacterial species to neutralize the effects of certain antibiotics. For example, Gram negative bacteria are naturally resistant to several antibiotics because the drug molecules cannot pass through their outer membrane. In the case of acquired drug resistance, microbes become insensitive to antibiotics as a result of mutations or horizontal gene transfer i.e. by exchanging resistance genes with each other. Gene transfer makes possible rapid spread of resistance genes both within the same microbial species and between different species. Three mechanisms of horizontal gene transfer are known: 1) conjugation, in which bacteria exchange DNA through cell-to-cell contact; 2) transformation, where bacteria assimilate from the environment free DNA from dead bacteria; 3) transduction, in which DNA is transferred from infecting bacteria by means of viruses (bacteriophages) (Liu et al. 2022).

The development of resistance is an inevitable side effect of antibiotic consumption. Development and spread of resistance is promoted by the incorrect and excessive use of antibiotics in medicine, animal husbandry and spread of drug residues in the environment. Residues of medicines can enter the environment during manufacturing, use and during disposal of left overs. Humans, animals and birds can metabolize only a portion of the quantities of drugs administered. 40-90% of consumed antibiotics remain undegraded in organisms and excreted into the environment with urine and feces (Polianciuc et al. 2020). It follows that as a result of the use of antibiotics, a significant amount of antibiotics enters the environment. Generally, the concentrations of antibiotics present in the environment are sub-inhibitory, i.e. they have no antimicrobial effect. On the other hand, it is known that subinhibitory antibiotic concentrations trigger in bacteria mechanisms related to increase in mutation frequency, activation of gene transfer and biofilm and persister formation (Bernier and Surette 2013). Thus, antibiotic residues in the environment cause selective pressure that enhances the development and spread of drug resistance. Moreover, antibiotics are excreted into the environment together with intestinal microbes who became antibiotic resistant during the course of treatment.

Wastewater treatment plants are considered as main reservoirs of drug-resistant microbes and resistance genes, as there a large number of microbes comes into contact with subinhibitory antibiotic concentrations. Large number of bacteria and sufficient nutrients content create good conditions for microbial reproduction and horizontal gene transfer to take place. In a recently conducted CWPharma study, four antimicrobials were found in the sludge of Estonian wastewater treatment plants: fluconazole, lincomycin, ofloxacin and trimethoprim (Henning et al. 2020). In addition, wastewater contains disinfectants and heavy metals, which in turn increases the selection pressure for spread of antibiotic resistance.

The links between antibiotic use and antibiotic resistance take a long time to develop and are usually expressed as differences between large geographical regions. In Europe there are national differences in the use of antibiotics in human medicine, which are reflected in resistance levels. In Estonia, the general use of antibiotics is one of the lowest in Europe, therefore, further activities should be aimed at maintaining the level, not so much at lowering it. At the same time, current study highlights details that cause concern. Examples here are large differences in antibiotic use between different hospitals or widespread use of restricted antibiotics in some central and local hospitals. Differences were also observed in the choice of antibiotics when comparing similar departments in different hospitals. The situation is similar in the use of ambulatory antibiotics, where unjustified differences where observed in the comparison of regions and specialities. The unwarranted use of later generation antibiotics (mainly cefuroxime) also deserves attention, as it increases the risk of development of multi-resistant Gram-negative bacteria (Dancer 2001). In this context, presence of Antibiotic Surveillance Service in all hospitals is of critical importance.

As a parameter affecting the level of resistance, the hygiene of the hospital should be noted. This influences the spread of pathogens at hospital level and thus the use of antibiotics and the development of resistance. Improvements in hygiene have helped to keep MRSA levels under control around the world, but the threat of continued increase of resistance in Gram-negative bacteria remains to be managed. Monitoring the use of antibiotics in hospitals is of critical importance - it must be done by the infection control doctor in cooperation with the hospital pharmacy.

In a globalized world, there is always the risk that the resistance problems of the rest of the world will reach people in regions with currently low levels of resistance. This can occur through the global movement of people and goods. For example, the NDM-1 gene, which causes carbapenem resistance probably originates from India, but was first described in European tourists visiting India (Charan et al. 2012). In Asia *Neisseria gonorrhoeae* has become highly resistant, creating a reservoir from where resistant strains can spread to other regions. In Estonia the resistance levels of gonorrhea are currently not tested. Considering the large picture, introduction of resistance testing is needed. Also, resistant strains can reach us with various goods, for example food and animal feed.

In Estonia the use of antibiotics in the treatment of farm animals is prevalent because the level of bacterial infections across all diseases is high. In current study, it was revealed that antibiotic treatment length, choice of active substances and indication for treatment varied in a very wide range between farms. The use of wide range of active ingredients and unwarranted treatment decisions cause increase in resistance. For the treatment of farm animals mainly first option active substances (beta-lactams and tetracyclines) were used. The biggest problem is unjustified use of 3.-4. generation cephalosporins that possibly creates pressure for the development of resistance in Gram-negative bacteria. It was found that the probability of finding ESBL producing *E. coli* is 2.5 times higher in cattle farms with high annual usage as compared to the farms with low usage. In pig farms the association was not found; this may be due to the small sample size.

One of the routes of transmission of antibiotic resistance is the transfer of resistance genes to humans through food. Travel and global trade influence the spread of resistance. Thereby, reasonable use of antibiotics locally does not avoid the importation of resistant strains. For example, it was found in this study that the incidence of ESBL-positive *E. coli* was significantly higher in imported meat than in domestic meat. Certain beta-lactamases were only present in imported meat (blaCARB-2, blaCTX-M27, blaCTX-M32). It has to be noted that blaCTX-M27 was also detected in 11.4% of human isolatesAntibiotic resistance genes are often inherited together with genes that confer resistance to disinfectants or metals (Karkman et al. 2018). This means that the use of certain disinfectants promotes the spread of antibiotic resistance. Well-known examples here include triclosan, which used in consumer goods and chlorhexidine, a disinfectant widely used in hospitals. Therefore, it is necessary to carefully monitor which disinfectants to choose. The use of metals is an additional problem problem. Silver and copper surfaces and metal salts have a germ-killing effect. In hospitals silver and copper surfaces are used to keep microbes away. Unfortunately, the emerging niche can be taken over *Acinetobacter baumanni* (Manchanda et al. 2010). In the currentt study, we did not analyze *A. baumanni*, but the issue definitely needs further analysis. This microbe is highly resistant to metals and is becoming resistant to more and more antibiotics. The use of various metal salts is widely used in animal husbandry, which can also cause the spread of antibiotic resistance.

## 7. Recommendations to reduce the spread of AMR

The first recommendation is the creation of a Competence Center that would deal with the topic of AMR across all fields. This action should also include funding for research. Dealing with AMR on a project basis is unfruitful.

### 7.1. Recommendations to reduce AMR in human medicine

In Estonia, there is a need to create real-time, sustainable AMR monitoring, which (1) collects and analyzes on continuous basis antimicrobial use and antimicrobial resistance data and provides regular feedback to relevant institutions (state, healthcare and research institutions), (2) evaluates the reliability of the data and ensures their quality by carrying out additional and confirming research, if necessary, (3) coordinates national and international monitoring networks and scientific projects.

Outputs to be achieved:

- Current data on antimicrobial resistance and antibiotic use are available and can be analyzed according to different parameters.
- There is a regular analysis of the aforementioned data and feedback to the relevant authorities.
- Antimicrobial treatment guidelines are regularly updated based on antimicrobial resistance and antibiotic usage data.
- Based on databases, “alarm situations” (eg outbreaks, certain drug-resistant microbes) are detected automatically.Corresponding risk analysis and counter-measures are implementated rapidly.
- Data are automatically transmitted to the monitoring networks of ECDC and others.
- AMR reference laboratory provides support for diagnostic laboratories with confirmatory tests and quality control measures.

### Specific recommendations for improving the use of antibiotics in human medicine

#### 7.1.1. Compliance with AWaRe classification

It is necessary to analyze what is the reason for the use of restricted antibiotics. It is necessary to clarify whether it is based on existing guidelines or due to their absence and how much it would be possible to replace the usage of restricted antibiotics with first-line drugs. This could be achived by changes in treatment guidelines. Hospital doctors also need to be trained because the AWaRe classification is new and needs introduction. AWaRe classification gives hospitals infection control doctors a new opportunity to monitor the use of antibiotics, set goals and make plans to encourage more rational antibiotic use. A national goal can be set, that 60% of antibiotic use in hospitals would constitute the first-line (Access) AWaRe classification category antibiotics. Compliance with the AWaRe classification should be among the quality indicators monitored by Health Insurance Fund.

#### 7.1.2. Availability of treatment guidelines and infection control doctors

A significant deficiency in the field of guidelines was also indirectly indicated by the 2019 hospital data report, which revealed that there are large variations in antibiotic use between hospitals and departments of the same type. At the national level, it is necessary to pay attention to antibiotic treatment guidelines and the availability of training in hospitals and the presence of an infection control doctor in all hospitals.

#### 7.1.3. Switching from intravenous to oral antibiotics

At present, antibiotic courses are mostly administered intravenously in hospitals. One of the methods to optimize the use of antimicrobial therapy is to encourage a switch to oral therapy, following the relevant instructions.

#### 7.1.4. Reducing the use of penicillins combined with a beta-lactamase inhibitor

Although the problem concerns different diagnoses, we still observed that for teeth and jaw diagnoses in almost a third of the cases penicillin combinations with beta-lactamase inhibitor were used. In Estonia, there is no antibacterial treatment guideline for oral and dental infections. The existence of treatment guidelines should be more thoroughly analyzed and the creation of missing treatment guidelines initiated.

#### 7.1.5. Adherence to treatment guidlines

To ensure adherence to treatment guideline recommendations, regular auditing of antibiotic use, feedback and continuous training of doctors are important.

### 7.2. Recommendations to reduce AMR in veterinary medicine

Reducing the use of category B (restricted) antibiotics is a complex series of activities, which must include more effective surveillance, increased awareness of veterinarians, change of behavior patterns and contribution of food producers.

#### 7.2.1. Implementation of nationally supported herd health programs to decrease the need of antibiotic treatment

Preventing diseases and improving the farm environment is a prerequisite for reducing infection pressure and thereby morbidity of animals. Improving herd health with disease prevention activities reduces the need for antibiotics. Nationally supported animal health programs are a mean by which animal health improves and the need for treatment decreases.

#### 7.2.2. Create a species-specific e-database of antibiotic use to implement recorded, analyzed and evaluated antibiotic treatment

Since the amount of antibiotics used, the rate of morbidity and the frequency of treatment are highly variable between herds, it is currently not possible to evaluate antibiotic treatment practices without a national registry of antibiotic use. In addition to registration, reference indicators must be developed, which, if exceeded, trigger implementation of counseling and supervision activities. From 2023 farm-based registration of antibiotic use in production animals is mandatory. The data are transferred to a national database, but at the same time there is no central aggregation and analysis of the collected data. Therefore, it is not possible to assess which animal species were treated, based on which diagnosis and in what amounts antibiotics were used.

#### 7.2.3. Establish an effective and sustainable national surveillance system for the evaluation of antibiotic treatment

Antibiotic treatment can only be prescribed by a licensed veterinarian, which means that the treatment decision and justification is within the competence of veterinarians. One option could be introduction of treatment service contracts. Since antibiotics can only be prescribed by a licensed veterinarian and medications can be purchased directly from wholesale companies, treatment service contract should be concluded between each livestock company and the serving veterinarian. This gives the supervisory authority opportunity to assess appropriate use of medicines bought in bulk and assess the competence of the veterinarian in the use of antibiotics and compliance to treatment guidelines. The control over the laws governing antibiotic treatment needs to be made more efficient. For this purpose, supervisory officials must be trained and human resources must be allocated for this activity. Based on the results of farm-based and veteterinarian-based data analysis, it is possible to conduct counseling activities or monitoring activities to determine the reasons for excessive or unreasonable antibiotic treatment.

The introduction of additional regulations on the sale of veterinary medicinal products will help to make supervision more efficient. A requirement must be established that veterinarians can order medicines from wholesale companies only based on a digitally signed order. This would rule out the possibility that drugs are sent to farms on the order of someone other than a veteterinarian, and the order form will be processed later. Such an act is well known, although the parties deny it. Considering the current technology, requiring a digital signature for the order is not limitating anyones lawful actions, but it is allows better control over medication use.

#### 7.2.4. Trainings and counseling system to improve the awareness of veterinarians and animal breeders

In addition to the improvement of antibiotic treatment rules and supervision, there is a need to continue to improve veterinarians and animal breeders’ awareness of antibiotic treatment. Conducting continuing education in smaller groups, motivation of veterinarians through professional organizations and targeted actions for promoting guidelines can result in changes of attitudes and behavior. The role of professional organizations and universities must be increased to advise and train veterinarians regarding the use of antibiotics. A special training program must be created for veterinarians advising on livestock health and current continuing education programs should be continued and improved. In addition to professional skills, veterinarians need to upgrade their communication and teamwork skills, as antibiotic treatment decisions are also relevant for entrepreneurs engaged in animal husbandry. The measures introduced can be evaluated with the help of studies on antibiotic consumption and reasons behind specific treatment decisions. These studies need to be continues, therefore also requiring continuous financing.

#### 7.2.5. Initiative of food producers and measures to obtain higher quality food raw materials

We recommend creating milk and meat quality classes that consider veterinary treatment practices and herd health situation in the farm. Promotion of domestic food and higher quality indicators guide consumers to make choices and give competitive advantage to goods produced in Estonia. Commitment of food manufacturers to agreement with the animal breeders enables implementation of the principle that local food comes from herds in good health where the use of antibiotics is minimal and based on need. It can happen only if food producers take the initiative and recognize farms where the use of antibiotics is minimal and animal welfare is at a high level.

#### 7.2.6. Antibiotic Resistance Surveillance System and National AMR Program

We recommend supplementing the annual monitoring program and starting to issue a consolidated AMR report similarly to the reports in Nordic contries. The European Union’s AMR surveillance is sufficient to provide an overview of *E. coli* resistance in clinically healthy pigs and broilers in Estonia. However, the European Union programs do not collect data on pathogens isolated from sick animals. In addition, many animal and microbial species important in veterinary medicine are not covered. In addition to their zoonotic significance, European Food Safety Agency has pointed out the following critical antibiotic resistant infections causing animal diseases: *E. coli*, *E. faecalis* and *E. cecorum* in poultry, *E. coli* and *Brachyspira hyodysenteriae* in pigs, *E. coli, S. aureus* and *Rhodococcus equi* in horses, *E. coli* and *S. aureus* in cattle, *E. coli* in sheep and goats, *Staphylococcus pseudintermedius*, *E. coli* and *P. aeruginosa* in dogs and cats. Surveillance of this list of animals species and pathogens is necessary for assessing AMR level in the country. The national AMR program helps to guide the responsible use of antibiotics and to evaluate the effectiveness of the introduced measures. In addition to the European Union monitoring program, every year the level of antibiotic resistant microbes should be evaluated in at least one animal species. A consolidated report, which includes both the use of antibiotics and the surveillance of AMR in the European Union as well as national AMR surveys should be completed regularly. The report will contribute to the implementation of the AMR reduction agenda.

### 7.3. Reducing AMR in the environment

One of the possible routes of pharmaceuticals entering the environment is that people do not dispose unused medications correctly. At this point, consistent outreach activities are necessary. Currently, pharmacies have an obligation to take back unused medicines, but this is an extra work. Withdrawal of unused medications should be made more attractive to pharmacies.

Residues of medicines reach the environment both through waste water treatment system (waste water, sewage sludge compost) and with manure. Determining more precise parameters of the processes need further research. In the case of sewage sludge, the efficiency of different composting technologies must be considered. In the case of manure, degradation of drugs during storage should be investigated. In addition, necessity of adding composting technology should be considered. It is also necessary to know how drug residues can move from the soil to the environment. The influence of soil properties and precipitation needs to be determined. Processes in karst areas need special attention because there the drug residues and microorganisms move into groundwater more easily.

The current study was based on observing functioning production and processing systems. It turned out that sometimes it is not possible to obtain accurate information from the farms. For example, information about liquid manure storage and spreading times is often not recorded in detailed manner. A more detailed study of the key issues influencing the release of medicines into the environment would be necessary by using an experimental approach - the use of experimental reactors and experimental fields.

## 8. Summary of results in the light of “One Health” approach

Microorganisms resistant to antibiotics occur in humans, animals and in the environment. Resistant microorganisms can spread between animals to humans and through direct contacts or through the environment. According to the “One Health” principle, human and animal health and the state of the environment are closely related. Following the One Health action plan of European Union from 2017 was one of the goals of current project..

Cooperation and coordinated data collection and analysis involving different One Health segments is important in order to understand the scope of a possible problem and to assess whether antibiotic use and AMR are related. “One Health” approach requires integrated actions to reduce antibiotic resistance in medicine, in veterinary medicine as well as in the environment.

As a result of the current project, we recognize that the use of antibiotics in Estonia is generally low when compared to other European countrieso. At the same time, there are bottlenecks that concern both human and veterinary medicine. In both cases, we have to admit that sometimes the treatment guidelines are missing, sometimes the guidlanes are not followed and antibiotics are used for the wrong indications. This is often related to the insufficient support from specialists. For example, many hospitals lack an infection control specialist. Major concerns are the unjustified use of broad-spectrum antibiotics in humans and the large use of antibiotics critical for human medicine (cephalosporins, quinolones) in animal treatment.

If antibiotics are used in higher amounts, resistance will also develop and spread. We observed that in the cattle farms where more cephalosporins are used, there is also a higher level of ESBL-mediated resistance. We also see that close intraspecies genetic clusters of bacteria are common in humans and animals. This is evidence of transfer of microbes between species. However, such a transfer takes place slowly and we did not detect transmission events from the previous few years.

Antibiotic residues, like other drug residues, end up in the environment. The main contributors are use of manure and composted sewage sludge as fertilizer. We detected comparable concentrations of fluroquinolones and tetracyclines in manure and in uncomposted sewage sludge. Composting reduces the content of drug residues and the efficiency of the process depends of the technology used. At the same time, composting of sewage sludge is mandatory when it is used as fertilizer, but processing of manure is not. Therefore, the use of untreated manure can release more antibiotic residues into the environment than using treated sewage sludge. In addition, we also detected some other drug residues, in addition to antibiotics, that accumulate in the environment. High levels of diclofenac and carbamazepine in surface water are a concern. These medicines are used only in humans, therefore they move into the environment only through sewage treatment plants.

In the course of the project, we established that both farms and hospitals collect data in very different formats. Thus, data collection and analysis should be improved. We suggest that One Health competence center for antibiotic resistance will be established. The competence center would provide input into the development of treatment guidelines and provides advice in more severe epidemiological situations.

## 9. Methods

### 9.1. Collection of microbial strains in human medicine

The study was carried out in eight health care institutions participating in the European Antimicrobial Resistance Surveillance Network (EARSNet) (Tartu University Clinic, Northern Estonian Regional Hospital, Western Tallinn Central Hospital, Pärnu Hospital, East-Tallinn Central Hospital, East-Viru Central Hospital, Narva Hospital, SYNLAB Estonia).

Hospitalized patients and outpatients with ESBL-positive isolates were included in the study of *E. coli, K. pneumoniae* complex (*K. pneumoniae, K. variicola, K. quasipneumoniae subsp. quasipneumoniae, K. quasipneumoniae subsp. similipneumoniae*), methicillin-resistant *S. aureus* (MRSA) and vancomycin-resistant *E. faecium* (VRE). For each patient with a resistant strain, a patient with a sensitive microbial strain from the same medical institution or in ambulatory setting a consecutive patient with a sensitive microbial strain (ESBL-negative *E. coli, K. pneumoniae*, methicillin-susceptible *S. aureus* or vancomycin-susceptible *E. faecium*) was included. Enterobacteriaceae from hospitalized patients were isolated from blood, and strains from ambulatory patients were isolated from urine. *S. aureus* and *E. faecium* strains were isolated from various clinical materials (e.g. blood, urine, wound). During the study period (2019-2022) 202 pairs of resistant and sensitive strains, i.e. 404 strains, were collected.

### 9.2. Determination of antibacterial sensitivity

Collected strains were identified with a mass spectrometer (MALDI-TOF; Bruker) and ESBL/AmpC/Carbapenemase strains were screened according to EUCAST (European committee of antimicrobial susceptibility testing) guidelines. To confirm MRSA and VRE, the *mec*A and *mec*C genes of *S. aureus* strains and the *van*A and *van*B genes of *E. faecium* strains were determined. MIC (minimum inhibitory concentration) was determined using the broth dilution method on a Phoenix analyzer (Becton Dickinson). The criteria of EUCAST were used to determine resistant and sensitive strains.

### 9.3. Statistics

The Wilcoxon rank-sum test was used to compare the MIC of microbial strains, and Fisher’s exact test was used to compare sensitive and resistant strains. We considered it statistically different if the p-value was <0.05.

### 9.4. Bioinformatics

#### 9.4.1. Genome assembly and quality

Sequencing reads of each sample were analysed with StrainSeeker (Roosare et al 2017) to ensure that the reads are not from a mixed populations. Samples containing more than 10% of non-target bacterial reads were removed from following analysis. Reads were filtered with fastp (Chen at al 2018) and assembled with SPAdes (Prjibelski et al 2020) using isolate mode. Contaminated contigs were removed based on blastn search against NCBI nt database. Assembly quality and completeness was checked with BUSCO (Seppey et al 2019).

#### 9.4.2. MLST analysis

Multi-locus sequence typing (MLST) analysis was carried out both on raw reads and assembled contigs using stringMLST v0.6.3 (Gupta et al 2017) and mlst software (Seemann, https://github.com/tseemann/mlst) correspondingly based on PubMLST data (https://pubmlst.org/) (Jolley & Maiden 2010).

#### 9.4.3. Phylogenetic analysis

Alignments for core genome trees and SNP trees were calculated with Harvest suite parsnp software (Treangen 2014). Core genome trees were calculated with RAxML-NG v.1.0.1 with using model GTR+G (Kozlov et al 2021) and SNP trees were based UPGMA algortihm utilized in MEGA 11 software (Tamura et al 2011). Phylogenetic trees were visualized in iTOL web application (Letunic and Bork 2021).

#### 9.4.4. Resistance genes detection

Acquired resistance genes were detected with ResFinder 4.0 (Bortolaia et al 2020).

Chromosomal or plasmid origin of each resistance gene in long enough contigs was determined with an in-house pipeline for plasmid detection including MobSuite software (Robertson and Nash 2018).

## Funding

The study was commissioned and funded by the Estonian Research Agency with support from the European Regional Development Fund of the program “Strengthening of sectoral research and development” (RITA) activity 1 “Strategic TA supporting activity”. The study was completed in the period July 1, 2019 - June 31, 2022 to implement the objectives of the Ministry of Rural Affairs, the Ministry of Social Affairs the Ministry of the Environment. The research budget was 947,370 euros.

## Data Availability

All data produced in the present study are available upon reasonable request to the authors

## Acknowledgement

We thank Irja Roots and Kaur Tenson for assistance in translation of the document.

## Abbreviations

AMR: antibiotic resistance (antimicrobial resistance)
ATC classification: anatomical-therapeutic-chemical classification
DDD: defined daily dose
ECDC: European Center for Disease Prevention and Control control
ESAC-Net: European surveillance network of consumption of antimicrobial substances (European Surveillance of Antimicrobial Consumption Network)
ESBL: extended-spectrum beta-lactamase
EUCAST: European Committee for Antimicrobial Susceptibility Testing
MIC: minimum inhibitory concentration
MRSA: methicillin-resistant *Staphylococcus aureus*
PCU: population correction unit
PAP: purchased antibiotic prescription
ST: sequence type
BD: bed day
VRE: vancomycin-resistant enterococcus

## Notes

### Competing Interest Statement

The authors have declared no competing interest.

### Author Declarations

University of Tartu Ethics Committee

